# Triangulating Causality Between Childhood Obesity and Neurobehavior: Twin and Longitudinal Evidence

**DOI:** 10.1101/2022.04.12.22273769

**Authors:** Leonard Konstantin Kulisch, Kadri Arumäe, D. A. Briley, Uku Vainik

**Affiliations:** Institute of Psychology, University of Tartu, 50409 Tartu, Estonia; Leipzig University, 04109 Leipzig, Germany; Department of Psychology, University of Illinois at Urbana-Champaign, Champaign, IL 61820, USA; Institute of Genomics, University of Tartu, 50409 Tartu, Estonia; Montreal Neurological Institute, McGill University, Montreal, QC H3A 2B4, Canada

**Keywords:** childhood obesity, causality, twin modeling, cross-lagged panel modeling, neurobehavior

## Abstract

**Objective:** Childhood obesity is a serious health concern that is not yet fully understood. Previous research has linked obesity with neurobehavioral factors such as behavior, cognition, and brain morphology. The causal directions of these relationships remain mostly untested.

**Methods:** We filled this gap by using the Adolescent Brain Cognitive Development study cohort comprising 11,875 children aged 9–10. First, correlations between body mass percentile and neurobehavioral measures were cross-sectionally analyzed. Effects were then aggregated by neurobehavioral domain for causal analyses. Direction of Causation twin modeling was used to test the direction of each relationship. Findings were validated by longitudinal cross-lagged panel modeling.

**Results:** Body mass percentile correlated with measures of impulsivity, motivation, psychopathology, eating behavior, and cognitive tests (executive functioning, language, memory, perception, working memory). Higher obesity was also associated with reduced cortical thickness in areas of the frontal and temporal lobe but with increased thickness in parietal and occipital brain areas. Similar although weaker patterns emerged for cortical surface area and volume. Twin modeling suggested causal effects of childhood obesity on eating behavior (β=.26), cognition (β=.05), cortical thickness (β=.15), and cortical surface area (β=.07). Personality/psychopathology (β=.09) and eating behavior (β=.16) appeared to causally influence childhood obesity. Longitudinal evidence broadly supported these findings. Results regarding cortical volume were inconsistent.

**Conclusions:** Results supported causal effects of obesity on brain functioning and morphology, consistent with effects of obesity-related brain inflammation on cognition. The present study highlights the importance of physical health for brain development during childhood and may inform interventions aimed at preventing or reducing pediatric obesity.

## Introduction

Childhood obesity is a growing health issue with every fifth child in the U.S. having obesity and these numbers expected to rise (Wang et al., 2020). High body weight as indexed by body mass index (BMI) tends to persist into adulthood and is associated with individual health risks and increased financial burden to the health and labor system (Hamilton et al., 2018; Llewellyn et al., 2016). Since all food intake happens through behaviors, such as choosing and eating it (Blundell & Finlayson, 2004), it is hoped that examining the association between obesity and neurobehavioral factors like behavior, cognition, and brain structure could aid in designing interventions that prevent or reduce childhood obesity.

Many studies have found links between such neurobehavioral factors and obesity. Regarding self- and parent-reported traits, correlations have been found between pediatric weight status and mental health issues like depression (Sutaria et al., 2019) as well as various eating behaviors (Kininmonth et al., 2021). Child and adolescent obesity have also been found to correlate with behavioral and self-reported measures of impulsivity (Thamotharan et al., 2013), motivation, and cognitive functioning (Pearce et al., 2018). Regarding brain morphology, associations between childhood obesity and cortical thickness have been found in diverse brain regions. Many studies have reported association to areas within the frontal lobe (Kim et al., 2020; Prats-Soteras et al., 2020; Rajan et al., 2021; Ross et al., 2015; Saute et al., 2018; Thapaliya et al., 2021), but other brain regions may also play a role (Steegers et al., 2021). In participants with higher obesity, cortical surface area was observed to be increased in the left rostral middle frontal gyrus and the right superior frontal gyrus (Prats-Soteras et al., 2020) while others found cortical volume to be altered in frontal and temporal brain regions (Alosco et al., 2014; Hashimoto et al., 2015; Kennedy et al., 2016).

However, it is unclear if obesity may have causal associations with these neurobehavioral domains and aspects of brain morphology and, if so, what the direction of these associations may be. A recent study in adults showed that neurobehavioral correlates of obesity are largely heritable (Vainik et al., 2018) and follow-up genetic analyses with two different methods suggest that obesity may cause variance in personality traits (Arumäe et al., 2021). In childhood, obesity is assumed to be causally influenced by eating, lifestyle, and socio-cultural behaviors and to have causal effects on cognitive abilities (Sahoo et al., 2015) and brain morphology (Alosco et al., 2014; Prats-Soteras et al., 2020). Psychopathology is conceptualized to have a reciprocal relationship with child weight status (Sahoo et al., 2015). However, few studies have tested these causal assumptions.

Genetically informed analyses revealed directional effects from polygenic scores for BMI to ADHD (Hübel et al., 2019) and overeating (Herle et al., 2021), as well as cortical volume (Lugo-Candelas et al., 2020). Anorexia nervosa genetic propensity was identified to predict childhood obesity (Hübel et al., 2019). Another genetically informed study found a reciprocal causal relationship between BMI and disordered eating (Reed et al., 2017).

Longitudinal evidence suggests causal links from BMI to internalizing problems (Bradley et al., 2008) and depression (Korczak et al., 2013). Additionally, inhibitory control (Anzman-Frasca et al., 2015), reward responsiveness (De Decker et al., 2017), prosociality (Christensen et al., 2019), depression (Blaine, 2008; Korczak et al., 2013), aggression (Derks et al., 2019), psychopathology in general (Camfferman et al., 2016), emotional eating (Shriver et al., 2019), picky eating (Antoniou et al., 2016), and cortical surface area (Park et al., 2020) were found to longitudinally influence childhood obesity. Therefore, a reciprocal relationship between childhood obesity and depressive symptoms seems plausible. Longitudinal studies found no evidence for any causal relations between BMI and intelligence (Belsky et al., 2013).

In this study, we sought to triangulate causality between obesity and its well-known correlates to gain further understanding about potential causes and consequences of childhood obesity. First, we aimed to validate previous cross-sectional obesity–neurobehavior associations using a large population-based study dataset. Childhood obesity was expected to correlate with impulsivity, motivation, prosociality, psychopathology, eating behavior, cognition, cortical thickness, cortical surface area, and cortical volume. The present investigation then aimed to shed light on the causal relationships driving these correlations. Twin and longitudinal designs were chosen for that task. Two causal inference methods with different assumptions on separate subsets of the data can aid in establishing robust causal effects (Munafò & Davey Smith, 2018). Based on previous findings, childhood obesity was expected to influence cortical volume while cortical surface area was expected to influence obesity. Literature suggested a reciprocal relationship between childhood obesity and behavioral measures such as personality/psychopathology and eating behavior. We had no expectations regarding the direction of the association with cognition and cortical thickness as causal associations have not been previously reported between them. The novelty of the present study lies in the usage of a large population-based dataset and the simultaneous consideration of a broad range of brain functioning and structure measures. No previous study has attempted to apply two causal methods to produce strong causal evidence for the relationships between childhood obesity and its neurobehavioral correlates.

## Methods

### Participants

We used data from the baseline, one-year and two-year follow-ups of the Adolescent Brain Cognitive Development (ABCD) study (Volkow et al., 2018). The study cohort includes 11875 children aged 9–10 years at baseline. After applying typical exclusion criteria to the available cases, the final sample comprised 7016 singletons and 2511 siblings (for exclusion criteria see Supplementary Table S1). Singletons were participants with unique family-IDs. Siblings were participants who had the same family-ID as one other participant within study waves and had information about their respective relationship available. Among singletons, the average age at baseline was 118.6 months (SD=7.3). 52.2% of singletons were boys, 47.7% were girls, and another gender was reported for 0.1%. Within the sibling subset, the average age at baseline was 119.8 months (SD=7.9). 51.3% of siblings were boys and 48.7% were girls (for demographics see Supplementary Table S2). Singleton data were used for cross-sectional analyses of associations between childhood obesity and neurobehavioral indicators as well as longitudinal analyses of those relationships. Sibling data were the basis for twin modeling.

The ABCD study dataset is a popular resource for scientist interested in behavioral neuroscience. Due to its open accessibility, some of the obesity–neurobehavior correlations being tested here have already been replicated within the dataset (Adise et al., 2021; Bohon & Welch, 2021; Dennis et al., 2022; Gray et al., 2020; Hall et al., 2021; Laurent et al., 2020; Ronan et al., 2020).

### Measures

#### Obesity

The percentage of the sex- and age-specific 95^th^ BMI percentile was calculated for each participant following guidelines for childhood obesity research (Freedman et al., 2017). Reference data was obtained from the US Centers for Disease Control and Prevention (http://www.cdc.gov/growthcharts/). All analyses used this continuous measure of obesity. Participants were considered having obesity when the percentage of the sex- and age-specific 95^th^ BMI percentile was 100 or greater. BMI is often used in research on obesity in adulthood, but the dependency of its distribution on sex and age during childhood renders it impractical for childhood obesity studies.

#### Neurobehavior

To group different neurobehavioral aspects, each variable of interest recorded in the ABCD study (called *feature*) was assigned to one of six neurobehavioral factors. These were personality/psychopathology, eating behavior, cognition, cortical thickness, cortical surface area, and cortical volume. The personality/psychopathology factor included 159 self- and parent-reported questionnaire items measuring impulsivity (Cyders et al., 2014), motivation (Carver & White, 1994), prosociality (Goodman, 1997), and psychopathology (Achenbach & Edelbrock, 1983). The eating behavior factor comprised two parent-reported eating items (Achenbach & Edelbrock, 1983). Thirteen cognitive test scores built the cognition factor (Lezak, 1976; Ratcliff, 1979; Wechsler, 1949; Weintraub et al., 2013). Cortical thickness, surface area, and volume included the respective measure in 68 regions of interest obtained from MRI scans. Brain morphology measures needed to successfully pass internal ABCD quality control. See Supplementary Table S3 and https://nda.nih.gov/general-query.html for detailed information on the measures.

### Statistical analyses

All analyses controlled for gender at birth, age, race, Hispanic ethnicity, handedness, puberty status and study site. Additionally, in analyses with brain morphology measures the individual mean or total of the respective measure was controlled for. This procedure was chosen to maintain consistency regarding controls for brain measures. Multiple control variables were correlated to childhood obesity (see Supplementary Figure S1). False discovery rate correction (Benjamini & Hochberg, 1995) was applied when screening for features within a neurobehavioral factor.

#### Correlational analyses within neurobehavioral factors

To validate previously found obesity–neurobehavior associations, cross-sectional correlations between childhood obesity and each neurobehavioral feature were calculated within the singleton subset. For an overview, the features within the personality/psychopathology factor (i.e., items of personality and psychopathology questionnaires) were summarized into their respective sum scales according to test manuals.

#### Poly-phenotype scores

Poly-phenotype scores (PPS; Vainik et al., 2018) were calculated to summarize the total association of each neurobehavioral factor with obesity. The PPS method is adapted from polygenic scores and helps to reduce measurement noise as well as multiple testing of redundant measures (e.g., Steinsbekk et al., 2016). PPSs are calculated by summing the products of each feature’s measure (i.e., personality/psychopathology/eating behavior item scores, cognitive test scores, and thickness in mm/surface area in mm^2^/volume in mm^3^ of cortical regions of interest) with its correlation with childhood obesity within a factor. To avoid overfitting, correlations were obtained from the singleton subset and applied to measures within the subset of siblings to generate six PPSs for each participant in the sibling subset. These PPSs informed preliminary correlation and twin modeling analyses. Ten-fold cross-validation within the singleton sample was later used to generate six corresponding PPSs in an independent sample for longitudinal analysis (see Supplementary Table S3).

#### Twin modeling

Direction of Causation (DoC) twin modeling was conducted to investigate the causal paths building the obesity–PPS associations. In DoC, the relationships of heritability variance components of two phenotypes are structurally modeled (Heath et al., 1993). This builds on the ACE heritability model which decomposes observed traits into additive genetic (A), common environment (C), and unique environment (E) variance components by comparing mono- and dizygotic twin pairs (non-twin siblings were treated equal to dizygotic twins; Neale & Cardon, 1992). If one trait has a causal influence another trait, then the A, C, and E variance components of this first trait should be proportionally represented in the variance of the second. In this way, DoC models can be specified for obesity causally influencing PPS, or PPS causally influencing obesity. A reciprocal model can be defined by allowing both causal paths. Removing any causal paths leads to the Cholesky model where observed correlations are driven by a third variable (Heath et al., 1993). For each obesity–PPS association, the best-fitting model was chosen by comparing model fit with regards to model complexity using Chi-squared and Akaike information criterion. For both, lower values indicate better model fit.

#### Longitudinal analyses

To validate DoC results, longitudinal analyses using cross-lagged panel models (CLPM) were performed. CLPMs estimate autoregressive paths which reflect stability of each measured trait over time (i.e., *a* paths) by structural modeling. Moreover, paths from previous measures of one trait to later measures of another trait are modeled. CLPMs refer to these as causal paths or *c* paths. However, they may not truly reflect causality: CLPMs test the ability of one trait’s past levels to predict later levels of another trait, but such relationships can be driven by unobserved third variables as well as causal influences (Usami et al., 2019).

Traditional CLPMs (Bentler & Speckart, 1981) were calculated for all six PPSs using baseline and two-year follow-up measures. Random-intercept CLPMs (Hamaker et al., 2015; Mund & Nestler, 2019) were applied where PPSs could be calculated for baseline, one-year and two-year follow-up (i.e., personality/psychopathology and eating behavior factors). The methodological advantage of Random-intercept CLPMs lays in the estimation of latent variables to account for stable interindividual trait differences (Hamaker et al., 2015). However, this method requires at least three observation waves. Due to varying availability of measures per study wave, PPSs for CLPM may not be fully congruent with PPSs for DoC (see Supplementary Table S3).

### Data and code availability

ABCD study data is available at https://abcdstudy.org/. A commented analysis script is available at osf.io/zgda4 for detailed data analysis procedure insight and replication. Analysis software information is noted in Supplementary Table S9.

## Results

At baseline, the average percentage of the sex- and age-specific 95th BMI percentile within the singleton subset was 85.0 (SD=17.9) with 17.7% having obesity. The average percentage of the sex- and age-specific 95th BMI percentile within the sibling subset was 82.0 (SD=16.4) with 13.3% having obesity. The results of the cross-sectional correlation analyses within six neurobehavioral factors, obesity–PPS correlation analyses, causal DoC twin modeling, and validating CLPM longitudinal analyses are reported in the following sections.

### Correlational analyses within neurobehavioral factors

#### Personality/psychopathology and eating behavior factors

In the subset of singletons, obesity was positively correlated to drive, reward responsiveness, social problems, somatic complaint, aggressive behavior, and being depressed. A negative correlation was found with sensation seeking. The strongest associations were between obesity and measures of psychopathology (Figure 1).

**Figure 1.**
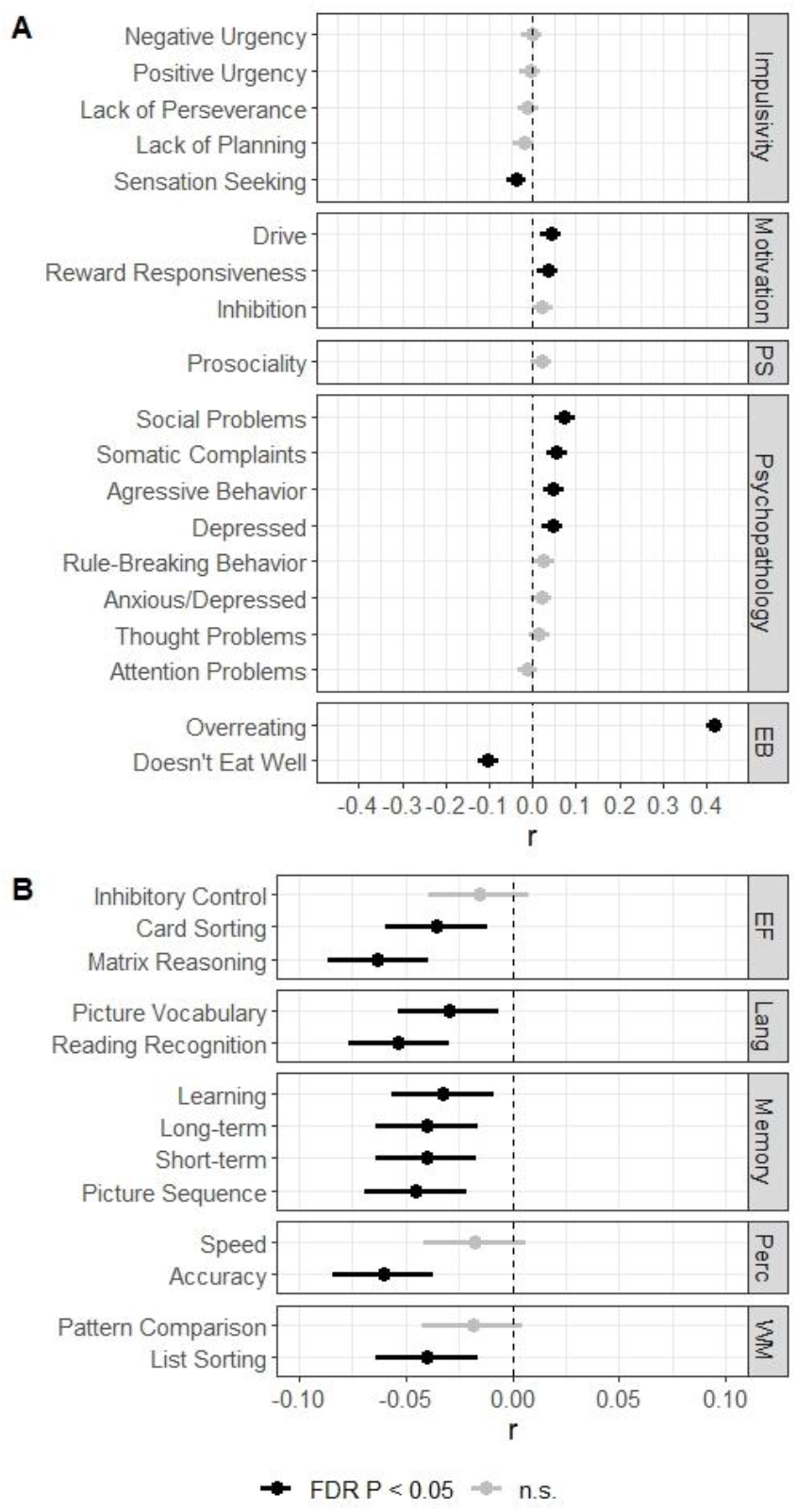
Associations between the percentage of the sex- and age-specific 95^th^ BMI percentile and personality/psychopathology scales, eating behavior items (A), as well as cognitive test scores (B) *Note*. Impulsivity, motivation and prosociality were self-reported. Psychopathology (including eating items) was parent-reported. Error bars represent 95% confidence intervals. EB, eating behavior; EF, executive functioning; FDR, false discovery rate; Lang, Language; Perc, perception; PS, prosociality; WM, working memory.

Correlation analyses were conducted between childhood obesity and the eating-specific items “overeating” and “doesn’t eat well” of the Child Behavior Checklist which formed the eating behavior factor. Both eating-related and parent-report items had considerable correlations with obesity. A stronger positive correlation was found with “overeating” and a weaker negative correlation with “doesn’t eat well” (Figure 1). The feature “doesn’t eat well” was also represented in the somatic complaints score within the personality/psychopathology factor for correlational analysis. In following analyses, eating-specific items were excluded from the personality/psychopathology factor.

#### Cognition

Cognitive test performance showed mainly negative correlations with obesity in the singleton subset. Exceptions were inhibitory control, perception speed, and pattern comparison where no significant associations were found. The obesity–cognition association was strongest for the Wechsler matrix reasoning task (Figure 1).

#### Brain morphology factors

Analyses of singletons revealed positive associations of posterior cortical thickness in the occipital and parietal lobe. Moreover, a positive correlation was also found with the precentral region of interest within the frontal lobe. Negative associations between cortical thickness and obesity were found for most regions of interest within the frontal lobe as well as superior temporal and temporal pole regions within the temporal lobe. Higher correlations for the right compared to the left hemisphere were discovered in some frontal and parietal brain regions (Figure 2).

**Figure 2.**
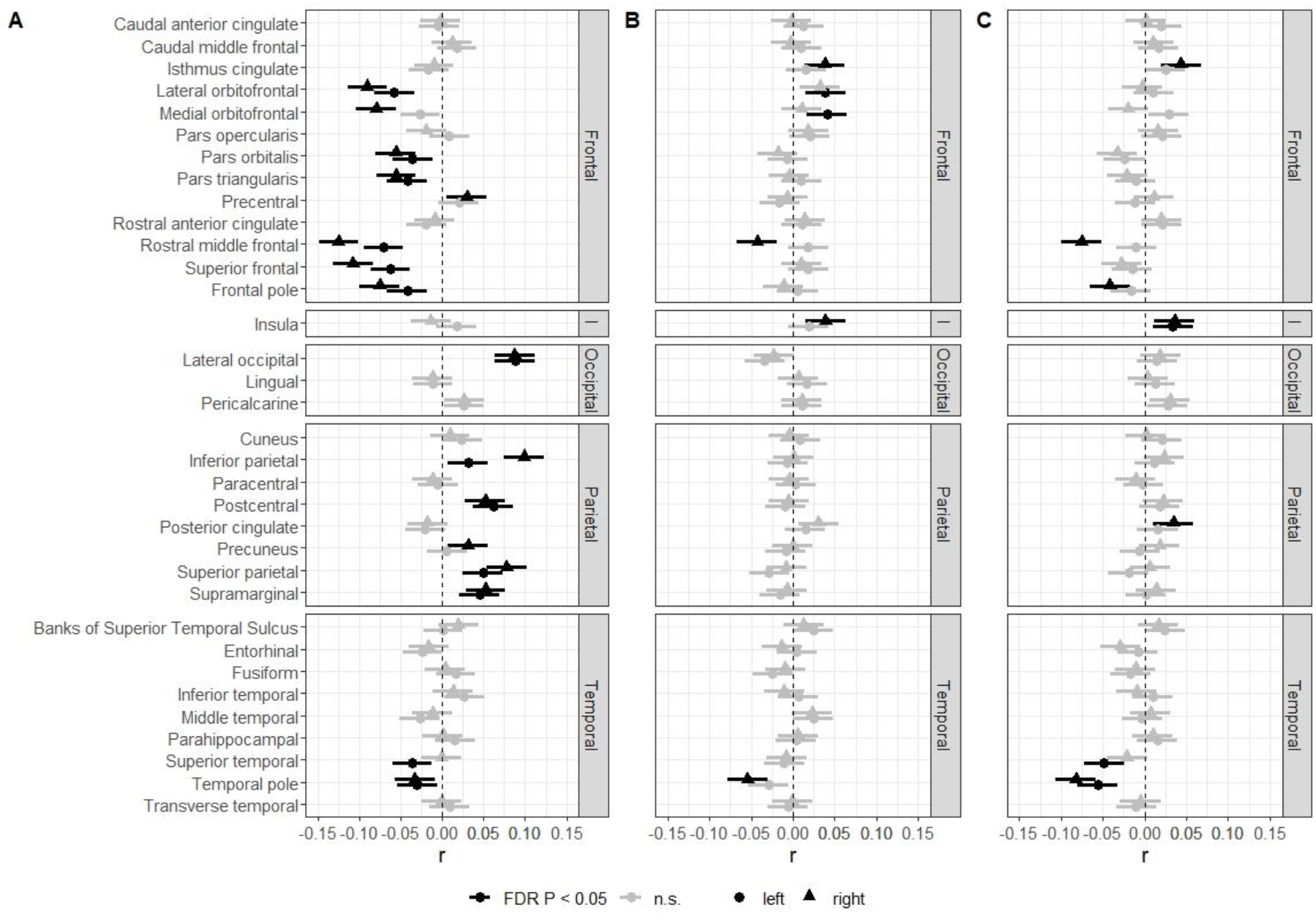
Associations between the percentage of the sex- and age-specific 95^th^ BMI percentile and region of interest cortical thickness (A), cortical surface area (B) as well as cortical volume (C) *Note*. Error bars represent 95% confidence intervals. FDR, false discovery rate; I, insula.

Fewer meaningful associations were found for cortical surface area and volume. Surface area–obesity associations were present in the isthmus cingulate, lateral orbitofrontal, medial orbitofrontal, rostral middle frontal, insula, and temporal pole regions of interest (Figure 2) while volume–obesity associations were found with the isthmus cingulate, rostral middle frontal, frontal pole, insula, posterior cingulate, superior temporal, and temporal pole regions of interest (Figure 2). See Supplementary Figure S2 for mapped visualization of obesity– morphology correlations.

### Poly-phenotype scores

Obesity–PPS correlations in the twin sample were r=.09, p<.001, n=2469 for personality/psychopathology, r=.36, p<.001, n=2475 for eating behavior, r=.04, p=.02, n=2296 for cognition, r=.14, p<.001, n=2391 for cortical thickness, r=.06, p<.01, n=2391 for cortical surface area, and r=.07, p<.001, n=2391 for cortical volume (for full correlation matrix see Supplementary Table S4).

### Direction of Causation

The heritability of obesity was 89.5% with the remaining 10.5% being due to unique experiences of a co-sibling. PPS heritability varied between 6.6% (eating behavior) and 57.5% (cortical thickness, see Supplementary Tables S5–6).

Based on model fit, data were consistent with obesity having unidirectional causal effects on cognition (β=.05, CI=[.00, .09], n_Pairs_=1056), cortical thickness (β=.15, CI=[.11, .20], n_Pairs_=1138), and cortical surface area (β=.07, CI=[.03, .11], n_Pairs_=1138). Contrarily, the personality/psychopathology (β=.09, CI=[.05, .12], n_Pairs_=1213) and cortical volume (β=.07, CI=[.04, .11], n_Pairs_=1138) PPS appeared to have unidirectional causal effects on obesity. The reciprocal causation model had the best fit for eating behavior (β_Obesity-to-PPS_ =.26, CI_Obesity-to-PPS_ =[.20, .32], β_PPS-to-Obesity_ =.16, CI_PPS-to-Obesity_ =[.11, .20], n_Pairs_=1218). Detailed DoC modeling statistics are reported in Supplementary Table S7.

### Cross-lagged panel models

Using the preferred random-intercept CLPM, meaningful *c* paths were found from obesity to eating behavior (β=.11, CI=[.03, .19], n=3497) and from personality/psychopathology to obesity (β=.04, CI=[.02, .07], n=3482). Traditional CLPMs found *c* paths from obesity to all neurobehavioral factors: personality/psychopathology (β=.07, CI=[.04, .10], n=3416), eating behavior (β=.22, CI=[.18, .26], n=3442), cognition (β=-.04, CI=[-.08, -.01], n=2952), cortical thickness (β=.08, CI=[.05, .11], n=3215), cortical surface area (β=.02, CI=[.01, .03], n=3215), and cortical volume (β=.02, CI=[.00, .03], n=3215). *C* paths also emerged from personality/psychopathology (β=.03, CI=[.01, .05], n=3416) and eating behavior (β=.05, CI=[.02, .08], n=3442) to obesity. Complete CLPM path estimates are shown in Supplementary Table S8.

The directional relationships found in DoC twin modeling were fully replicated except for the causal effects of cortical volume and eating behavior (only traditional CLPM) on childhood obesity. Additional longitudinal paths emerged from childhood obesity to personality/psychopathology (only traditional CLPM) and cortical volume (Figure 3). The negative longitudinal childhood obesity–cognition effect implies that higher obesity at age nine to ten is associated with higher cognitive test scores at age eleven to twelve.

**Figure 3.**
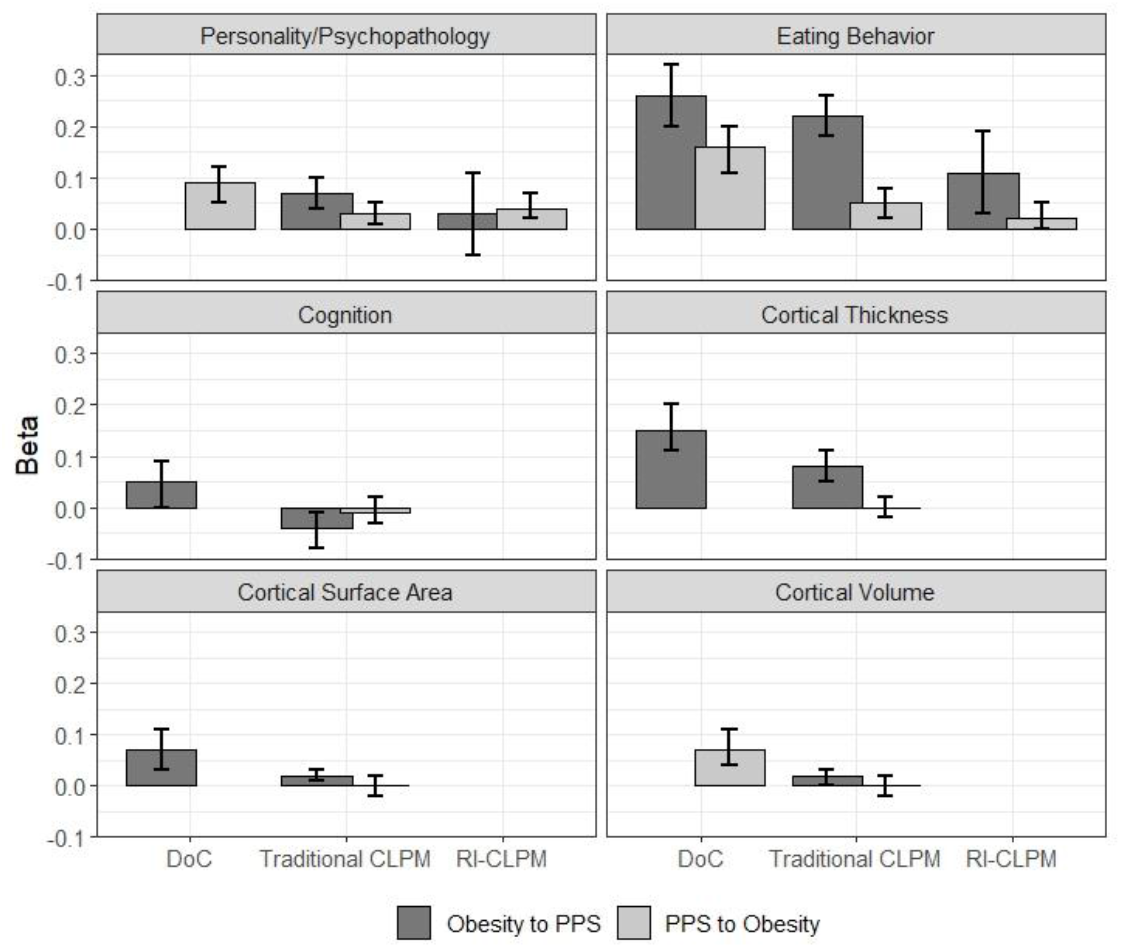
Causal and c path estimates for Direction of Causation (DoC), traditional cross-lagged panel (CLPM) and random-intercept cross-lagged panel (RI-CLPM) models *Note*. Only best fitting model plotted for Direction of Causation modeling. Error bars represent 95% confidence intervals. PPS, poly-phenotype score.

## Discussion

The purpose of the present study was to apply multiple causal inference methods to advance the understanding of associations between childhood obesity and neurobehavioral factors. Replicating previous cross-sectional findings, childhood obesity at age nine to ten was found to be associated to measures of personality/psychopathology, eating behavior, cognition, cortical thickness, cortical surface area, and cortical volume. The correlations between obesity and aggregated PPSs were similar to meta-analytic estimates (Kininmonth et al., 2021; Sutaria et al., 2019) and previous studies using the PPS method (Vainik et al., 2018) with the exception of a lower obesity–cognition estimate than previously reported (Pearce et al., 2018).

Regarding the direction of these relationships, the data supported causal influences of obesity on eating behavior, cognition, cortical thickness, and cortical surface area across analysis methods. Personality/psychopathology showed consistent causal and longitudinal effects on obesity. A reciprocal relationship may drive the associations between obesity and eating behavior. However, a random-intercept CLPM found only longitudinal effects from obesity to eating behavior. Both reciprocal obesity–eating behavior models (i.e., DoC and traditional CLPM) implied the latter to be the stronger path, too. For the association between obesity and personality/psychopathology, only the methodologically weaker traditional CLPM suggested a reciprocal relationship. Conflicting findings leave the relationship between obesity and cortical volume unclear. Together, the results of this multi-method study in a large sample highlight the importance of obesity as a driving force behind individual differences in brain functioning and morphology during childhood.

Results are in line with previous longitudinal evidence for a causal influence of personality and psychopathology measures on childhood obesity (Anzman-Frasca et al., 2015; Blaine, 2008; Camfferman et al., 2016; Christensen et al., 2019; De Decker et al., 2017; Derks et al., 2019; Korczak et al., 2013) as well as studies suggesting obesity and eating behavior to have a bidirectional relationship (Antoniou et al., 2016; Herle et al., 2021; Reed et al., 2017). Some studies suggested that childhood obesity may also influence psychopathology (Bradley et al., 2008; Hübel et al., 2019; Korczak et al., 2013). Evidence for this direction was only generated by traditional CLPM modeling. A longitudinal influence of cortical surface area on body mass (Park et al., 2020) was not replicated. Novel relationships emerged between childhood obesity and cognition as well as cortical thickness.

In recent studies, deficits in cognitive abilities of obese adults have been attributed to an obesity-associated inflammation in the brain (Spyridaki et al., 2016). Even though less pronounced, this effect can be partly observed in children, too (Adelantado-Renau et al., 2019). The beginning inflammation in the brains of obese children may explain the small effect from obesity to cognition in children aged 9–10. Brain inflammation may cause accelerated brain aging leading to cortical thinning. Albeit the effect being most pronounced in adulthood, similar degeneration processes may drive the effect of obesity on cortical thickness and surface area in the present study (Ronan et al., 2016). Reduced cortical thickness or surface area in frontal brain areas associated with executive functioning could mediate effects of obesity on performance in several cognitive tests.

However, a longitudinal correlation from increased obesity to *better* cognitive functioning was observed using a traditional CLPM. The previously negative association disappeared at age 11–12. Changes in the association between obesity and intelligence across childhood have been previously described, implying that negative effects of obesity can potentially be compensated in certain developmental periods (Wraw et al., 2018). However, the positive effect could not be replicated across methods. Further research is necessary to map this effect.

On the other hand, potential self- and parent-reported contributors to childhood obesity (i.e., eating behavior, psychopathology, impulsivity, and motivation) were identified. Of them, strongest effects could be summarized as increased behavioral approach motivation (drive, reward responsiveness), which may lead to unhealthy food choices that maximize short term satisfaction (Zhou et al., 2019). Besides food quality, food quantity could also be affected. The reduced ability to stop eating when hunger is satisfied may lead to overeating (van den Berg et al., 2011). The other domain seems to be stress-related psychopathology. Children with increased mental health problems may attempt to regulate adverse emotions by stress eating, which may contribute to weight gain (Hill et al., 2018). Current evidence favors focusing on interventions that aid reducing stress and/or try to manage increased sensation seeking.

Although the results suggest that eating behavior could contribute to obesity, its effect seemed trivial compared to the reverse effect of obesity on eating behavior. While rarely considered, such association is consistent with obesity increasing fat-free mass which in turn increases food intake, which then maintains obesity (Blundell, 2018). Fat-free mass and food intake have been linked in adolescents (Vainik et al., 2016), and genetic obesity predicts future childhood eating behaviors (Herle et al., 2021). Therefore, studies intervening on eating behaviors should also consider reverse causality.

The results are limited using the percentage of the sex- and age-specific 95th BMI percentile as a proxy for obesity and the assumption of obesity being continuous while most participants did not have obesity. Relative fat mass was explored as an alternative approach to obesity (Arumäe et al., 2022). However, correlations were highly similar and relative fat mass was dropped. The PPS method limits the results by ignoring causal relationships of individual features within a factor, but this was offset by gain in statistical power. DoC results are influenced by measurement reliability in a way that DoC tends to model the more reliable variable to have causal effects on the less reliable variable. Reliability could be lower for behavioral (Nakamura et al., 2009) and cognitive factors (Taylor et al., 2020) which could contribute to the finding that obesity had causal effects on eating behavior and cognition. Personality/psychopathology and cognitive test results can be compromised by intrapersonal components like response bias or motivation which are irrelevant for the directly observed measures of childhood obesity and brain morphology. At the same time, previous analyses showed that explicitly modeling reliability did not change DoC conclusions (Arumäe et al., 2021). Future studies using different causal inference methods should build complementary evidence (Arumäe et al., 2021; Munafò & Davey Smith, 2018).

In summary, the present study provides evidence that childhood obesity may influence rather than be influenced by the various aspects of brain functioning and morphology that have been found to differ between children with and without obesity. Therefore, obesity may play an important role in brain development during childhood. The present findings are hoped to inspire further investigation of the underlying mechanisms behind these associations and development of more effective obesity interventions.

## Supporting information

commented analysis script

## Data Availability

All data produced are available online at https://abcdstudy.org/scientists/data-sharing/ . Analysis script is part of the supplementary material.

## References

Achenbach, T. M., & Edelbrock, C. S. (1983). Manual for the Child Behavior Checklist and Revised Child Behavior Profile. University Associates in Psychiatry.

Adelantado-Renau, M., Esteban-Cornejo, I., Rodriguez-Ayllon, M., Cadenas-Sanchez, C., Gil-Cosano, J. J., Mora-Gonzalez, J., Solis-Urra, P., Verdejo-Román, J., Aguilera, C. M., Escolano-Margarit, M. V., Verdejo-Garcia, A., Catena, A., Moliner-Urdiales, D., & Ortega, F. B. (2019). Inflammatory biomarkers and brain health indicators in children with overweight and obesity: The ActiveBrains project. Brain, Behavior, and Immunity, 81, 588–597. https://doi.org/10.1016/j.bbi.2019.07.020

Adise, S., Allgaier, N., Laurent, J., Hahn, S., Chaarani, B., Owens, M., Yuan, D., Nyugen, P., Mackey, S., Potter, A., & Garavan, H. P. (2021). Multimodal brain predictors of current weight and weight gain in children enrolled in the ABCD study ®. Developmental Cognitive Neuroscience,49(100948). https://doi.org/10.1016/j.dcn.2021.100948

Alosco, M. L., Stanek, K. M., Galioto, R., Korgaonkar, M. S., Grieve, S. M., Brickman, A. M., Spitznagel, M. B., & Gunstad, J. (2014). Body mass index and brain structure in healthy children and adolescents. International Journal of Neuroscience, 124(1), 49–55. https://doi.org/10.3109/00207454.2013.817408

Antoniou, E. E., Roefs, A., Kremers, S. P. J., Jansen, A., Gubbels, J. S., Sleddens, E. F. C., & Thijs, C. (2016). Picky eating and child weight status development: a longitudinal study. Journal of Human Nutrition and Dietetics, 29(3), 298–307. https://doi.org/10.1111/jhn.12322

Anzman-Frasca, S., Francis, L. A., & Birch, L. L. (2015). Inhibitory control is associated with psychosocial, cognitive, and weight outcomes in a longitudinal sample of girls. Translational Issues in Psychological Science, 1(3), 203–216. https://doi.org/10.1037/tps0000028

Arumäe, K., Briley, D., Colodro-Conde, L., Mortensen, E. L., Jang, K., Ando, J., Kandler, C., Sørensen, T. I. A., Dagher, A., Mõttus, R., & Vainik, U. (2021). Two genetic analyses to elucidate causality between body mass index and personality. International Journal of Obesity (2005), 45(10), 2244–2251. https://doi.org/10.1038/s41366-021-00885-4

Arumäe, K., Mõttus, R., & Vainik, U. (2022). Beyond BMI: Personality traits’ associations with adiposity and metabolic rate. Physiology & Behavior, 246, 113703. https://doi.org/10.1016/j.physbeh.2022.113703

Belsky, D. W., Caspi, A., Goldman-Mellor, S., Meier, M. H., Ramrakha, S., Poulton, R., & Moffitt, T. E. (2013). Is obesity associated with a decline in intelligence quotient during the first half of the life course? American Journal of Epidemiology, 178(9), 1461–1468. https://doi.org/10.1093/aje/kwt135

Benjamini, Y., & Hochberg, Y. (1995). Controlling the False Discovery Rate: A Practical and Powerful Approach to Multiple Testing. Journal of the Royal Statistical Society: Series B (Methodological), 57(1), 289–300. https://doi.org/10.1111/j.2517-6161.1995.tb02031.x

Bentler, P. M., & Speckart, G. (1981). Attitudes “cause” behaviors: A structural equation analysis. Journal of Personality and Social Psychology, 40(2), 226–238. https://doi.org/10.1037/0022-3514.40.2.226

Blaine, B. (2008). Does Depression Cause Obesity? Journal of Health Psychology, 13(8), 1190–1197. https://doi.org/10.1177/1359105308095977

Blundell, J. E. (2018). Behaviour, energy balance, obesity and capitalism. European Journal of Clinical Nutrition, 72(9), 1305–1309. https://doi.org/10.1038/s41430-018-0231-x

Blundell, J. E., & Finlayson, G. (2004). Is susceptibility to weight gain characterized by homeostatic or hedonic risk factors for overconsumption? Physiology & Behavior, 82(1), 21–25. https://doi.org/10.1016/j.physbeh.2004.04.021

Bohon, C., & Welch, H. (2021). Quadratic relations of BMI with depression and brain volume in children: Analysis of data from the ABCD study. Journal of Psychiatric Research, 136, 421–427. https://doi.org/10.1016/j.jpsychires.2021.02.038

Bradley, R. H., Houts, R., Nader, P. R., O’Brien, M., Belsky, J., & Crosnoe, R. (2008). The Relationship between Body Mass Index and Behavior in Children. The Journal of Pediatrics, 153(5), 629-634.e3. https://doi.org/10.1016/j.jpeds.2008.05.026

Camfferman, R., Jansen, P. W., Rippe, R. C. A., Mesman, J., Derks, I. P. M., Tiemeier, H., Jaddoe, V., & van der Veek, S. M. C. (2016). The association between overweight and internalizing and externalizing behavior in early childhood. Social Science & Medicine (1982), 168, 35–42. https://doi.org/10.1016/j.socscimed.2016.09.001

Carver, C. S., & White, T. L. (1994). Behavioral inhibition, behavioral activation, and affective responses to impending reward and punishment: The BIS/BAS Scales. Journal of Personality and Social Psychology, 67(2), 319–333.

Christensen, K. G., Nielsen, S. G., Olsen, N. J., Dalgård, C., Heitmann, B. L., & Larsen, S. C. (2019). Child behaviour and subsequent changes in body weight, composition and shape. PloS One, 14(12), e0226003. https://doi.org/10.1371/journal.pone.0226003

Cyders, M. A., Littlefield, A. K., Coffey, S., & Karyadi, K. A. (2014). Examination of a short English version of the UPPS-P Impulsive Behavior Scale. Addictive Behaviors, 39(9), 1372–1376. https://doi.org/10.1016/j.addbeh.2014.02.013

De Decker, A., De Clercq, B., Verbeken, S., Wells, J. C. K., Braet, C., Michels, N., De Henauw, S., & Sioen, I. (2017). Fat and lean tissue accretion in relation to reward motivation in children. Appetite, 108, 317–325. https://doi.org/10.1016/j.appet.2016.10.017

Dennis, E., Manza, P., & Volkow, N. D. (2022). Socioeconomic status, BMI, and brain development in children. Translational Psychiatry, 12(1), 33. https://doi.org/10.1038/s41398-022-01779-3

Derks, I. P. M., Bolhuis, K., Yalcin, Z., Gaillard, R., Hillegers, M. H. J., Larsson, H., Lundström, S., Lichtenstein, P., van Beijsterveldt, C. E. M., Bartels, M., Boomsma, D. I., Tiemeier, H., & Jansen, P. W. (2019). Testing Bidirectional Associations Between Childhood Aggression and BMI: Results from Three Cohorts. Obesity, oby.22419. https://doi.org/10.1002/oby.22419

Freedman, D. S., Butte, N. F., Taveras, E. M., Lundeen, E. A., Blanck, H. M., Goodman, A. B., & Ogden, C. L. (2017). BMI z -Scores are a poor indicator of adiposity among 2-to 19-year-olds with very high BMIs, NHANES 1999-2000 to 2013-2014. Obesity, 25(4), 739–746. https://doi.org/10.1002/oby.21782

Goodman, R. (1997). The Strengths and Difficulties Questionnaire: A Research Note. Journal of Child Psychology and Psychiatry, 38(5), 581–586. https://doi.org/10.1111/j.1469-7610.1997.tb01545.x

Gray, J. C., Schvey, N. A., & Tanofsky-Kraff, M. (2020). Demographic, psychological, behavioral, and cognitive correlates of BMI in youth: Findings from the Adolescent Brain Cognitive Development (ABCD) study. Psychological Medicine, 50(9), 1539–1547. https://doi.org/10.1017/S0033291719001545

Hall, P. A., Best, J., Beaton, E. A., Sakib, M. N., & Danckert, J. (2021). Morphology of the Prefrontal Cortex Predicts Body Composition in Early Adolescence: Cognitive Mediators and Environmental Moderators in the ABCD Study. Social Cognitive and Affective Neuroscience. https://doi.org/10.1093/scan/nsab104

Hamaker, E. L., Kuiper, R. M., & Grasman, R. P. P. P. (2015). A critique of the cross-lagged panel model. Psychological Methods, 20(1), 102–116. https://doi.org/10.1037/a0038889

Hamilton, D., Dee, A., & Perry, I. J. (2018). The lifetime costs of overweight and obesity in childhood and adolescence: a systematic review. Obesity Reviews, 19(4), 452–463. https://doi.org/10.1111/obr.12649

Hashimoto, T., Takeuchi, H., Taki, Y., Yokota, S., Hashizume, H., Asano, K., Asano, M., Sassa, Y., Nouchi, R., & Kawashima, R. (2015). Increased posterior hippocampal volumes in children with lower increase in body mass index: a 3-year longitudinal MRI study. Developmental Neuroscience, 37(2), 153–160. https://doi.org/10.1159/000370064

Heath, A. C., Kessler, R. C., Neale, M. C., Hewitt, J. K., Eaves, L. J., & Kendler, K. S. (1993). Testing hypotheses about direction of causation using cross-sectional family data. Behavior Genetics, 23(1), 29–50. https://doi.org/10.1007/BF01067552

Herle, M., Abdulkadir, M., Hübel, C., Ferreira, D. S., Bryant-Waugh, R., Loos, R. J. F., Bulik, C. M., De Stavola, B., & Micali, N. (2021). The genomics of childhood eating behaviours. Nature Human Behaviour, 5(5), 625–630. https://doi.org/10.1038/s41562-020-01019-y

Hill, D. C., Moss, R. H., Sykes-Muskett, B., Conner, M., & O’Connor, D. B. (2018). Stress and eating behaviors in children and adolescents: Systematic review and meta-analysis. Appetite, 123, 14–22. https://doi.org/10.1016/j.appet.2017.11.109

Hübel, C., Gaspar, H. A., Coleman, J. R. I., Hanscombe, K. B., Purves, K., Prokopenko, I., Graff, M., Ngwa, J. S., Workalemahu, T., ADHD Working Group of the Psychiatric Genomics Consortium, Meta-Analyses of Glucose and Insulin-related traits consortium (MAGIC), Autism Working Group of the Psychiatric Genomics Consortium, Bipolar Disorder Working Group of the Psychiatric Genomics Consortium, Eating Disorders Working Group of the Psychiatric Genomics Consortium, Major Depressive Disorder Working Group of the Psychiatric Genomics Consortium, OCD & Tourette Syndrome Working Group of the Psychiatric Genomics Consortium, PTSD Working Group of the Psychiatric Genomics Consortium, Schizophrenia Working Group of the Psychiatric Genomics Consortium, Sex Differences Cross Disorder Working Group of the Psychiatric Genomics Consortium, … Breen, G. (2019). Genetic correlations of psychiatric traits with body composition and glycemic traits are sex-and age-dependent. Nature Communications, 10(1), 5765. https://doi.org/10.1038/s41467-019-13544-0

Kennedy, J. T., Collins, P. F., & Luciana, M. (2016). Higher Adolescent Body Mass Index Is Associated with Lower Regional Gray and White Matter Volumes and Lower Levels of Positive Emotionality. Frontiers in Neuroscience, 10. https://doi.org/10.3389/fnins.2016.00413

Kim, M. S., Luo, S., Azad, A., Campbell, C. E., Felix, K., Cabeen, R. P., Belcher, B. R., Kim, R., Serrano-Gonzalez, M., & Herting, M. M. (2020). Prefrontal Cortex and Amygdala Subregion Morphology Are Associated With Obesity and Dietary Self-control in Children and Adolescents. Frontiers in Human Neuroscience, 14, 563415. https://doi.org/10.3389/fnhum.2020.563415

Kininmonth, A., Smith, A., Carnell, S., Steinsbekk, S., Fildes, A., & Llewellyn, C. (2021). The association between childhood adiposity and appetite assessed using the Child Eating Behavior Questionnaire and Baby Eating Behavior Questionnaire: A systematic review and meta-analysis. Obesity Reviews,22(5). https://doi.org/10.1111/obr.13169

Korczak, D. J., Lipman, E., Morrison, K., & Szatmari, P. (2013). Are children and adolescents with psychiatric illness at risk for increased future body weight? A systematic review. Developmental Medicine & Child Neurology, 55(11), 980–987. https://doi.org/10.1111/dmcn.12168

Laurent, J. S., Watts, R., Adise, S., Allgaier, N., Chaarani, B., Garavan, H., Potter, A., & Mackey, S. (2020). Associations Among Body Mass Index, Cortical Thickness, and Executive Function in Children. JAMA Pediatrics, 174(2), 170–177. https://doi.org/10.1001/jamapediatrics.2019.4708

Lezak, M. D. (1976). Neuropsychological assessment. Oxford University Press.

Llewellyn, A., Simmonds, M., Owen, C. G., & Woolacott, N. (2016). Childhood obesity as a predictor of morbidity in adulthood: a systematic review and meta-analysis. Obesity Reviews, 17(1), 56–67. https://doi.org/10.1111/obr.12316

Lugo-Candelas, C., Pang, Y., Lee, S., Cha, J., Hong, S., Ranzenhofer, L., Korn, R., Davis, H., McInerny, H., Schebendach, J., Chung, W. K., Leibel, R. L., Walsh, B. T., Posner, J., Rosenbaum, M., & Mayer, L. (2020). Differences in brain structure and function in children with the FTO obesity-risk allele. Obesity Science & Practice, 6(4), 409–424. https://doi.org/10.1002/osp4.417

Munafò, M. R., & Davey Smith, G. (2018). Robust research needs many lines of evidence. Nature, 553(7689), 399–401. https://doi.org/10.1038/d41586-018-01023-3

Mund, M., & Nestler, S. (2019). Beyond the Cross-Lagged Panel Model: Next-generation statistical tools for analyzing interdependencies across the life course. Advances in Life Course Research, 41, 100249. https://doi.org/10.1016/j.alcr.2018.10.002

Nakamura, B. J., Ebesutani, C., Bernstein, A., & Chorpita, B. F. (2009). A Psychometric Analysis of the Child Behavior Checklist DSM-Oriented Scales. Journal of Psychopathology and Behavioral Assessment, 31(3), 178–189. https://doi.org/10.1007/s10862-008-9119-8

Neale, M. C., & Cardon, L. R. (1992). Methodology for genetic studies of twins and families. Kluwer Academic Publishers.

Park, B.-Y., Chung, C.-S., Lee, M. J., & Park, H. (2020). Accurate neuroimaging biomarkers to predict body mass index in adolescents: a longitudinal study. Brain Imaging and Behavior, 14(5), 1682–1695. https://doi.org/10.1007/s11682-019-00101-y

Pearce, A. L., Leonhardt, C. A., & Vaidya, C. J. (2018). Executive and Reward-Related Function in Pediatric Obesity: A Meta-Analysis. Childhood Obesity, 14(5), 265–279. https://doi.org/10.1089/chi.2017.0351

Prats-Soteras, X., Jurado, M. A., Ottino-González, J., García-García, I., Segura, B., Caldú, X., Sánchez-Garre, C., Miró, N., Tor, C., Sender-Palacios, M., & Garolera, M. (2020). Inflammatory agents partially explain associations between cortical thickness, surface area, and body mass in adolescents and young adulthood. International Journal of Obesity, 44(7), 1487–1496. https://doi.org/10.1038/s41366-020-0582-y

Rajan, L., McKay, C. C., Santos Malavé, G., Pearce, A. L., Cherry, J. B. C., Mackey, E., Nadler, E. P., & Vaidya, C. J. (2021). Effects of severe obesity and sleeve gastrectomy on cortical thickness in adolescents. Obesity, 29(9), 1516–1525. https://doi.org/10.1002/oby.23206

Ratcliff, G. (1979). Spatial thought, mental rotation and the right cerebral hemisphere. Neuropsychologia, 17(1), 49–54. https://doi.org/10.1016/0028-3932(79)90021-6

Reed, Z. E., Micali, N., Bulik, C. M., Davey Smith, G., & Wade, K. H. (2017). Assessing the causal role of adiposity on disordered eating in childhood, adolescence, and adulthood: a Mendelian randomization analysis. The American Journal of Clinical Nutrition, 106(3), 764–772. https://doi.org/10.3945/ajcn.117.154104

Ronan, L., Alexander-Bloch, A. F., Wagstyl, K., Farooqi, S., Brayne, C., Tyler, L. K., & Fletcher, P. C. (2016). Obesity associated with increased brain age from midlife. Neurobiology of Aging, 47, 63–70. https://doi.org/10.1016/j.neurobiolaging.2016.07.010

Ronan, L., Alexander-Bloch, A., & Fletcher, P. C. (2020). Childhood Obesity, Cortical Structure, and Executive Function in Healthy Children. Cerebral Cortex (New York, N.Y. : 1991), 30(4), 2519–2528. https://doi.org/10.1093/cercor/bhz257

Ross, N., Yau, P. L., & Convit, A. (2015). Obesity, fitness, and brain integrity in adolescence. Appetite, 93, 44–50. https://doi.org/10.1016/j.appet.2015.03.033

Sahoo, K., Sahoo, B., Choudhury, A. K., Sofi, N. Y., Kumar, R., & Bhadoria, A. S. (2015). Childhood obesity: causes and consequences. Journal of Family Medicine and Primary Care, 4(2), 187–192. https://doi.org/10.4103/2249-4863.154628

Saute, R. L., Soder, R. B., Alves Filho, J. O., Baldisserotto, M., & Franco, A. R. (2018). Increased brain cortical thickness associated with visceral fat in adolescents. Pediatric Obesity, 13(1), 74–77. https://doi.org/10.1111/ijpo.12190

Shriver, L. H., Dollar, J. M., Lawless, M., Calkins, S. D., Keane, S. P., Shanahan, L., & Wideman, L. (2019). Longitudinal Associations between Emotion Regulation and Adiposity in Late Adolescence: Indirect Effects through Eating Behaviors. Nutrients,11(3). https://doi.org/10.3390/nu11030517

Spyridaki, E. C., Avgoustinaki, P. D., & Margioris, A. N. (2016). Obesity, inflammation and cognition. Current Opinion in Behavioral Sciences, 9, 169–175. https://doi.org/10.1016/j.cobeha.2016.05.004

Steegers, C., Blok, E., Lamballais, S., Jaddoe, V., Bernardoni, F., Vernooij, M., van der Ende, J., Hillegers, M., Micali, N., Ehrlich, S., Jansen, P., Dieleman, G., & White, T. (2021). The association between body mass index and brain morphology in children: a population-based study. Brain Structure & Function, 226(3), 787–800. https://doi.org/10.1007/s00429-020-02209-0

Steinsbekk, S., Belsky, D., Guzey, I. C., Wardle, J., & Wichstrøm, L. (2016). Polygenic Risk, Appetite Traits, and Weight Gain in Middle Childhood. JAMA Pediatrics, 170(2), e154472. https://doi.org/10.1001/jamapediatrics.2015.4472

Sutaria, S., Devakumar, D., Yasuda, S. S., Das, S., & Saxena, S. (2019). Is obesity associated with depression in children? Systematic review and meta-analysis. Archives of Disease in Childhood, 104(1), 64–74. https://doi.org/10.1136/archdischild-2017-314608

Taylor, B. K., Frenzel, M. R., Eastman, J. A., Wiesman, A. I., Wang, Y.-P., Calhoun, V. D., Stephen, J. M., & Wilson, T. W. (2020). Reliability of the NIH toolbox cognitive battery in children and adolescents: a 3-year longitudinal examination. Psychological Medicine, 1–10. https://doi.org/10.1017/S0033291720003487

Thamotharan, S., Lange, K., Zale, E. L., Huffhines, L., & Fields, S. (2013). The role of impulsivity in pediatric obesity and weight status: A meta-analytic review. Clinical Psychology Review, 33(2), 253–262. https://doi.org/10.1016/j.cpr.2012.12.001

Thapaliya, G., Chen, L., Jansen, E., Smith, K. R., Sadler, J. R., Benson, L., Papantoni, A., & Carnell, S. (2021). Familial Obesity Risk and Current Excess Weight Influence Brain Structure in Adolescents. Obesity (Silver Spring, Md.), 29(1), 184–193. https://doi.org/10.1002/oby.23042

Usami, S., Murayama, K., & Hamaker, E. L. (2019). A unified framework of longitudinal models to examine reciprocal relations. Psychological Methods, 24(5), 637–657. https://doi.org/10.1037/met0000210

Vainik, U., Baker, T. E., Dadar, M., Zeighami, Y., Michaud, A., Zhang, Y., García Alanis, J. C., Misic, B., Collins, D. L., & Dagher, A. (2018). Neurobehavioral correlates of obesity are largely heritable. Proceedings of the National Academy of Sciences, 115(37), 9312–9317. https://doi.org/10.1073/pnas.1718206115

Vainik, U., Konstabel, K., Lätt, E., Mäestu, J., Purge, P., & Jürimäe, J. (2016). Diet misreporting can be corrected: confirmation of the association between energy intake and fat-free mass in adolescents. British Journal of Nutrition, 116(8), 1425–1436. https://doi.org/10.1017/S0007114516003317

van den Berg, L., Pieterse, K., Malik, J. A., Luman, M., Willems van Dijk, K., Oosterlaan, J., & Delemarre-van de Waal, H. A. (2011). Association between impulsivity, reward responsiveness and body mass index in children. International Journal of Obesity, 35(10), 1301–1307. https://doi.org/10.1038/ijo.2011.116

Volkow, N. D., Koob, G. F., Croyle, R. T., Bianchi, D. W., Gordon, J. A., Koroshetz, W. J., Pérez-Stable, E. J., Riley, W. T., Bloch, M. H., Conway, K., Deeds, B. G., Dowling, G. J., Grant, S., Howlett, K. D., Matochik, J. A., Morgan, G. D., Murray, M. M., Noronha, A., Spong, C. Y., … Weiss, S. R. B. (2018). The conception of the ABCD study: From substance use to a broad NIH collaboration. Developmental Cognitive Neuroscience, 32, 4–7. https://doi.org/10.1016/j.dcn.2017.10.002

Wang, Y., Beydoun, M. A., Min, J., Xue, H., Kaminsky, L. A., & Cheskin, L. J. (2020). Has the prevalence of overweight, obesity and central obesity levelled off in the United States? Trends, patterns, disparities, and future projections for the obesity epidemic. International Journal of Epidemiology, 49(3), 810–823. https://doi.org/10.1093/ije/dyz273

Wechsler, D. (1949). Wechsler Intelligence Scale for Children. Psychological Corporation.

Weintraub, S., Dikmen, S. S., Heaton, R. K., Tulsky, D. S., Zelazo, P. D., Bauer, P. J., Carlozzi, N. E., Slotkin, J., Blitz, D., Wallner-Allen, K., Fox, N. A., Beaumont, J. L., Mungas, D., Nowinski, C. J., Richler, J., Deocampo, J. A., Anderson, J. E., Manly, J. J., Borosh, B., … Gershon, R. C. (2013). Cognition assessment using the NIH Toolbox. Neurology, 80(11 Suppl 3), S54–64. https://doi.org/10.1212/WNL.0b013e3182872ded

Wraw, C., Deary, I. J., Der, G., & Gale, C. R. (2018). Maternal and offspring intelligence in relation to BMI across childhood and adolescence. International Journal of Obesity (2005), 42(9), 1610–1620. https://doi.org/10.1038/s41366-018-0009-1

Zhou, Z., SooHoo, M., Zhou, Q., Perez, M., & Liew, J. (2019). Temperament as Risk and Protective Factors in Obesogenic Eating: Relations Among Parent Temperament, Child Temperament, and Child Food Preference and Eating. The Journal of Genetic Psychology, 180(1), 75–79. https://doi.org/10.1080/00221325.2019.1575180

