## supplementary for "Triangulating Causality Between Childhood Obesity and Neurobehavior: Twin and Longitudinal Evidence"

#### Supplementary Information

**Table S1**

*Exclusion of cases (baseline wave)*

| | N | % |
|---|---|---|
| Total available at baseline | 11933 | 100 |
| Excluded of missing important values (sex, puberty, race, ethnicity, age, $\%BMI_{p95}$) | 1062 | 8.9 |
| Excluded for Medical Condition (diabetes, brain injury, cancer, cerebral palsy, epilepsy) | 412 | 3.5 |
| Excluded for BMI calculation error | 38 | 0.3 |
| Excluded for underweight BMI | 212 | 1.8 |
| Excluded for taking birth control | 2 | <0.1 |
| Excluded of inaccurate relatedness info | 680 | 5.7 |
| Total at baseline after exclusion | 9527 | 79.8 |

*Note*. $\%BMI_{p95,}$ percentage of the sex- and age-specific 95th BMI percentile.

**Table S2**

*Demographic information*

| | Singletons (N = 7016) | | Siblings (N = 2511) | |
|---|---|---|---|---|
| | M | SD | M | SD |
| Age (months) | 118.6 | 7.3 | 119.8 | 7.9 |
| %BMIp95 | 85.0 | 17.9 | 82.0 | 16.4 |
| | n | % | n | % |
| Obese BMI | 1239 | 17.7 | 334 | 13.3 |
| Sex at birth | | | | |
| *-Male* | 3665 | 52.2 | 1288 | 51.3 |
| *-Female* | 3349 | 47.7 | 1223 | 48.7 |
| *-Other* | 2 | 0.1 | 0 | - |
| Race | | | | |
| *-White* | 4449 | 63.4 | 1758 | 70.0 |
| *-Black/African American* | 1044 | 14.9 | 323 | 12.9 |
| *-Other/multiple* | 1523 | 21.7 | 430 | 17.1 |
| Hispanic ethnicity | 1589 | 22.6 | 348 | 13.9 |
| Handedness | | | | |
| *-Right* | 5603 | 79.9 | 1993 | 79.4 |
| *-Left* | 475 | 6.8 | 197 | 7.8 |
| *-Mixed* | 938 | 13.4 | 321 | 12.8 |
| College education of mother | 5813 | 82.9 | 2235 | 89.0 |
| Family income | | | | |
| *-Less than $5,000* | 228 | 3.2 | 62 | 2.5 |
| *-$5,000 through $11,999* | 244 | 3.5 | 69 | 2.7 |
| *-$12,000 through $15,999* | 168 | 2.4 | 32 | 1.3 |
| *-$16,000 through $24,999* | 318 | 4.5 | 94 | 3.7 |
| *-$25,000 through $34,999* | 412 | 5.9 | 103 | 4.1 |
| *-$35,000 through $49,999* | 574 | 8.2 | 181 | 7.2 |
| *-$50,000 through $74,999* | 876 | 12.5 | 341 | 13.6 |
| *$75,000 through $99,999* | 959 | 13.7 | 362 | 14.1 |
| *-$100,000 through $199,999* | 1928 | 27.5 | 798 | 31.8 |
| *-$200,000 and greater* | 735 | 10.5 | 324 | 12.9 |
| *-No information* | 574 | 8.2 | 145 | 5.8 |

| | | | | |
|---|---|---|---|---|
| Parent informant | | | | |
| *-Childs Biological Mother* | 6012 | 85.7 | 2191 | 87.3 |
| *-Childs Biological Father* | 684 | 9.7 | 225 | 9.0 |
| *-Adoptive Parent* | 173 | 2.5 | 49 | 2.0 |
| *-Childs Custodial Parent* | 64 | 0.9 | 17 | 0.7 |
| *-Other* | 83 | 1.2 | 29 | 1.2 |

*Note*. Children were considered having obesity when their percentage of the sex- and age-specific 95th BMI percentile was equal or greater 100. $\%BMI_{p95}$, percentage of the sex- and age-specific 95th BMI percentile.

**Table S3**

*Composition of poly-phenotype scores*

| | DoC | Random-Intercept CLPM | Traditional CLPM |
|---|---|---|---|
| Calculation method | Weights obtained from singletons, applied to siblings | Weights obtained from singletons (baseline) using 10-fold cross-validation, applied to singletons (all waves) | |
| Personality/psychopathology | CBCL (116 items),<br>SDQ-PSB (3 items),<br>UPPS-P (20 items),<br>BIS/BAS (20 items) | CBCL (116 items),<br>PSB (3 items) | CBCL (116 items),<br>PSB (3 items),<br>UPPS-P (20 items),<br>BIS/BAS (20 items) |
| Eating behavior | overeating,<br>doesn’t eat well | overeating,<br>doesn’t eat well | overeating,<br>doesn’t eat well |
| Cognition | NIH-TB (7 scores),<br>RAVLT (3 scores),<br>LMT (2 score),<br>WISC Matrix test (1 score) | - | NIH-TB (5 scores),<br>RAVLT (3 scores),<br>LMT (2 score),<br>Matrix test (1 score) |
| Brain morphology factors | 68 MRI regions of interest | - | 68 MRI regions of interest |

*Note.* For description of instruments see Data Dictionary at https://nda.nih.gov/general-query.html. BIS/BAS, Behavioral Inhibition System/Behavioral Activation System Scale (Carver & White, 1994); CBCL, Child Behavior Checklist (Achenbach & Edelbrock, 1983); CLPM, cross-lagged panel model; DoC, Direction of Causation; LMT, Little Man Task

(Ratcliff, 1979); NIH-TB, NIH Toolbox for the Assessment of Neurological and Behavioral Function domain Cognition (Dimensional Change Card Sort Test and List Sorting Working Memory Test unique to baseline wave; Weintraub et al., 2013); RAVLT, Rey Auditory Verbal Learning Test (Lezak, 1976); SDQ-PSB, Strength and Difficulties Questionnaire subscale Prosocial Behavior (Goodman, 1997); UPPS-P urgency-premeditation-perseverance-sensation seeking-positive urgency Impulsive Behaviors Scale (Cyders et al., 2014); WISC, Wechsler Intelligence Scale for Children (Wechsler, 1949).

**Table S4**

*Estimates (below diagonal) and sample sizes (above diagonal) for correlation analyses between percentage of the sex- and age-specific 95th BMI percentile (%$BMI_{p95}$) and neurobehavior poly-phenotype scores*

| Variable | 1 | 2 | 3 | 4 | 5 | 6 | 7 |
|---|---|---|---|---|---|---|---|
| 1. %$BMI_{p95}$ | – | 2469 | 2475 | 2296 | 2391 | 2391 | 2391 |
| 2. Personality/psychopathology | .09*** | – | 2469 | 2292 | 2386 | 2386 | 2386 |
| 3. Eating behavior | .36*** | .16*** | – | 2295 | 2390 | 2390 | 2390 |
| 4. Cognition | .04* | .11*** | .04 | – | 2219 | 2219 | 2219 |
| 5. Cortical thickness | .14*** | -.02 | .09*** | .00 | – | 2391 | 2391 |
| 6. Cortical surface area | .06** | .02 | .05** | .01 | .12*** | – | 2391 |
| 7. Cortical volume | .07*** | -.01 | .03 | -.01 | .21*** | .35*** | - |

*Note.* Weights were trained on singletons and applied to siblings using all available data per variable combination. Associations are shown for the sibling subset. All correlations controlled for family structure. * p<.05; ** p<.01; *** p<.001.

**Table S5**

*Sibling pair sample sizes for twin modelling (including Direction of Causation)*

| | $N_{MZ}$ | $N_{DZ}$ | $N_{total}$ |
|---|---|---|---|
| Personality/psychopathology | 259 | 954 | 1213 |
| Eating behavior | 259 | 959 | 1218 |
| Cognition | 228 | 828 | 1056 |
| Cortical thickness | 245 | 893 | 1138 |
| Cortical surface area | 245 | 893 | 1138 |
| Cortical volume | 245 | 893 | 1138 |

*Note.* MZ, monozygotic twin pairs; DZ, dizygotic twin or sibling pairs.

**Table S6**

*Cholesky ACE model estimates of sex- and age-specific 95th BMI percentile (%$BMI_{p95}$) and neurobehavior poly-phenotype scores*

| Variable | A (%) | C (%) | E (%) | $r_a$ | $r_e$ |
|---|---|---|---|---|---|
| %$BMI_{p95}$ | 89.5 | - | 10.5 | - | - |
| Personality/psychopathology | 48.6 | 10.6 | 40.8 | .11 | .15 |
| Eating behavior | 6.6 | - | 93.4 | >.99 | .48 |
| Cognition | 31.4 | - | 68.6 | .09 | -.01 |
| Cortical thickness | 57.5 | - | 42.5 | .22 | .02 |
| Cortical surface area | 36.6 | 9.6 | 53.8 | .11 | .04 |
| Cortical volume | 44.6 | 0.8 | 54.6 | .06 | .16 |

*Note.* The sibling subset was used for twin modeling. A, additive genetic; C, common environment; E, unique environment; $r_a$, genetic correlation with %$BMI_{p95}$; $r_e$, unique environmental correlation with %$BMI_{p95}$.

**Table S7**

*Comparison of Direction of Causation models*

| PPS | Model | AIC | $\chi^2$ | Model comparison ($\chi^2$-test) | | $\beta_{\text{Obesity-to-PPS}}$ [CI] | $\beta_{\text{PPS-to-Obesity}}$ [CI] |
|---|---|---|---|---|---|---|---|
| | | | | vs. Cholesky (p) | vs. Reciprocal (p) | | |
| Personality/ | Cholesky | 13068.58 | 46.77 | - | - | - | - |
| psychopathology | Reciprocal | 13067.94 | 48.14 | .242 | - | .01 [-.09, .11] | .08 [-.01, .17] |
| | Obesity-to-PPS | 13068.98 | 51.18 | .110 | .081 | .09 [.05, .13] | - |
| | PPS-to-Obesity | 13065.98 | 48.17 | .496 | .855 | - | .09 [.05, .12] |
| Eating behavior | Cholesky | 12862.60 | 77.99 | - | - | - | - |
| | Reciprocal | 12861.50 | 78.89 | .342 | - | .26 [.20, .32] | .16 [.11, .20] |
| | Obesity-to-PPS | 12894.79 | 114.17 | <.001 | <.001 | .40 [.36, .44] | - |
| | PPS-to-Obesity | 12917.35 | 136.74 | <.001 | <.001 | - | .31 [.28, .35] |
| Cognition | Cholesky | 12780.38 | 29.10 | - | - | - | - |
| | Reciprocal | 12778.38 | 29.10 | >.999 | - | .06 [-.02, .13] | -.01 [-.07, .05] |
| | Obesity-to-PPS | 12776.51 | 29.24 | .935 | .713 | .05 [.00, .09] | - |
| | PPS-to-Obesity | 12778.52 | 31.25 | .342 | .143 | - | .03 [-.01, .06] |
| Cortical thickness | Cholesky | 12873.78 | 24.75 | - | - | - | - |
| | Reciprocal | 12871.78 | 24.75 | >.999 | - | .20 [.10, .30] | -.04 [-.13, .04] |
| | Obesity-to-PPS | 12870.70 | 25.66 | .633 | .339 | .15 [.11, .20] | - |
| | PPS-to-Obesity | 12884.42 | 39.39 | <.001 | <.001 | - | .11 [.08, .15] |

| | | | | | | | |
|---|---|---|---|---|---|---|---|
| Cortical surface area | Cholesky | 12942.59 | 21.02 | - | - | - | - |
| | Reciprocal | 12940.94 | 21.36 | .559 | - | .06 [-.02, .15] | .01 [-.06, .08] |
| | Obesity-to-PPS | 12938.99 | 21.42 | .822 | .819 | .07 [.03, .11] | - |
| | PPS-to-Obesity | 12941.07 | 23.50 | .290 | .144 | - | .05 [.02, .09] |
| Cortical volume | Cholesky | 12966.38 | 41.20 | - | - | - | - |
| | Reciprocal | 12964.38 | 41.20 | .981 | - | .01 [-.08, .09] | .07 [.00, .14] |
| | Obesity-to-PPS | 12966.30 | 45.13 | .141 | .048 | .08 [.04, .12] | - |
| | PPS-to-Obesity | 12962.42 | 41.25 | .978 | .833 | - | .07 [.04, .11] |

*Note*. The sibling subset was used for twin modeling. Estimated parameters per model: Cholesky 11, Reciprocal 10, Obesity-to-PPS and PPS-to-Obesity 9. $\chi^2$, Chi-squared model fit; AIC, Akaike information criterion; CI, 95% confidence interval; PPS, poly-phenotype score.

**Table S8**

*Cross-lagged panel model (CLPM) path estimates*

| | | Autoregressive path (*a*) | | "Causal" path (*c*) | |
|---|---|---|---|---|---|
| | N | Obesity | PPS | Obesity-to-PPS | PPS-to-Obesity |
| *Random-intercept CLPM (three waves)* | | | | | |
| Personality/psychopathology | 3482 | .47 [.40, .54] *** | .13 [.07, .19] *** | .03 [-.05, .11] | .04 [.02, .07] ** |
| Eating behavior | 3497 | .46 [.39, .53] *** | .11 [.06, .17] *** | .11 [.03, .19] ** | .02 [.00,.05] |
| *Traditional CLPM (two waves)* | | | | | |
| Personality/psychopathology | 3416 | .82 [.80, .84] *** | .50 [.47, .53] *** | .07 [.04, .10] *** | .03 [.01, .05] ** |
| Eating behavior | 3442 | .80 [.78, .83] *** | .38 [.34, .43] *** | .22 [.18, .26] *** | .05 [.02, .08] ** |
| Cognition | 2952 | .83 [.81, .85] *** | .18 [.14 .22] *** | -.04 [-.08, -.01] * | -.01 [-.03,.02] |
| Cortical thickness | 3215 | .82 [.80, .84] *** | .66 [.63, .68] *** | .08 [.05, .11] *** | .00 [-.02,.02] |
| Cortical surface area | 3215 | .82 [.80, .84] *** | .91 [.90, .92] *** | .02 [.01, .03] ** | .00 [-.02, .02] |
| Cortical volume | 3215 | .82 [.80, .84] *** | .90 [.89, .91] *** | .02 [.00, .03] * | .00 [-.02, .02] |

*Note.* The singleton subset was used for longitudinal modeling. N, sample size; * p <.05; ** p<.01; *** p<.001.

**Table S9**

Main R (R Core Team, 2020) packages used

| Analysis | Package |
|---|---|
| Twin analysis including Direction of Causation | umx (Bates et al., 2019) |
| Cross-lagged panel modelling | lavaan (Rosseel, 2012) |
| Figures | ggplot2 (Wickham, 2016), ggseg (Mowinckel & Vidal-Piñeiro, 2019) |

**Figure S1**

*Standardized regression coefficients for the associations between the percentage of the sex- and age-specific 95$^{th}$ BMI percentile and control variables*

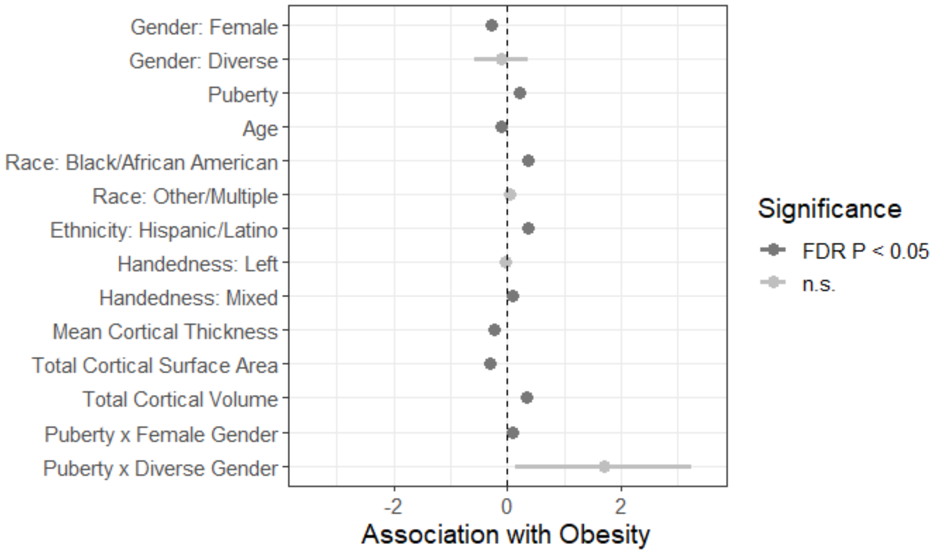


*Note*. Error bars represent 95% confidence intervals. FDR, false discovery rate.

**Figure S2**

*2D mapped associations between the percentage of the sex- and age-specific 95$^{th}$ BMI percentile and region of interest cortical thickness (A), cortical surface area (B) as well as cortical volume (C)*

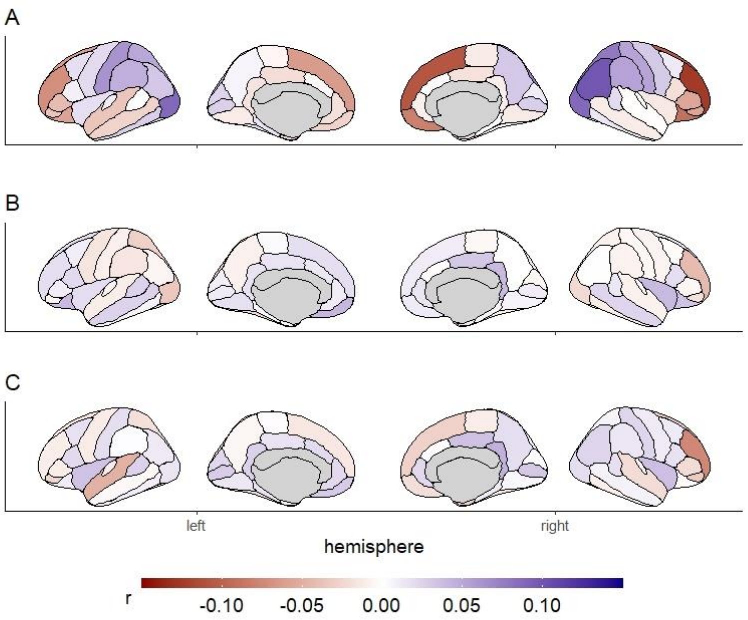

### References


Achenbach, T. M., & Edelbrock, C. S. (1983). *Manual for the Child Behavior Checklist and Revised Child Behavior Profile*. University Associates in Psychiatry.

Bates, T. C., Maes, H., & Neale, M. C. (2019). umx: Twin and Path-Based Structural Equation Modeling in R. *Twin Research and Human Genetics*, *22*(1), 27–41. https://doi.org/10.1017/thg.2019.2

Carver, C. S., & White, T. L. (1994). Behavioral inhibition, behavioral activation, and affective responses to impending reward and punishment: The BIS/BAS Scales. *Journal of Personality and Social Psychology*, *67*(2), 319–333.

Cyders, M. A., Littlefield, A. K., Coffey, S., & Karyadi, K. A. (2014). Examination of a short English version of the UPPS-P Impulsive Behavior Scale. *Addictive Behaviors*, *39*(9), 1372–1376. https://doi.org/10.1016/j.addbeh.2014.02.013

Goodman, R. (1997). The Strengths and Difficulties Questionnaire: A Research Note. *Journal of Child Psychology and Psychiatry, 38*(5), 581–586. https://doi.org/10.1111/j.1469-7610.1997.tb01545.x

Lezak, M. D. (1976). *Neuropsychological assessment*. Oxford University Press.

Mowinckel, A. M., & Vidal-Piñeiro, D. (2019). *Visualisation of Brain Statistics with R-packages ggseg and ggseg3d*. arXiv.

R Core Team. (2020). *R: A language and environment for statistical computing. R Foundation for Statistical Computing*.

Ratcliff, G. (1979). Spatial thought, mental rotation and the right cerebral hemisphere. *Neuropsychologia*, *17*(1), 49–54. https://doi.org/10.1016/0028-3932(79)90021-6

Rosseel, Y. (2012). lavaan : An R Package for Structural Equation Modeling. *Journal of Statistical Software*, *48*(2). https://doi.org/10.18637/jss.v048.i02

Wechsler, D. (1949). *Wechsler Intelligence Scale for Children*. Psychological Corporation.

Weintraub, S., Dikmen, S. S., Heaton, R. K., Tulsky, D. S., Zelazo, P. D., Bauer, P. J., Carlozzi, N. E., Slotkin, J., Blitz, D., Wallner-Allen, K., Fox, N. A., Beaumont, J. L., Mungas, D., Nowinski, C. J., Richler, J., Deocampo, J. A., Anderson, J. E., Manly, J. J., Borosh, B., … Gershon, R. C. (2013). Cognition assessment using the NIH Toolbox. *Neurology*, *80*(11 Suppl 3), S54-64. https://doi.org/10.1212/WNL.0b013e3182872ded

Wickham, H. (2016). *ggplot2: Elegant Graphics for Data Analysis*. Springer.
