## Supplementary material for "Triangulating Causality Between Childhood Obesity and Neurobehavior: Twin and Longitudinal Evidence": commented analysis script: Rscript_2022-06-15.R

```
## ABCD-BMI R Analysis Script - Version 01

############################################################################
#
# Triangulating Causality Between Childhood Obesity and Neurobehavior
# Twin and Longitudinal Evidence
#
# Leonard Konstantin Kulisch, Kadri Arumäe, Daniel A. Briley, Uku Vainik
#
# 2022
# 
# Corresponding author: Uku Vainik 
#
############################################################################

#########################
#
# Part I: CREATE DATA SET
#
######################### ----

rm(list = ls())

## "SWITCH BOARD"

# set time to be analyzed:
setTime = "1_year_follow_up_y_arm_1" # choose "baseline_year_1_arm_1" for Baseline or "1_year_follow_up_y_arm_1" / "2_year_follow_up_y_arm_1" for FUs

## PREPARATION

# set wd
setwd("~/Job/Tartu Traineeship/R Analysis/abcd_bmi")

# packages
library(stringr)
library(zscorer)
library(childsds)

# read files
dem <- read.table("rawdata/pdem02.txt", sep="\t", dec=".", header=TRUE) # Demographics
lng <- read.table("rawdata/abcd_lpds01.txt", sep="\t", dec=".", header=TRUE) # Longitudinal Info
hnd <- read.table("rawdata/abcd_ehis01.txt", sep="\t", dec=".", header=TRUE) # Handedness
fam <- read.table("rawdata/acspsw03.txt", sep="\t", dec=".", header=TRUE) # Family Relations
pub <- read.table("rawdata/abcd_ppdms01.txt", sep="\t", dec=".", header=TRUE) # puberty info
med <- read.table("rawdata/abcd_mx01.txt", sep="\t", dec=".", header=TRUE) # past medical conditions
pha <- read.table("rawdata/medsy01.txt", sep="\t", dec=".", header=TRUE, fill=TRUE) # medication taken
sit <- read.table("rawdata/participants.tsv", sep="\t", dec=".", header=TRUE, fill=TRUE) # processed data incl site info
mriqc <- read.table("rawdata/abcd_imgincl01.txt", sep="\t", dec=".", header=TRUE) # MRI quality control

ant <- read.table("rawdata/abcd_ant01.txt", sep="\t", dec=".", header=TRUE) # Anthropometrics

mhp <- read.table("rawdata/abcd_mhp02.txt", sep="\t", dec=".", header=TRUE) # Sum Scores Mental Health/Personality Parent-report
mhs <- read.table("rawdata/abcd_mhy02.txt", sep="\t", dec=".", header=TRUE) # Sum Scores Mental Health/Personality Self-report
cbc <- read.table("rawdata/abcd_cbcls01.txt", sep="\t", dec=".", header=TRUE) # Child Behvior Checklist 
psb <- read.table("rawdata/abcd_psb01.txt", sep="\t", dec=".", header=TRUE) # Prosocial Behavior (self-report)
poa <- read.table("rawdata/abcd_yssbpm01.txt", sep="\t", dec=".", header=TRUE) # Positive Affect
gdc <- read.table("rawdata/abcd_gdss01.txt", sep="\t", dec=".", header=TRUE) # Game of Dice 
cct <- read.table("rawdata/cct01.txt", sep="\t", dec=".", header=TRUE) # Cash Choice Task 

nih <- read.table("rawdata/abcd_tbss01.txt", sep="\t", dec=".", header=TRUE) # NIH Toolbox Sum Scores
prs <- read.table("rawdata/abcd_ps01.txt", sep="\t", dec=".", header=TRUE) # Pearson: Verbal Learning, Matrix reasoning
lmt <- read.table("rawdata/lmtp201.txt", sep="\t", dec=".", header=TRUE) # Little Man Task
dds <- read.table("rawdata/abcd_yddss01.txt", sep="\t", dec=".", header=TRUE) # Delay Discounting Task
est <- read.table("rawdata/abcd_yest01.txt", sep="\t", dec=".", header=TRUE) # Emotional Stroop Task

mri <- read.table("rawdata/abcd_smrip101.txt", sep="\t", dec=".", header=TRUE) # MRI scan files

it1 <- read.table("rawdata/abcd_pssrs01.txt", sep="\t", dec=".", header=TRUE) # Item Level Data SSRS Social Responsiveness
it2 <- read.table("rawdata/abcd_eatqp01.txt", sep="\t", dec=".", header=TRUE) # Item Level Data EAT-Q
it3 <- read.table("rawdata/abcd_upps01.txt", sep="\t", dec=".", header=TRUE) # Item Level Data UPPS-P
it4 <- read.table("rawdata/abcd_bisbas01.txt", sep="\t", dec=".", header=TRUE) # Item Level Data BIS/BAS
it5 <- read.table("rawdata/abcd_cbcl01.txt", sep="\t", dec=".", header=TRUE) # Item Level Data CBCL
it6 <- read.table("rawdata/abcd_psb01.txt", sep="\t", dec=".", header=TRUE) # Item Level Data Prosocial Behavior
it7 <- read.table("rawdata/abcd_ytbpai01.txt", sep="\t", dec=".", header=TRUE) # Item Level Data Positive Affect

## ID AND CONTROL VARIABLES

data <- data.frame(Sep_IdControls=NA)

# prepare demographics
dem <- dem[c("subjectkey", "demo_sex_v2", "demo_prim", "demo_race_a_p___10", "demo_race_a_p___11", "demo_race_a_p___12", "demo_race_a_p___13", "demo_race_a_p___14", "demo_race_a_p___15", "demo_race_a_p___16", "demo_race_a_p___17", "demo_race_a_p___18", "demo_race_a_p___19", "demo_race_a_p___20", "demo_race_a_p___21", "demo_race_a_p___22", "demo_race_a_p___23","demo_race_a_p___24", "demo_race_a_p___25", "demo_ethn_v2", "demo_prnt_prtnr_v2", "demo_comb_income_v2", "demo_prnt_ed_v2",  	"demo_prtnr_ed_v2", "demo_prnt_marital_v2")]
dem <- dem[-1,]
names(dem) <- c("ID", "sex", "infosource", "white", "black", "native", "alaskan", "hawaiian","guamanian","samoan","pacificIslander","asianIndian","chinese","filipino","japanese","korean","vietnamese","otherAsian","otherRace", "hispanic", "singleParent", "income", "collegemat", "collegepat", "married")
dem[dem == 999 | dem == 777 | dem == ""] <- NA
levels(dem$sex) <- c(0, 1, 2, NA) # male 0/ female 1/ div 2
dem$infosource <- droplevels(dem$infosource) # 1 bio mother, 2 bio father, 3 adoptive parent, 4 custodial parent, 5 other
dem[,4:19] <- lapply(dem[,4:19], as.character)
dem[,4:19] <- lapply(dem[,4:19], as.numeric)
dem$race[dem$white==1 & rowSums(dem[,4:19])==1] <- 0 # race: white 0
dem$race[dem$black==1 & rowSums(dem[,4:19])==1] <- 1 # race: black/african 1
dem$race[(dem$white!=1 & dem$black!=1) | rowSums(dem[,4:19])!=1] <- 2 # race: other/multiple 2
dem$race <- as.factor(dem$race)
levels(dem$hispanic) <- c(NA, 1, 0, NA, NA, NA) # hispanic yes 1/no 0
levels(dem$singleParent) <- c(NA, 0, 1, NA, NA) # single parent household yes 1/no 0
dem$income <- droplevels(dem$income) # 1 <5k, 2 5k-11(,999)k, 3 12-15k, 4 16-24k, 5 25-34k, 6 35-49k, 7 50-74k, 8 75-99k, 9 100-199k, 10 >200k
levels(dem$collegemat) <- c(0, 0, 0, 0, 0, 0, 1, 1, 1, 1, 1, 0, 1, 1, 0, 0, 0, 0, 0, NA, 0, 0, NA) # college degree yes 1/no 2
levels(dem$collegepat) <- c(NA, 0, 0, 0, 0, 0, 0, 0, 1, 1, 1, 1, 1, 0, 1, 1, 0, 0, 0, 0, 0, NA, 0, 0, NA, NA) 
levels(dem$married) <- c(NA, 1, 0, 0, 0, 0, 0, NA, NA) # married yes 1/no 0
dem <- dem[c("ID", "sex", "infosource", "race", "hispanic", "singleParent", "income", "collegemat", "collegepat", "married")]
data <- cbind(data, dem)

# prepare longitudinal info (i.e. age at time of measurement)
lng <- lng[c("subjectkey", "eventname", "interview_age")]
lng <- lng[-1,]
names(lng) <- c("ID", "time", "age")
lng$age <- as.numeric(as.character(lng$age)) # age in months
lng <- lng[lng$time == setTime,]
lng <- lng[,-2]
data <- merge(data, lng, by="ID", all = T)

# prepare handedness
hnd <- hnd[c("subjectkey", "eventname", "ehi_y_ss_scoreb")]
names(hnd) <- c("ID", "time", "handedness")
hnd <- hnd[hnd$time == "baseline_year_1_arm_1",] # was only asked at baseline
hnd$handedness <- droplevels(hnd$handedness) # right-handed 1/ left-handed 2/ mixed-handed 3
hnd <- hnd[,-2]
data <- merge(data, hnd, by="ID", all = T)

# prepare family relations
fam <- fam[c("subjectkey", "eventname", "rel_relationship", "rel_family_id", "rel_group_id", "rel_ingroup_order", "genetic_zygosity_status_1", "genetic_zygosity_status_2", "genetic_zygosity_status_3", "genetic_zygosity_status_4")]
names(fam) <- c("ID", "time", "siblings", "family", "famgroup", "twingroup", "zygosity1", "zygosity2", "zygosity3", "zygosity4")
fam[fam == 999 | fam == 777 | fam == ""] <- NA
fam <- fam[fam$time == "baseline_year_1_arm_1",] # was only asked at baseline
fam[,3:10] <- lapply(fam[,3:10], droplevels)
# siblings = 0 singleton, 1 sibling, 2 twin, 3 triplet 
# family = unique ID per family
# famgroup = twin/triplets share same value (differentiation from other sibs)
# twingroup = twin/triplets have different values (differentiation between twins/triplets)
# zygosity = 1 monozygotic, 2 dizygotic, 3 siblings
fam <- fam[,-2]
data <- merge(data, fam, by="ID", all = T)

# prepare puberty/birth control info
pub <- pub[c("subjectkey", "eventname", "pds_f5b_p", "menstrualcycle4_p", "pds_1_p", "pds_2_p", "pds_3_p", "pds_f4_p", "pds_f5b_p", "pds_m4_p", "pds_m5_p")]
pub <- pub[-1,]
names(pub) <- c("ID", "time", "menstruation", "birthcontrol", "pub_ht", "pub_hair", "pub_skin", "pub_breast", "pub_period", "pub_voice", "pub_beard")
pub[pub == 999 | pub == 777 | pub == ""] <- NA
levels(pub$menstruation) <- c(NA, 0, 1, NA, NA) # menstruation started yes 1/ no 0
pub$birthcontrol <- droplevels(pub$birthcontrol) # birth control yes 1/no 0
pub[,4:11] <- lapply(pub[,4:11], as.character)
pub[,4:11] <- lapply(pub[,4:11], as.numeric)
pub$puberty_m <- ifelse(is.na(pub$pub_ht)==F & is.na(pub$pub_skin)==F & is.na(pub$pub_hair)==F & is.na(pub$pub_voice)==F & is.na(pub$pub_beard)==F, (pub$pub_ht + pub$pub_skin + pub$pub_hair + pub$pub_voice + pub$pub_beard)/5, NA)
pub$puberty_f <- ifelse(is.na(pub$pub_ht)==F & is.na(pub$pub_skin)==F & is.na(pub$pub_hair)==F & is.na(pub$pub_breast)==F & is.na(pub$pub_period)==F, (pub$pub_ht + pub$pub_skin + pub$pub_hair + pub$pub_breast + pub$pub_period)/5, NA)
pub$puberty <- ifelse(is.na(pub$puberty_m)==F, pub$puberty_m, pub$puberty_f)
pub <- pub[c("ID", "time", "menstruation", "birthcontrol", "puberty")]
pub <- pub[pub$time == setTime,]
pub <- pub[,-2]
data <- merge(data, pub, by="ID", all = T)

# generate sex+menstruation variable
data$sex_menstruation <- data$sex
levels(data$sex_menstruation) <- c(0, 1, 3)
levels(data$sex_menstruation) <- c(levels(data$sex_menstruation), 2)
data$sex_menstruation[data$menstruation==1 & data$sex==1] <- 2 # 0 male, 1 female no menstruation, 2 female menstruation, 3 other sex

# prepare past medical conditions
med <- med[c("subjectkey", "medhx_2g", "medhx_2c", "medhx_2e", "medhx_2f", "medhx_2h")]
med <- med[-1,]
names(med) <- c("ID", "diabetes", "braininjury", "cancer", "cerebralpalsy", "epilepsy")
med[med == ""] <- NA
med[,2:6] <- lapply(med[,2:6], droplevels)
data <- merge(data, med, by="ID", all = T)

# prepare medication taken
pha <- pha[c("subjectkey", "eventname", "med1_rxnorm_p", "med2_rxnorm_p", "med3_rxnorm_p", "med4_rxnorm_p", "med5_rxnorm_p", "med6_rxnorm_p", "med7_rxnorm_p", "med8_rxnorm_p", "med9_rxnorm_p", "med10_rxnorm_p", "med11_rxnorm_p", "med12_rxnorm_p", "med13_rxnorm_p", "med14_rxnorm_p", "med15_rxnorm_p")]
pha <- pha[-1,]
names(pha) <- c("ID", "time", "med1", "med2", "med3", "med4", "med5", "med6", "med7", "med8", "med9", "med10", "med11", "med12", "med13", "med14", "med15")
pha$stimulants <- ifelse(str_count(pha$med1, '11706|118760|116812|117188|131258|118534|118504|160069') + 
                           str_count(pha$med2, '11706|118760|116812|117188|131258|118534|118504|160069') +
                           str_count(pha$med3, '11706|118760|116812|117188|131258|118534|118504|160069') +
                           str_count(pha$med4, '11706|118760|116812|117188|131258|118534|118504|160069') +
                           str_count(pha$med5, '11706|118760|116812|117188|131258|118534|118504|160069') +
                           str_count(pha$med6, '11706|118760|116812|117188|131258|118534|118504|160069') +
                           str_count(pha$med7, '11706|118760|116812|117188|131258|118534|118504|160069') +
                           str_count(pha$med8, '11706|118760|116812|117188|131258|118534|118504|160069') +
                           str_count(pha$med9, '11706|118760|116812|117188|131258|118534|118504|160069') +
                           str_count(pha$med10, '11706|118760|116812|117188|131258|118534|118504|160069') +
                           str_count(pha$med11, '11706|118760|116812|117188|131258|118534|118504|160069') +
                           str_count(pha$med12, '11706|118760|116812|117188|131258|118534|118504|160069') +
                           str_count(pha$med13, '11706|118760|116812|117188|131258|118534|118504|160069') +
                           str_count(pha$med14, '11706|118760|116812|117188|131258|118534|118504|160069') +
                           str_count(pha$med15, '11706|118760|116812|117188|131258|118534|118504|160069') > 0, 1, 0)
pha <- pha[c("ID", "time", "stimulants")]
pha <- pha[pha$time == setTime,]
pha <- pha[,-2]
data <- merge(data, pha, by="ID", all = T)

# prepare site info
sit <- sit[c("participant_id", "site")]
sit$ID <- as.character(sit$participant_id)
sit$ID <- substring(sit$ID, 9)
sit$ID <- paste("NDAR_", sit$ID, sep = "")
sit <- sit[,-1]
sit[sit == "888" | sit == ""] <- NA
data <- merge(data, sit, by="ID", all = T)

# prepare MRI quality control
mriqc <- mriqc[c("subjectkey", "eventname", "imgincl_t1w_include")]
mriqc <- mriqc[-1,]
names(mriqc) <- c("ID", "time", "mriqc")
mriqc <- mriqc[mriqc$time == setTime,]
mriqc <- mriqc[,-2]
data <- merge(data, mriqc, by="ID", all = T)

## OBESITY VARIABLES

data$Sep_Obesity <- NA

# prepare BMI & RFMp
ant <- ant[c("subjectkey", "eventname", "anthroheightcalc", "anthroweightcalc", "anthro_waist_cm", "interview_age", "sex")]
ant <- ant[-1,]
names(ant) <- c("ID", "time", "height_in", "weight_lbs", "waist_in", "ant_age", "ant_sex")
ant[,3:6] <- lapply(ant[,3:6], as.character)
ant[,3:6] <- lapply(ant[,3:6], as.numeric)
ant$height <- ant$height_in * 0.0254
ant$weight <- ant$weight_lbs / 2.2046
ant$BMI <- ant$weight / (ant$height * ant$height) 
levels(ant$ant_sex) <- c("2", "1", NA)
ant$ant_sex_num <- as.numeric(as.character(ant$ant_sex))
ant$height_cm <- ant$height * 100
ant$ant_age_days <- ant$ant_age * 30.417
ant <- addWGSR(data = ant, sex = "ant_sex_num", firstPart = "weight", secondPart = "height_cm", thirdPart = "ant_age_days", index = "bfa")
ant$BMIz <- ant$bfaz
ant$ant_age_yrs <- ant$ant_age / 12
ant$ant_sex_ch <- ant$ant_sex
levels(ant$ant_sex_ch) <- c("female", "male")
ant$ant_sex_ch <- as.character(ant$ant_sex_ch)
BMI_CDC_percentiles <- make_percentile_tab(ref = cdc.ref, item = "bmi", perc = 95, stack = F)
BMI_CDC_percentiles$age <- round(BMI_CDC_percentiles$age, 1)
BMI_CDC_percentiles <- aggregate(x = BMI_CDC_percentiles$perc_95_0, by = list(BMI_CDC_percentiles$age, BMI_CDC_percentiles$sex), FUN = mean)
ant$BMI95_percentile <- NA
for (i in 1:nrow(ant)) {
  BMI_percentile_age <- round(ant[i,]$ant_age_yrs, 1)
  BMI_percentile_sex <- ant[i,]$ant_sex_ch
  ant[i,]$BMI95_percentile <- BMI_CDC_percentiles[BMI_CDC_percentiles$Group.1 == BMI_percentile_age & BMI_CDC_percentiles$Group.2 == BMI_percentile_sex, 3]
}
ant$BMI95 <- ant$BMI / ant$BMI95_percentile * 100
levels(ant$ant_sex) <- c("1", "0")
ant$ant_sex <- as.numeric(as.character(ant$ant_sex))
ant$RFMp <- 74 - (22 * ant$height_in / ant$waist_in) + (5 * ant$ant_sex)
ant <- ant[c("ID", "time", "BMI", "BMIz", "BMI95", "RFMp")]
# issue: many wrong entries e.g. hight measure of some kids stated as 0 inch , 128 inch (>3m), 5 inch (13cm), ...
# ant$BMI[ant$BMI > 100] <- NA # NA-s cases with wrong BMI or RFMp
ant <- ant[ant$time == setTime,]
ant <- ant[,-2]
data <- merge(data, ant, by="ID", all = T)

## PERSONALITY VARIABLES

data$Sep_PersScales <- NA

# prepare Mental Health/Personality Questionnaires Parent-Report
mhp <- mhp[c("subjectkey", "eventname", "ssrs_p_ss_sum", "eatq_p_ss_attention", "eatq_p_ss_depressive_mood", "eatq_p_ss_fear", "eatq_p_ss_frustration", "eatq_p_ss_inhibitory", "eatq_p_ss_activation", "eatq_p_ss_shyness", "eatq_p_ss_surgency", "eatq_p_ss_affiliation", "eatq_p_ss_aggression")]
mhp <- mhp[-1,]
names(mhp) <- c("ID", "time", "socialresponsiveness", "eat_attention", "eat_depressive", "eat_fear", "eat_frustration", "eat_inhibitory", "eat_activation", "eat_shyness", "eat_surgency", "eat_affiliation", "eat_aggression")
mhp[,3:13] <- lapply(mhp[,3:13], as.character)
mhp[,3:13] <- lapply(mhp[,3:13], as.numeric)
mhp <- mhp[mhp$time == setTime,] 
mhp <- mhp[,-2]
if (nrow(mhp) > 0) {
data <- merge(data, mhp, by="ID", all = T) }

# prepare Mental Health/Personality Questionnaires Self-Report
mhs <- mhs[c("subjectkey", "eventname", "upps_y_ss_negative_urgency", "upps_y_ss_lack_of_planning", "upps_y_ss_sensation_seeking", "upps_y_ss_positive_urgency", "upps_y_ss_lack_of_perseverance", "bis_y_ss_bism_sum", "bis_y_ss_basm_rr", "bis_y_ss_basm_drive")]
mhs <- mhs[-1,]
names(mhs) <- c("ID", "time", "upps_negurgency", "upps_planning", "upps_sensation", "upps_posurgency", "upps_perseverance", "bis_inhibition", "bis_reward", "bis_drive")
mhs[,3:10] <- lapply(mhs[,3:10], as.character)
mhs[,3:10] <- lapply(mhs[,3:10], as.numeric)
mhs <- mhs[mhs$time == setTime,] 
mhs <- mhs[,-2]
if (nrow(mhs) > 0) {
data <- merge(data, mhs, by="ID", all = T) }

# prepare Child Behavior Checklist
cbc <- cbc[c("subjectkey", "eventname", "cbcl_scr_syn_anxdep_r", "cbcl_scr_syn_withdep_r", "cbcl_scr_syn_somatic_r", "cbcl_scr_syn_social_r", "cbcl_scr_syn_thought_r", "cbcl_scr_syn_attention_r", "cbcl_scr_syn_rulebreak_r", "cbcl_scr_syn_aggressive_r")]
cbc <- cbc[-1,]
names(cbc) <- c("ID", "time", "cbcl_anxietydep", "cbcl_depression", "cbcl_somatic", "cbcl_social", "cbcl_thought", "cbcl_attention", "cbcl_rulebreak", "cbcl_aggression")
cbc[,3:10] <- lapply(cbc[,3:10], as.character)
cbc[,3:10] <- lapply(cbc[,3:10], as.numeric)
cbc <- cbc[cbc$time == setTime,] 
cbc <- cbc[,-2]
if (nrow(cbc) > 0) {
data <- merge(data, cbc, by="ID", all = T) }

# prepare Prosocial Behavior (Self-report)
psb <- psb[c("subjectkey", "eventname", "prosocial_q1_y", "prosocial_q2_y", "prosocial_q3_y")]
psb <- psb[-1,]
names(psb) <- c("ID", "time", "psb_1", "psb_2", "psb_3")
psb[,3:5] <- lapply(psb[,3:5], as.character)
psb[,3:5] <- lapply(psb[,3:5], as.numeric)
psb$prosociality <- ifelse(is.na(psb$psb_1)==F & is.na(psb$psb_2)==F & is.na(psb$psb_3)==F, psb$psb_1 + psb$psb_2 + psb$psb_3, NA)
psb <- psb[c("ID", "time", "prosociality")]
psb <- psb[psb$time == setTime,] 
psb <- psb[,-2]
if (nrow(psb) > 0) {
data <- merge(data, psb, by="ID", all = T) }

# prepare positive affect
poa <- poa[c("subjectkey", "eventname", "poa_y_ss_sum")]
poa <- poa[-1,]
names(poa) <- c("ID", "time", "posaffect")
poa$posaffect <- as.numeric(as.character(poa$posaffect))
poa <- poa[poa$time == setTime,] 
poa <- poa[,-2]
if (nrow(poa) > 0) {
data <- merge(data, poa, by="ID", all = T) }

# prepare Game of Dice
gdc <- gdc[c("subjectkey", "eventname", "gdt_scr_expressions_net_score")]
gdc <- gdc[-1,]
names(gdc) <- c("ID", "time", "gameofdice")
gdc$gameofdice <- as.numeric(as.character(gdc$gameofdice))
gdc <- gdc[gdc$time == setTime,] 
gdc <- gdc[,-2]
if (nrow(gdc) > 0) {
data <- merge(data, gdc, by="ID", all = T) }

# prepare Cash Choice Task
cct <- cct[c("subjectkey", "eventname", "cash_choice_task")]
cct <- cct[-1,]
names(cct) <- c("ID", "time", "cashchoice")
cct$cashchoice <- as.numeric(as.character(cct$cashchoice))
cct[cct == 3] <- NA
cct <- cct[gdc$time == setTime,] 
cct <- cct[,-2]
if (nrow(cct) > 0) {
data <- merge(data, cct, by="ID", all = T) }

## COGNITION VARIABLES

data$Sep_Cognition <- NA

# prepare NIH Toolbox (here selected age-uncorrected results, bc correction will happen later (age, sex, race, ...))
nih <- nih[c("subjectkey", "eventname","nihtbx_picvocab_uncorrected", "nihtbx_flanker_uncorrected", "nihtbx_list_uncorrected", "nihtbx_cardsort_uncorrected", "nihtbx_pattern_uncorrected", "nihtbx_picture_uncorrected", "nihtbx_reading_uncorrected")]
nih <- nih[-1,]
names(nih) <- c("ID", "time", "nih_picvoc", "nih_flanker", "nih_list", "nih_card", "nih_pattern", "nih_picture", "nih_reading")
nih[,3:9] <- lapply(nih[,3:9], as.character)
nih[,3:9] <- lapply(nih[,3:9], as.numeric)
nih <- nih[nih$time == setTime,] 
nih <- nih[,-2]
if (nrow(nih) > 0) {
data <- merge(data, nih, by="ID", all = T) }

# prepare Pearson Tests
prs <- prs[c("subjectkey", "eventname","pea_ravlt_sd_trial_i_tc", "pea_ravlt_sd_trial_ii_tc", "pea_ravlt_sd_trial_iii_tc", "pea_ravlt_sd_trial_iv_tc", "pea_ravlt_sd_trial_v_tc", "pea_ravlt_sd_trial_vi_tc", "pea_ravlt_ld_trial_vii_tc", "pea_wiscv_trs")]
prs <- prs[-1,]
names(prs) <- c("ID", "time", "ravlt_1", "ravlt_2", "ravlt_3", "ravlt_4", "ravlt_5", "ravlt_immediaterecall", "ravlt_delayedrecall", "matrixreasoning")
prs[,3:10] <- lapply(prs[,3:10], as.character)
prs[,3:10] <- lapply(prs[,3:10], as.numeric)
prs$ravlt_learning <- ifelse(is.na(prs$ravlt_1)==F & is.na(prs$ravlt_2)==F & is.na(prs$ravlt_3)==F & is.na(prs$ravlt_4)==F & is.na(prs$ravlt_5)==F, (prs$ravlt_1 + prs$ravlt_2 + prs$ravlt_3 + prs$ravlt_4 + prs$ravlt_5)/5, NA)
prs <- prs[c("ID", "time", "ravlt_learning", "ravlt_immediaterecall", "ravlt_delayedrecall", "matrixreasoning")]
prs <- prs[prs$time == setTime,] 
prs <- prs[,-2]
if (nrow(prs) > 0) {
data <- merge(data, prs, by="ID", all = T) }

# prepare Little Man Task
lmt <- lmt[c("subjectkey", "eventname", "lmt_scr_num_correct", "lmt_scr_avg_rt")]
lmt <- lmt[-1,]
names(lmt) <- c("ID", "time", "lmt_correct", "lmt_RT")
lmt[,3:4] <- lapply(lmt[,3:4], as.character)
lmt[,3:4] <- lapply(lmt[,3:4], as.numeric)
lmt <- lmt[lmt$time == setTime,] 
lmt <- lmt[,-2]
if (nrow(lmt) > 0) {
data <- merge(data, lmt, by="ID", all = T) }

# prepare Delay Discounting Task
dds <- dds[c("subjectkey", "eventname", "ddis_scr_val_indif_point_6h", "ddis_scr_val_indif_pnt_1da", "ddis_scr_val_indif_pnt_1week", "ddis_scr_val_indif_pnt_1mth", "ddis_scr_val_indif_pnt_3mth", "ddis_scr_val_indif_pnt_1yr", "ddis_scr_val_indif_pnt_5yr", "ddis_scr_expr_mnrt_allcho")]
dds <- dds[-1,]
names(dds) <- c("ID", "time", "dd_6h", "dd_day", "dd_week", "dd_1m", "dd_3m", "dd_1y", "dd_5y", "dd_RT")
dds[,3:10] <- lapply(dds[,3:10], as.character)
dds[,3:10] <- lapply(dds[,3:10], as.numeric)
dds <- dds[dds$time == setTime,] 
dds <- dds[,-2]
if (nrow(dds) > 0) {
data <- merge(data, dds, by="ID", all = T) }

# prepare Emotional Stroop Task
est <- est[c("subjectkey", "eventname", "strp_scr_acc_all", "strp_scr_mnrt_all")]
est <- est[-1,]
names(est) <- c("ID", "time", "estroop_correct", "estroop_RT")
est[,3:4] <- lapply(est[,3:4], as.character)
est[,3:4] <- lapply(est[,3:4], as.numeric)
est <- est[est$time == setTime,] 
est <- est[,-2]
if (nrow(est) > 0) {
data <- merge(data, est, by="ID", all = T) }

## BRAIN MORPHOLOGY VARIABLES

# prepare MRI data
data$Sep_CT <- NA
mr1 <- mri[,c(4, 9, 11:78, 117)]
mr1 <- mr1[-1,]
names(mr1) <- c("ID", "time", names(mr1[,3:70]), "meanthickness")
mr1[,3:71] <- lapply(mr1[,3:71], as.character)
mr1[,3:71] <- lapply(mr1[,3:71], as.numeric)
mr1 <- mr1[mr1$time == setTime,] 
mr1 <- mr1[,-2]
data <- merge(data, mr1, by="ID", all = T)

data$Sep_CA <- NA
mr2 <- mri[,c(4, 9, 225:292, 331)]
mr2 <- mr2[-1,]
names(mr2) <- c("ID", "time", names(mr2[,3:70]), "totalarea")
mr2[,3:71] <- lapply(mr2[,3:71], as.character)
mr2[,3:71] <- lapply(mr2[,3:71], as.numeric)
mr2 <- mr2[mr2$time == setTime,] 
mr2 <- mr2[,-2]
data <- merge(data, mr2, by="ID", all = T)

data$Sep_CV <- NA
mr3 <- mri[,c(4, 9, 332:399, 438)]
mr3 <- mr3[-1,]
names(mr3) <- c("ID", "time", names(mr3[,3:70]), "totalvolume")
mr3[,3:71] <- lapply(mr3[,3:71], as.character)
mr3[,3:71] <- lapply(mr3[,3:71], as.numeric)
mr3 <- mr3[mr3$time == setTime,] 
mr3 <- mr3[,-2]
data <- merge(data, mr3, by="ID", all = T)

## ITEM LEVEL DATA

data$Sep_PersItems <- NA

# prepare SSRS Social Responsiveness Questionnaire Items
it1 <- it1[,c(4, 9, 11:(ncol(it1)-2))]
it1 <- it1[-1,]
it1[it1 == 999 | it1 == 777 | it1 == ""] <- NA
names(it1) <- c("ID", "time", names(it1[,3:ncol(it1)]))
it1[,3:ncol(it1)] <- lapply(it1[,3:ncol(it1)], as.character)
it1[,3:ncol(it1)] <- lapply(it1[,3:ncol(it1)], as.numeric)
it1 <- it1[it1$time == setTime,] 
it1 <- it1[,-2]
if (nrow(it1) > 0) {
data <- merge(data, it1, by="ID", all = T) }

# prepare EAT-Q Questionnaire Items
it2 <- it2[,c(4, 9, 11:(ncol(it2)-2))]
it2 <- it2[-1,]
it2[it2 == 999 | it2 == 777 | it2 == ""] <- NA
names(it2) <- c("ID", "time", names(it2[,3:ncol(it2)]))
it2[,3:ncol(it2)] <- lapply(it2[,3:ncol(it2)], as.character)
it2[,3:ncol(it2)] <- lapply(it2[,3:ncol(it2)], as.numeric)
it2 <- it2[it2$time == setTime,] 
it2 <- it2[,-2]
if (nrow(it2) > 0) {
data <- merge(data, it2, by="ID", all = T) }

# prepare UPPS-P Questionnaire Items
it3 <- it3[,c(4, 9:(ncol(it3)-2))]
it3 <- it3[-1,]
it3[it3 == 999 | it3 == 777 | it3 == ""] <- NA
names(it3) <- c("ID", "time", names(it3[,3:ncol(it3)]))
it3[,3:ncol(it3)] <- lapply(it3[,3:ncol(it3)], as.character)
it3[,3:ncol(it3)] <- lapply(it3[,3:ncol(it3)], as.numeric)
it3 <- it3[it3$time == setTime,] 
it3 <- it3[,-2]
if (nrow(it3) > 0) {
data <- merge(data, it3, by="ID", all = T) }

# prepare BIS/BAS Items
it4 <- it4[,c(4, 9:(ncol(it4)-2))]
it4 <- it4[-1,]
it4[it4 == 999 | it4 == 777 | it4 == ""] <- NA
names(it4) <- c("ID", "time", names(it4[,3:ncol(it4)]))
it4[,3:ncol(it4)] <- lapply(it4[,3:ncol(it4)], as.character)
it4[,3:ncol(it4)] <- lapply(it4[,3:ncol(it4)], as.numeric)
it4 <- it4[it4$time == setTime,] 
it4 <- it4[,-2]
if (nrow(it4) > 0) {
data <- merge(data, it4, by="ID", all = T) }

# prepare CBCL Items
it5 <- it5[,c(4, 129, 10:(ncol(it5)-4))]
it5 <- it5[-1,]
it5[it5 == 999 | it5 == 777 | it5 == ""] <- NA
names(it5) <- c("ID", "time", names(it5[,3:ncol(it5)]))
it5[,3:ncol(it5)] <- lapply(it5[,3:ncol(it5)], as.character)
it5[,3:ncol(it5)] <- lapply(it5[,3:ncol(it5)], as.numeric)
it5 <- it5[it5$time == setTime,] 
it5 <- it5[,-2]
if (nrow(it5) > 0) {
data <- merge(data, it5, by="ID", all = T) }

# prepare Prosocial Behavior Items
it6 <- it6[,c(4, 9:(ncol(it6)-2))]
it6 <- it6[-1,]
it6[it6 == 999 | it6 == 777 | it6 == ""] <- NA
names(it6) <- c("ID", "time", names(it6[,3:ncol(it6)]))
it6[,3:ncol(it6)] <- lapply(it6[,3:ncol(it6)], as.character)
it6[,3:ncol(it6)] <- lapply(it6[,3:ncol(it6)], as.numeric)
it6 <- it6[it6$time == setTime,] 
it6 <- it6[,-2]
if (nrow(it6) > 0) {
data <- merge(data, it6, by="ID", all = T) }

# prepare Positive Affect Items
it7 <- it7[,c(4, 9:(ncol(it7)-2))]
it7 <- it7[-1,]
it7[it7 == 999 | it7 == 777 | it7 == ""] <- NA
names(it7) <- c("ID", "time", names(it7[,3:ncol(it7)]))
it7[,3:ncol(it7)] <- lapply(it7[,3:ncol(it7)], as.character)
it7[,3:ncol(it7)] <- lapply(it7[,3:ncol(it7)], as.numeric)
it7 <- it7[it7$time == setTime,] 
it7 <- it7[,-2]
if (nrow(it7) > 0) {
data <- merge(data, it7, by="ID", all = T) }

## Reorder variables to correct sections
data$Sep_Eating <- NA
data$Sep_Calculations <- NA
data <- subset(data, select=c(ID:mriqc, meanthickness, totalarea, totalvolume, Sep_Obesity:smri_thick_cdk_insularh, 
                              Sep_CA:smri_area_cdk_insularh, Sep_CV:smri_vol_cdk_insularh, Sep_PersItems:cbcl_q23_p, 
                              cbcl_q25_p:cbcl_q52_p, cbcl_q54_p:prosocial_q3_y, Sep_Eating, cbcl_q24_p, cbcl_q53_p, 
                              Sep_Calculations))

#### SAVE DATA FRAME
save(data, file="rsaves/data-1yFU-orig_2022-02-18.Rda")
write.csv(data, file="rsaves/data-1yFU-orig_2022-02-18.csv", row.names = FALSE)
rm(list = ls())

############################################################################

#########################
#
# Part II: CLEAN DATA SET & SIMPLE CORRELATIONS (SINGLETONS) 
#
######################### ----

rm(list = ls())

## "SWITCH BOARD"
#-

## PREPARATION

# set wd
setwd("~/Job/Tartu Traineeship/R Analysis/abcd_bmi")

# packages
library(psych)
library(ggplot2)
library(ggseg)
library(ggseg3d)
library(gridExtra)
library(ggpubr)
library(dplyr)
library(tidyr)
library(patchwork)

# read files

data <- read.csv("rsaves/data-BL-orig_2022-02-18.csv", sep=",", dec=".", header=TRUE) # read Baseline

## DROP VARIABLES NOT IN THE DATA SET

# for Baseline:
data <- data[,-c(which(colnames(data)=="socialresponsiveness"):which(colnames(data)=="eat_aggression"))] # delete instruments not in the set
data <- data[,-which(colnames(data)=="bisbas5_y")] # delete BIS/BAS item that was never answered
# for 2-year-FU:
# data <- ... 

# work on Eating items
data <- data[,-which(colnames(data)=="cbcl_q55_p")] #delete item "Overweight"
data$cbcl_q24_p <- as.factor(data$cbcl_q24_p) # make Eating items 0/1 bc of low freq of "2"
levels(data$cbcl_q24_p) <- c(0, 1, 1)
data$cbcl_q24_p <- as.numeric(as.character(data$cbcl_q24_p))
data$cbcl_q53_p <- as.factor(data$cbcl_q53_p)
levels(data$cbcl_q53_p) <- c(0, 1, 1)
data$cbcl_q53_p <- as.numeric(as.character(data$cbcl_q53_p))

## EXCLUDE CASES

data_raw <- data
nrow(data) # 11.933 participants at Basline total

data <- data[is.na(data$sex)==F & +
               is.na(data$puberty)==F & +
               is.na(data$race)==F & +
               is.na(data$hispanic)==F & +
               is.na(data$age)==F & +
               #is.na(data$RFMp)==F & +
               is.na(data$BMI95)==F,]
NrExcluded <- as.numeric(nrow(data_raw) - nrow(data))
SumExcluded <- NrExcluded
print(c("Excluded for Missing Important Values: ", NrExcluded))

data <- data[data$diabetes == 0 & data$braininjury == 0 & +
               data$cancer == 0 & data$cerebralpalsy == 0 & +
               data$epilepsy == 0,]
NrExcluded <- as.numeric(nrow(data_raw) - nrow(data)) - SumExcluded
SumExcluded <- SumExcluded + NrExcluded
print(c("Excluded for Medical Conditions: ", NrExcluded))

data <- data[data$BMIz <= 5,] # & data$RFMp > 0,]
NrExcluded <- as.numeric(nrow(data_raw) - nrow(data)) - SumExcluded
SumExcluded <- SumExcluded + NrExcluded
print(c("Excluded for low/irrational high BMI (and RFMp): ", NrExcluded))

data <- data[data$BMIz > -2,]
NrExcluded <- as.numeric(nrow(data_raw) - nrow(data)) - SumExcluded
SumExcluded <- SumExcluded + NrExcluded
print(c("Excluded for Underweight (BMI 'wasted'): ", NrExcluded))

data$birthcontrol[is.na(data$birthcontrol) == T] <- 0
data <- data[data$birthcontrol == 0,]
NrExcluded <- as.numeric(nrow(data_raw) - nrow(data)) - SumExcluded
SumExcluded <- SumExcluded + NrExcluded
print(c("Excluded for taking birth control: ", NrExcluded))

nrow(data) # 10.207 participants after 1st step of exclusion of cases

# separate twins (TEST) from rest (TRAIN)

data$famcount <- NA
data <- subset(data, is.na(family)==F)
nrow(data)
for (i in 1:nrow(data)) {
  famnr <- as.numeric(as.character(data[i,]$family))
  famcnt <- nrow(subset(data, family == famnr))
  data[i,]$famcount <- famcnt }

data <- subset(data, famcount < 3)

data$zygosity1[is.na(data$zygosity1)] <- 0
data$zygosity2[is.na(data$zygosity2)] <- 0
data$zygosity3[is.na(data$zygosity3)] <- 0
data$zygosity4[is.na(data$zygosity4)] <- 0

data$twinstatus <- "not processed"
for (i in 1:nrow(data)) {
  famnr <- as.numeric(as.character(data[i,]$family))
  if (data[i,]$famcount == 1) {
    data[i,]$twinstatus <- "singleton"
  } 
  else if (data[i,]$famcount == 2) { 
    if (subset(data, family == famnr)$zygosity1[1] == subset(data, family == famnr)$zygosity1[2]) { 
      if (data[i,]$zygosity1 == 1) { 
        data[i,]$twinstatus <- "MZ"
      } #close umatch
      else if (data[i,]$zygosity1 == 2) { 
        data[i,]$twinstatus <- "DZ"
      } 
      else if (data[i,]$zygosity1 == 3 | data[i,]$zygosity1 == 0) {
        data[i,]$twinstatus <- "sibling"
      }
    } 
    else if (subset(data, family == famnr)$zygosity1[1] != subset(data, family == famnr)$zygosity1[2]) { 
      data[i,]$twinstatus <- "unmatching pair"
    } 
  } 
} 

# mark possible errors, i.e. twin pairs w/o zygosity info 

table(data$twinstatus, data$siblings)
#data[data$twinstatus == "MZ" & data$siblings != 2,]$twinstatus <- "possible error" # no such case
data[data$twinstatus == "DZ" & data$siblings != 2,]$twinstatus <- "possible error"
data[data$twinstatus == "sibling" & data$siblings != 1,]$twinstatus <- "possible error"

data <- subset(data, select=c(ID:siblings, twinstatus, family:Sep_Calculations)) # move twin status to front of data set

data <- subset(data, twinstatus == "singleton" | twinstatus == "MZ" | twinstatus == "DZ" | twinstatus == "sibling")

NrExcluded <- as.numeric(nrow(data_raw) - nrow(data)) - SumExcluded
SumExcluded <- SumExcluded + NrExcluded
print(c("Excluded for inaccurate relatedness info: ", NrExcluded))

nrow(data) # TOAL: 9527 participants, after selecting cases for TRAIN and TEST sets
table(data$twinstatus)

## GET DEMOGRAPHIC INFO FOR REPORT
#save.image(file = "rsaves/data_for_demographics2.RData")
#load(file = "rsaves/data_for_demographics2.RData")
           
data$demgroup <- ifelse(data$twinstatus=="singleton", "singleton", "sib/twin")
table(data$demgroup)

table(subset(data, demgroup=="singleton")$infosource) #who is the informant
table(subset(data, demgroup=="singleton")$infosource)/nrow(subset(data, demgroup=="singleton"))
table(subset(data, demgroup=="sib/twin")$infosource)
table(subset(data, demgroup=="sib/twin")$infosource)/nrow(subset(data, demgroup=="sib/twin"))
# 1 = Childs Biological Mother; 2 = Childs Biological Father; 3 = Adoptive Parent; 
# 4 = Childs Custodial Parent; 5 = Other 

tapply(data$age, data$demgroup, mean) #age
tapply(data$age, data$demgroup, sd)
tapply(data$age, data$demgroup, range)

tapply(data$BMI95, data$demgroup, mean) #%BMI95
tapply(data$BMI95, data$demgroup, sd)
tapply(data$BMI95, data$demgroup, range)
nrow(subset(data, BMI95 >= 100 & demgroup=="singleton"))/nrow(subset(data, demgroup=="singleton")) #proportion of obese
nrow(subset(data, BMI95 >= 100 & demgroup=="sib/twin"))/nrow(subset(data, demgroup=="sib/twin"))

table(subset(data, demgroup=="singleton")$sex) #sex (0 male, 1 female, 2 div)
table(subset(data, demgroup=="singleton")$sex)/nrow(subset(data, demgroup=="singleton"))
table(subset(data, demgroup=="sib/twin")$sex)
table(subset(data, demgroup=="sib/twin")$sex)/nrow(subset(data, demgroup=="sib/twin"))

table(subset(data, demgroup=="singleton")$race) #race (0 white, 1 black/AfAm, 2 other/multiple)
table(subset(data, demgroup=="singleton")$race)/nrow(subset(data, demgroup=="singleton"))
table(subset(data, demgroup=="sib/twin")$race)
table(subset(data, demgroup=="sib/twin")$race)/nrow(subset(data, demgroup=="sib/twin"))

table(subset(data, demgroup=="singleton")$hispanic) #hispanic ethnicity (0 no, 1 yes)
table(subset(data, demgroup=="singleton")$hispanic)/nrow(subset(data, demgroup=="singleton"))
table(subset(data, demgroup=="sib/twin")$hispanic)
table(subset(data, demgroup=="sib/twin")$hispanic)/nrow(subset(data, demgroup=="sib/twin"))

table(subset(data, demgroup=="singleton")$income) #income
table(subset(data, demgroup=="singleton")$income)/nrow(subset(data, demgroup=="singleton"))
table(is.na(subset(data, demgroup=="singleton")$income))
table(is.na(subset(data, demgroup=="singleton")$income))/nrow(subset(data, demgroup=="singleton"))
table(subset(data, demgroup=="sib/twin")$income)
table(subset(data, demgroup=="sib/twin")$income)/nrow(subset(data, demgroup=="sib/twin"))
table(is.na(subset(data, demgroup=="sib/twin")$income))
table(is.na(subset(data, demgroup=="sib/twin")$income))/nrow(subset(data, demgroup=="sib/twin"))
# 1= Less than $5,000; 2=$5,000 through $11,999; 3=$12,000 through $15,999; 
# 4=$16,000 through $24,999; 5=$25,000 through $34,999; 6=$35,000 through $49,999; 
# 7=$50,000 through $74,999; 8= $75,000 through $99,999; 9=$100,000 through $199,999; 
# 10=$200,000 and greater

table(subset(data, demgroup=="singleton")$collegemat) #does mother have college degree 
table(subset(data, demgroup=="singleton")$collegemat)/nrow(subset(data, demgroup=="singleton"))
table(is.na(subset(data, demgroup=="singleton")$collegemat))
table(is.na(subset(data, demgroup=="singleton")$collegemat))/nrow(subset(data, demgroup=="singleton"))
table(subset(data, demgroup=="sib/twin")$collegemat)
table(subset(data, demgroup=="sib/twin")$collegemat)/nrow(subset(data, demgroup=="sib/twin"))
table(is.na(subset(data, demgroup=="sib/twin")$collegemat))
table(is.na(subset(data, demgroup=="sib/twin")$collegemat))/nrow(subset(data, demgroup=="sib/twin"))

table(subset(data, demgroup=="singleton")$collegepat) #does father have college degree 
table(subset(data, demgroup=="singleton")$collegepat)/nrow(subset(data, demgroup=="singleton"))
table(is.na(subset(data, demgroup=="singleton")$collegepat))
table(is.na(subset(data, demgroup=="singleton")$collegepat))/nrow(subset(data, demgroup=="singleton"))
table(subset(data, demgroup=="sib/twin")$collegepat)
table(subset(data, demgroup=="sib/twin")$collegepat)/nrow(subset(data, demgroup=="sib/twin"))
table(is.na(subset(data, demgroup=="sib/twin")$collegepat))
table(is.na(subset(data, demgroup=="sib/twin")$collegepat))/nrow(subset(data, demgroup=="sib/twin"))

table(subset(data, demgroup=="singleton")$singleParent) #single parent household 
table(subset(data, demgroup=="singleton")$singleParent)/nrow(subset(data, demgroup=="singleton"))
table(is.na(subset(data, demgroup=="singleton")$singleParent))
table(is.na(subset(data, demgroup=="singleton")$singleParent))/nrow(subset(data, demgroup=="singleton"))
table(subset(data, demgroup=="sib/twin")$singleParent)
table(subset(data, demgroup=="sib/twin")$singleParent)/nrow(subset(data, demgroup=="sib/twin"))
table(is.na(subset(data, demgroup=="sib/twin")$singleParent))
table(is.na(subset(data, demgroup=="sib/twin")$singleParent))/nrow(subset(data, demgroup=="sib/twin"))

table(subset(data, demgroup=="singleton")$handedness) #handedness (0 right, 1 left, 2 mixed)
table(subset(data, demgroup=="singleton")$handedness)/nrow(subset(data, demgroup=="singleton"))
table(is.na(subset(data, demgroup=="singleton")$handedness))
table(is.na(subset(data, demgroup=="singleton")$handedness))/nrow(subset(data, demgroup=="singleton"))
table(subset(data, demgroup=="sib/twin")$handedness)
table(subset(data, demgroup=="sib/twin")$handedness)/nrow(subset(data, demgroup=="sib/twin"))
table(is.na(subset(data, demgroup=="sib/twin")$handedness))
table(is.na(subset(data, demgroup=="sib/twin")$handedness))/nrow(subset(data, demgroup=="sib/twin"))

# Tidy up Factor variables, use largest group as reference

data$sex <- as.factor(data$sex)
data$sex <- relevel(data$sex, ref = "0")
data$race <- as.factor(data$race)
data$race <- relevel(data$race, ref = "0")
data$hispanic <- as.factor(data$hispanic)
data$hispanic <- relevel(data$hispanic, ref = "0")
data$handedness <- as.factor(data$handedness) 
data$handedness <- relevel(data$handedness, ref = "1")
data$site <- as.factor(data$site)
data$site <- relevel(data$site, ref = "site16")

# Delete MRI data, that doesn't pass quality control

data$mriqc <- ifelse(data$mriqc==1, 1, 0)
table(data$mriqc) # 434 scans fail quality control at baseline
tempdat1 <- subset(data, mriqc==1)
tempdat2 <- subset(data, mriqc!=1)
tempdat2[, which(colnames(tempdat2)=="Sep_CT") : which(colnames(tempdat2)=="Sep_PersItems")] <- NA
data <- rbind(tempdat1, tempdat2)

# GET CORRELATIONS OF CONTROLS WITH BMI

controlsfit <- lm(scale(BMI95) ~ sex*scale(puberty) + scale(age) + race + hispanic + handedness
                  + scale(meanthickness) + scale(totalarea) + scale(totalvolume), data = data)
controlsout <- summary(controlsfit)
controlscoeff <- data.frame(controlsout$coefficients)
controlscoeff <- controlscoeff[-1,]
controlscoeff$control <- row.names(controlscoeff)
controlscoeff$label <- c("Gender: Female", "Gender: Diverse", "Puberty", "Age",
                         "Race: Black/African American", "Race: Other/Multiple",
                         "Ethnicity: Hispanic/Latino", "Handedness: Left", "Handedness: Mixed",
                         "Mean Cortical Thickness", "Total Cortical Surface Area", "Total Cortical Volume",
                         "Puberty x Female Gender", "Puberty x Diverse Gender")
controlscoeff$label <- factor(controlscoeff$label, levels=unique(controlscoeff$label))
controlscoeff$lower <- controlscoeff$Estimate - controlscoeff$Std..Error /2
controlscoeff$upper <- controlscoeff$Estimate + controlscoeff$Std..Error /2
controlscoeff$p_FDR <- p.adjust(controlscoeff[,4], method = "fdr")
controlscoeff$Significance <- ifelse(controlscoeff$p_FDR < .05, "FDR P < 0.05", "n.s.")

theme_set(theme_bw())

ggplot(controlscoeff, aes(x=label, y=Estimate, color=Significance)) +
  geom_errorbar(aes(ymin=lower, ymax=upper), size=1.2, width=0) +
  geom_point(size=3) +
  ylim(-3.5, 3.5) +
  scale_x_discrete(name="", limits=rev) +
  coord_flip() +
  geom_hline(yintercept=0, linetype="dashed") +
  scale_color_manual(breaks = c("FDR P < 0.05", "n.s."), values=c("grey47", "grey75")) +
  ylab("Association with Obesity") +
  theme(text = element_text(size=14))

## RESIDUALISE

# model 1 for personality and cognition
mod1 <- data[,i] ~ data$sex*data$puberty + data$age + data$race + data$hispanic + data$handedness + data$site
# model 2 for cortical thickness
mod2 <- data[,i] ~ data$sex*data$puberty + data$age + data$race + data$hispanic + data$handedness + data$site + data$meanthickness
# model 3 for cortical area
mod3 <- data[,i] ~ data$sex*data$puberty + data$age + data$race + data$hispanic + data$handedness + data$site + data$totalarea
# model 4 for cortical volume
mod4 <- data[,i] ~ data$sex*data$puberty + data$age + data$race + data$hispanic + data$handedness + data$site + data$totalvolume

# other potential controls: data$married, data$singleParent, data$stimulants, data$income, data$collgemat, data$collgepat, data$sex_menstruation

for (i in (which(colnames(data)=="Sep_Obesity")+1) : (which(colnames(data)=="Sep_PersScales")-1)) {
  data[,i] <- residuals(lm(mod1, na.action = na.exclude)) }

for (i in (which(colnames(data)=="Sep_PersScales")+1) : (which(colnames(data)=="Sep_Cognition")-1)) {
  data[,i] <- residuals(lm(mod1, na.action = na.exclude)) }

for (i in (which(colnames(data)=="Sep_Cognition")+1) : (which(colnames(data)=="Sep_CT")-1)) {
  data[,i] <- residuals(lm(mod1, na.action = na.exclude)) }

for (i in (which(colnames(data)=="Sep_CT")+1) : (which(colnames(data)=="Sep_CA")-1)) {
  data[,i] <- residuals(lm(mod2, na.action = na.exclude)) }

for (i in (which(colnames(data)=="Sep_CA")+1) : (which(colnames(data)=="Sep_CV")-1)) {
  data[,i] <- residuals(lm(mod3, na.action = na.exclude)) }

for (i in (which(colnames(data)=="Sep_CV")+1) : (which(colnames(data)=="Sep_PersItems")-1)) {
  data[,i] <- residuals(lm(mod4, na.action = na.exclude)) }

for (i in (which(colnames(data)=="Sep_PersItems")+1) : (which(colnames(data)=="Sep_Eating")-1)) {
  data[,i] <- residuals(lm(mod1, na.action = na.exclude)) }

for (i in (which(colnames(data)=="Sep_Eating")+1) : (which(colnames(data)=="Sep_Calculations")-1)) {
  data[,i] <- residuals(lm(mod1, na.action = na.exclude)) }

## SAVE FOR LATER ANALYSIS STEPS
save(data, file="rsaves/data-BL-clean_2022-02-18.Rda")
write.csv(data, file="rsaves/data-BL-clean_2022-02-18.csv", row.names = FALSE)
#load(file="rsaves/data-BL-clean_2022-02-18.Rda")

## SELECT ONLY SINGLETONS
table(data$twinstatus)
data <- data[data$twinstatus == "singleton",]

nrow(data) # 6976 participants after selecting only "singletons" (only child per family in data)

# BMI skew test

skew(data$BMI95) # 1.2 - skew is okay

## CORRELATION

feature <- colnames(data)
cat <- c(rep('other', which(colnames(data)=="Sep_PersScales")-1), 
         rep('personality_scales', which(colnames(data)=="Sep_Cognition") - which(colnames(data)=="Sep_PersScales")), 
         rep('cognition', which(colnames(data)=="Sep_CT") - which(colnames(data)=="Sep_Cognition")), 
         rep('corticalthickness', which(colnames(data)=="Sep_CA") - which(colnames(data)=="Sep_CT")), 
         rep('corticalarea', which(colnames(data)=="Sep_CV") - which(colnames(data)=="Sep_CA")), 
         rep('corticalvolume', which(colnames(data)=="Sep_PersItems") - which(colnames(data)=="Sep_CV")), 
         rep('personality_items', which(colnames(data)=="Sep_Eating") - which(colnames(data)=="Sep_PersItems")), 
         rep('eating_items', which(colnames(data)=="Sep_Calculations") - which(colnames(data)=="Sep_Eating")),
         rep ('other', 2))

# BMI
BMIcorr <- data.frame(feature, cat)
BMIcorr$r <- NA
BMIcorr$ci_lower <- NA
BMIcorr$ci_upper <- NA
BMIcorr$p <- NA
for (i in (which(colnames(data)=="RFMp")+1) : (ncol(data))) {
  BMIcorr$r[i] <- corr.test(data$BMI95, data[,i])$ci[,2]
  BMIcorr$ci_lower[i] <- corr.test(data$BMI95, data[,i])$ci[,1]
  BMIcorr$ci_upper[i] <- corr.test(data$BMI95, data[,i])$ci[,3]
  BMIcorr$p[i] <- corr.test(data$BMI95, data[,i])$ci[,4] }
factors <- levels(BMIcorr$cat)
BMIcorrList <- list()
for (i in 1:length(factors)) {
  if (factors[i] != "other") {
    BMIcorrList[[i]] <- BMIcorr[BMIcorr$cat==factors[i],]
    BMIcorrList[[i]] <- BMIcorrList[[i]][-1,]
    BMIcorrList[[i]]$p_FDR <- p.adjust( BMIcorrList[[i]]$p, method = "fdr")
    BMIcorrList[[i]]$Significance <- ifelse( BMIcorrList[[i]]$p_FDR < 0.05, "FDR P < 0.05", "n.s.")
  }
}
BMIcorrPersScales <- BMIcorrList[[8]]
BMIcorrCog <- BMIcorrList[[1]]
BMIcorrCT <- BMIcorrList[[3]]
BMIcorrCA <- BMIcorrList[[2]]
BMIcorrCV <- BMIcorrList[[4]]
BMIcorrPersItems <- BMIcorrList[[7]]
BMIcorrEatItems <- BMIcorrList[[5]]

# # RFM --> RFM was explored as additional indicator but dropped because of strong similarity with %BMIp95
# RFMcorr <- data.frame(feature, cat)
# RFMcorr$r <- NA
# RFMcorr$ci_lower <- NA
# RFMcorr$ci_upper <- NA
# RFMcorr$p <- NA
# for (i in (which(colnames(data)=="RFMp")+1) : (ncol(data))) {
#   RFMcorr$r[i] <- corr.test(data$RFMp, data[,i])$ci[,2]
#   RFMcorr$ci_lower[i] <- corr.test(data$RFMp, data[,i])$ci[,1]
#   RFMcorr$ci_upper[i] <- corr.test(data$RFMp, data[,i])$ci[,3]
#   RFMcorr$p[i] <- corr.test(data$RFMp, data[,i])$ci[,4] }
# RFMcorrPers <- RFMcorr[RFMcorr$cat=='personality',]
# RFMcorrPers$p_FDR <- p.adjust(RFMcorrPers$p, method = "fdr")
# RFMcorrPers$Significance <- ifelse(RFMcorrPers$p_FDR < 0.05, "FDR P < 0.05", "n.s.")
# RFMcorrCog <- RFMcorr[RFMcorr$cat=='cognition',]
# RFMcorrCog$p_FDR <- p.adjust(RFMcorrCog$p, method = "fdr")
# RFMcorrCog$Significance <- ifelse(RFMcorrCog$p_FDR < 0.05, "FDR P < 0.05", "n.s.")
# RFMcorrCT <- RFMcorr[RFMcorr$cat=='corticalthickness',]
# RFMcorrCT$p_FDR <- p.adjust(RFMcorrCT$p, method = "fdr")
# RFMcorrCT$Significance <- ifelse(RFMcorrCT$p_FDR < 0.05, "FDR P < 0.05", "n.s.")
# RFMcorrPersItems <- RFMcorr[RFMcorr$cat=='personality_items',]
# RFMcorrPersItems$p_FDR <- p.adjust(RFMcorrPersItems$p, method = "fdr")
# RFMcorrPersItems$Significance <- ifelse(RFMcorrPersItems$p_FDR < 0.05, "FDR P < 0.05", "n.s.")
# RFMcorrEatItems <- RFMcorr[RFMcorr$cat=='eating_items',]
# RFMcorrEatItems$p_FDR <- p.adjust(RFMcorrEatItems$p, method = "fdr")
# RFMcorrEatItems$Significance <- ifelse(RFMcorrEatItems$p_FDR < 0.05, "FDR P < 0.05", "n.s.")

## PLOTS

perslabels <- c("Negative Urgency", 
                       "Lack of Planning", 
                       "Sensation Seeking", 
                       "Positive Urgency", 
                       "Lack of Perseverance", 
                       "Inhibition", 
                       "Reward Responsiveness", 
                       "Drive", 
                       "Anxious/Depressed", 
                       "Depressed", 
                       "Somatic Complaints", 
                       "Social Problems", 
                       "Thought Problems", 
                       "Attention Problems", 
                       "Rule-Breaking Behavior", 
                       "Agressive Behavior", 
                       "Prosociality")

coglabels <- c("Picture Vocabulary", 
                      "Inhibitory Control", 
                      "List Sorting", 
                      "Card Sorting", 
                      "Pattern Comparison", 
                      "Picture Sequence", 
                      "Reading Recognition", 
                      "Learning", 
                      "Short-term", 
                      "Long-term", 
                      "Matrix Reasoning", 
                      "Accuracy", 
                      "Speed")

brainlabels <- c("Banks of Superior Temporal Sulcus", 
                     "Caudal anterior cingulate", 
                     "Caudal middle frontal",
                     "Cuneus",
                     "Entorhinal",
                     "Fusiform",
                     "Inferior parietal",
                     "Inferior temporal",
                     "Isthmus cingulate",
                     "Lateral occipital",
                     "Lateral orbitofrontal",
                     "Lingual",
                     "Medial orbitofrontal",
                     "Middle temporal",
                     "Parahippocampal",
                     "Paracentral",
                     "Pars opercularis",
                     "Pars orbitalis",
                     "Pars triangularis",
                     "Pericalcarine",
                     "Postcentral",
                     "Posterior cingulate",
                     "Precentral",
                     "Precuneus",
                     "Rostral anterior cingulate",
                     "Rostral middle frontal",
                     "Superior frontal",
                     "Superior parietal",
                     "Superior temporal",
                     "Supramarginal",
                     "Frontal pole",
                     "Temporal pole",
                     "Transverse temporal",
                     "Insula")

brainlobes <- c("Temporal", 
                 "Frontal", 
                 "Frontal",
                 "Parietal",
                 "Temporal",
                 "Temporal",
                 "Parietal",
                 "Temporal",
                 "Frontal",
                 "Occipital",
                 "Frontal",
                 "Occipital",
                 "Frontal",
                 "Temporal",
                 "Temporal",
                 "Parietal",
                 "Frontal",
                 "Frontal",
                 "Frontal",
                 "Occipital",
                 "Parietal",
                 "Parietal",
                 "Frontal",
                 "Parietal",
                 "Frontal",
                 "Frontal",
                 "Frontal",
                 "Parietal",
                 "Temporal",
                 "Parietal",
                 "Frontal",
                 "Temporal",
                 "Temporal",
                 "I")

theme_set(theme_bw())

# BMI

# for personality scales

BMIcorrPersScales$label <- perslabels
BMIcorrPersScales$vargroup <- c(rep("Impulsivity",5), rep("Motivation",3), rep("Psychopathology",8), "PS")

ggplot(BMIcorrPersScales, aes(x=reorder(label, -(r)), y=r, color=Significance)) +
  geom_errorbar(aes(ymin=ci_lower, ymax=ci_upper), size=1.2, width=0) +
  geom_point(size=3) +
  ylim(-0.1, 0.1) +
  scale_x_discrete(name="", limits=rev) +
  facet_grid(rows = vars(vargroup), scales='free', space = "free") +
  coord_flip() +
  geom_hline(yintercept=0, linetype="dashed") +
  scale_color_manual(breaks = c("FDR P < 0.05", "n.s."), values=c("grey47", "grey75")) +
  #ggtitle("Obesity-Personality Correlations") +
  theme(text = element_text(size=14), legend.position="bottom", legend.title = element_blank())

# for cognition

BMIcorrCog$label <- coglabels
BMIcorrCog$vargroup <- c("Lang","EF","WM", "EF",
                                      "WM","Memory","Lang","Memory","Memory","Memory","EF", 
                                      "Perc", "Perc")

cogplot <- ggplot(BMIcorrCog, aes(x=reorder(label, -(r)), y=r, color=Significance)) + 
  geom_errorbar(aes(ymin=ci_lower, ymax=ci_upper), size=1.2, width=0) +
  geom_point(size=3) +
  ylim(-0.1, 0.1) +
  scale_x_discrete(name="", limits=rev) +
  facet_grid(rows = vars(vargroup), scales='free', space = "free") +
  coord_flip() +
  geom_hline(yintercept=0, linetype="dashed") +
  scale_color_manual(breaks = c("FDR P < 0.05", "n.s."), values=c("grey47", "grey75")) +
  #ggtitle("Obesity-Cognition Correlations")
  theme(text = element_text(size=14), legend.position="none", legend.title = element_blank())
cogplot

# for cortical thickness

BMIcorrCT$Hemisphere <- c(rep('left', 34), rep('right', 34))
CTcopy <- BMIcorrCT[1:34,]$feature
BMIcorrCT$feature <- rep(CTcopy, 2)

BMIcorrCT$label <- brainlabels
BMIcorrCT$label <- factor(BMIcorrCT$label, levels=unique(BMIcorrCT$label))
BMIcorrCT$vargroup <- brainlobes

ctplot <- ggplot(BMIcorrCT, aes(x=label, y=r, group=Hemisphere, color=Significance)) + 
  geom_errorbar(aes(ymin=ci_lower, ymax=ci_upper),  size=1.2, width=0, position = position_dodge(width = 0.5)) +
  geom_point(size=3, aes(shape=Hemisphere), position = position_dodge(width = 0.5)) +
  #ylim(-0.15, 0.15) +
  scale_x_discrete(name="", limits=rev) +
  scale_y_continuous(limits = c(-0.15, 0.15), breaks = c(-.15,-.1,-.05,0,.05,.1,.05,.1,.15)) +
  facet_grid(rows = vars(vargroup), scales='free', space = "free") +
  coord_flip() +
  geom_hline(yintercept=0, linetype="dashed") +
  scale_color_manual(breaks = c("FDR P < 0.05", "n.s."), values=c("grey47", "grey75")) +
  #ggtitle("Obesity-Cortical Thickness Correlations")
  theme(text = element_text(size=14), legend.title = element_blank(),
        legend.position='none')
ctplot

# Make data for 2D brain plot
r_right <- subset(BMIcorrCT, Hemisphere=="right")$r 
r_right <- c(r_right[1:3], NA, r_right[4:34])
r_right <- data.frame(r= r_right, id= 2:36, hemi="right")
r_left <- subset(BMIcorrCT, Hemisphere=="left")$r
r_left <- c(r_left[1:3], NA, r_left[4:34])
r_left <- data.frame(r= r_left, id= 2:36, hemi="left")
rhData2d <- dk$data
rhData2d$id <- as.numeric(rhData2d$roi)
rhData2d <- merge(rhData2d, r_right, by=c("id","hemi"), all.x=T, all.y=F)
rhData2d <- merge(rhData2d, r_left, by=c("id","hemi"), all.x=T, all.y=F)
rhData2d$r <- ifelse(is.na(rhData2d$r.x)==F, rhData2d$r.x, rhData2d$r.y)
rhData2d$atlas <- "dk"
rhData2d$type <- "cortical"

# Generate 2D plot
brainCT <- ggseg(.data = rhData2d, 
      mapping=aes(fill=r),
      colour = "black",
      adapt_scales = T,
      position = "dispersed",
      show.legend = F) +
  scale_fill_gradient2(low = "darkred", mid = "white", high = "darkblue", midpoint = 0, na.value = "lightgrey", limits = c(-.15,.15)) +
  theme_classic() +
  theme(text = element_text(size=14), axis.title.x=element_blank(), axis.text.x=element_blank(),) +
  labs(title = "A")

# for cortical area

BMIcorrCA$Hemisphere <- c(rep('left', 34), rep('right', 34))
CAcopy <- BMIcorrCA[1:34,]$feature
BMIcorrCA$feature <- rep(CAcopy, 2)

BMIcorrCA$label <- brainlabels
BMIcorrCA$label <- factor(BMIcorrCA$label, levels=unique(BMIcorrCA$label))
BMIcorrCA$vargroup <- brainlobes

caplot <- ggplot(BMIcorrCA, aes(x=label, y=r, group=Hemisphere, color=Significance)) + 
  geom_errorbar(aes(ymin=ci_lower, ymax=ci_upper),  size=1.2, width=0, position = position_dodge(width = 0.5)) +
  geom_point(size=3, aes(shape=Hemisphere), position = position_dodge(width = 0.5)) +
  #ylim(-0.15, 0.15) +
  scale_x_discrete(name="", limits=rev) +
  scale_y_continuous(limits = c(-0.15, 0.15), breaks = c(-.15,-.1,-.05,0,.05,.1,.05,.1,.15)) +
  facet_grid(rows = vars(vargroup), scales='free', space = "free") +
  coord_flip() +
  geom_hline(yintercept=0, linetype="dashed") +
  scale_color_manual(breaks = c("FDR P < 0.05", "n.s."), values=c("grey47", "grey75")) +
  #ggtitle("Obesity-Cortical Surface Area Correlations")
  theme(text = element_text(size=14), legend.position="bottom", legend.title = element_blank(), 
        axis.text.y=element_blank(), axis.ticks.y=element_blank())
ctplot + caplot

# Make data for 2D brain plot
r_right <- subset(BMIcorrCA, Hemisphere=="right")$r 
r_right <- c(r_right[1:3], NA, r_right[4:34])
r_right <- data.frame(r= r_right, id= 2:36, hemi="right")
r_left <- subset(BMIcorrCA, Hemisphere=="left")$r
r_left <- c(r_left[1:3], NA, r_left[4:34])
r_left <- data.frame(r= r_left, id= 2:36, hemi="left")
rhData2d <- dk$data
rhData2d$id <- as.numeric(rhData2d$roi)
rhData2d <- merge(rhData2d, r_right, by=c("id","hemi"), all.x=T, all.y=F)
rhData2d <- merge(rhData2d, r_left, by=c("id","hemi"), all.x=T, all.y=F)
rhData2d$r <- ifelse(is.na(rhData2d$r.x)==F, rhData2d$r.x, rhData2d$r.y)
rhData2d$atlas <- "dk"
rhData2d$type <- "cortical"

# Generate 2D plot
brainCA <- ggseg(.data = rhData2d, 
            mapping=aes(fill=r),
            colour = "black",
            adapt_scales = T,
            position = "dispersed",
            show.legend = F) +
  scale_fill_gradient2(low = "darkred", mid = "white", high = "darkblue", midpoint = 0, na.value = "lightgrey", limits = c(-.15,.15)) +
  theme_classic() + 
  theme(text = element_text(size=14), axis.title.x=element_blank(), axis.text.x=element_blank(),) +
  labs(title = "B")

# for cortical volume

BMIcorrCV$Hemisphere <- c(rep('left', 34), rep('right', 34))
CAcopy <- BMIcorrCV[1:34,]$feature
BMIcorrCV$feature <- rep(CAcopy, 2)

BMIcorrCV$label <- brainlabels
BMIcorrCV$label <- factor(BMIcorrCV$label, levels=unique(BMIcorrCV$label))
BMIcorrCV$vargroup <- brainlobes

cvplot <- ggplot(BMIcorrCV, aes(x=label, y=r, group=Hemisphere, color=Significance)) + 
  geom_errorbar(aes(ymin=ci_lower, ymax=ci_upper),  size=1.2, width=0, position = position_dodge(width = 0.5)) +
  geom_point(size=3, aes(shape=Hemisphere), position = position_dodge(width = 0.5)) +
  #ylim(-0.15, 0.15) +
  scale_x_discrete(name="", limits=rev) +
  scale_y_continuous(limits = c(-0.15, 0.15), breaks = c(-.15,-.1,-.05,0,.05,.1,.05,.1,.15)) +
  facet_grid(rows = vars(vargroup), scales='free', space = "free") +
  coord_flip() +
  geom_hline(yintercept=0, linetype="dashed") +
  scale_color_manual(breaks = c("FDR P < 0.05", "n.s."), values=c("grey47", "grey75")) +
  #ggtitle("Obesity-Cortical Volume Correlations")
  theme(text = element_text(size=14), legend.title = element_blank(), axis.text.y=element_blank(),
        axis.ticks.y=element_blank(), legend.position='none')
ctplot + caplot + cvplot

# Make data for 2D brain plot
r_right <- subset(BMIcorrCV, Hemisphere=="right")$r 
r_right <- c(r_right[1:3], NA, r_right[4:34])
r_right <- data.frame(r= r_right, id= 2:36, hemi="right")
r_left <- subset(BMIcorrCV, Hemisphere=="left")$r
r_left <- c(r_left[1:3], NA, r_left[4:34])
r_left <- data.frame(r= r_left, id= 2:36, hemi="left")
rhData2d <- dk$data
rhData2d$id <- as.numeric(rhData2d$roi)
rhData2d <- merge(rhData2d, r_right, by=c("id","hemi"), all.x=T, all.y=F)
rhData2d <- merge(rhData2d, r_left, by=c("id","hemi"), all.x=T, all.y=F)
rhData2d$r <- ifelse(is.na(rhData2d$r.x)==F, rhData2d$r.x, rhData2d$r.y)
rhData2d$atlas <- "dk"
rhData2d$type <- "cortical"

# Generate 2D plot
brainCV <- ggseg(.data = rhData2d, 
      mapping=aes(fill=r),
      colour = "black",
      adapt_scales = T,
      position = "dispersed",
      show.legend = T) +
  scale_fill_gradient2(low = "darkred", mid = "white", high = "darkblue", midpoint = 0, na.value = "lightgrey", limits = c(-.15,.15)) +
  theme_classic() +
  theme(text = element_text(size=14), legend.position = "bottom", legend.text = element_text(size = 14)) + 
  guides(fill = guide_colourbar(barwidth = 25, barheight = .5)) +
  labs(title = "C")

# combine all 3 brain plots
ggarrange(brainCT, brainCA, brainCV, ncol = 1, nrow = 3, heights = c(1,1,1.5))
ggarrange(ctplot, caplot, cvplot, labels = c("A", "B", "C"), ncol = 3, nrow = 1, common.legend = T, 
          legend="bottom", widths = c(1.75,1,1))

# for eating items

BMIcorrEatItems$label <- c("Doesn't Eat Well", "Overreating")
BMIcorrEatItems$vargroup <- rep("EB",2)

ggplot(BMIcorrEatItems, aes(x=reorder(label, -(r)), y=r, color=Significance)) +
  geom_errorbar(aes(ymin=ci_lower, ymax=ci_upper), size=1.2, width=0) +
  geom_point(size=3) +
  ylim(-0.5, 0.5) +
  scale_x_discrete(name="", limits=rev) +
  facet_grid(rows = vars(vargroup), scales='free', space = "free") +
  coord_flip() +
  geom_hline(yintercept=0, linetype="dashed") +
  scale_color_manual(breaks = c("FDR P < 0.05", "n.s."), values=c("grey47", "grey75")) +
  #ggtitle("Obesity-Eating Correlations") +
  theme(text = element_text(size=14), legend.position="bottom", legend.title = element_blank())

# for both behavioral factors


BMIcorrBehav <- rbind(BMIcorrPersScales, BMIcorrEatItems)
BMIcorrBehav$vargroup_f = factor(BMIcorrBehav$vargroup, levels=c('Impulsivity','Motivation',
                                                                 'PS','Psychopathology', 'EB'))

behavplot <- ggplot(BMIcorrBehav, aes(x=reorder(label, -(r)), y=r, color=Significance)) +
  geom_errorbar(aes(ymin=ci_lower, ymax=ci_upper), size=1.2, width=0) +
  geom_point(size=3) +
  scale_y_continuous(limits = c(-0.45, 0.45), breaks = c(-.4,-.3,-.2,-.1,0,.1,.2,.3,.4)) +
  scale_x_discrete(name="", limits=rev) +
  facet_grid(rows = vars(vargroup_f), scales='free', space = "free") +
  coord_flip() +
  geom_hline(yintercept=0, linetype="dashed") +
  scale_color_manual(breaks = c("FDR P < 0.05", "n.s."), values=c("grey47", "grey75")) +
  #ggtitle("B") +
  theme(text = element_text(size=14), legend.position="bottom", legend.title = element_blank())
behavplot

# combine behavior + cognition plots
ggarrange(behavplot, cogplot, labels = c("A", "B"), ncol = 1, nrow = 2, heights = c(1.45,1),
          common.legend = T, legend="bottom")

# for personality at item level

ggplot(BMIcorrPersItems, aes(x=reorder(feature, r), y=r, color=Significance)) + 
  geom_errorbar(aes(ymin=ci_lower, ymax=ci_upper), size=.5, width=0) +
  geom_point(size=1.0) +
  ylim(-0.1, 0.2) +
  scale_x_discrete(name="", limits=rev) +
  theme(axis.text.y=element_text(size=rel(0.7))) +
  coord_flip() +
  geom_hline(yintercept=0, linetype="dashed") +
  scale_color_manual(breaks = c("FDR P < 0.05", "n.s."), values=c("grey47", "grey75")) +
  ggtitle("Obesity-Personality Correlations at Item Level")

# # RFM --> RFM was explored as additional indicator but dropped because of strong similarity with %BMIp95
# 
# # for personality
# 
# RFMcorrPers$label <- perslabels
# 
# ggplot(RFMcorrPers, aes(x=reorder(label, label), y=r, color=Significance)) + 
#   geom_errorbar(aes(ymin=ci_lower, ymax=ci_upper), size=1.2, width=0) +
#   geom_point(size=3) +
#   ylim(-0.2, 0.2) +
#   scale_x_discrete(name="", limits=rev) +
#   coord_flip() +
#   geom_hline(yintercept=0, linetype="dashed") +
#   scale_color_manual(breaks = c("FDR P < 0.05", "n.s."), values=c("black", "grey")) +
#   ggtitle("RFM-Personality Correlations")
# 
# # for cognition
# 
# RFMcorrCog$label <- coglabels
# 
# ggplot(RFMcorrCog, aes(x=reorder(label, label), y=r, color=Significance)) + 
#   geom_errorbar(aes(ymin=ci_lower, ymax=ci_upper), size=1.2, width=0) +
#   geom_point(size=3) +
#   ylim(-0.2, 0.2) +
#   scale_x_discrete(name="", limits=rev) +
#   coord_flip() +
#   geom_hline(yintercept=0, linetype="dashed") +
#   scale_color_manual(breaks = c("FDR P < 0.05", "n.s."), values=c("black", "grey")) +
#   ggtitle("RFM-Cognition Correlations")
# 
# # for cortical thickness
# 
# RFMcorrCT$Hemisphere <- c(rep('left', 34), rep('right', 34))
# CTcopy <- RFMcorrCT[1:34,]$feature
# RFMcorrCT$feature <- rep(CTcopy, 2)
# 
# RFMcorrCT$label <- ctlabels
# RFMcorrCT$label <- factor(RFMcorrCT$label, levels=unique(RFMcorrCT$label))
# 
# ggplot(RFMcorrCT, aes(x=label, y=r, group=Hemisphere, color=Significance)) + 
#   geom_errorbar(aes(ymin=ci_lower, ymax=ci_upper),  size=1.2, width=0) +
#   geom_point(size=3, aes(shape=Hemisphere)) +
#   ylim(-0.2, 0.2) +
#   scale_x_discrete(name="", limits=rev) +
#   coord_flip() +
#   geom_hline(yintercept=0, linetype="dashed") +
#   scale_color_manual(breaks = c("FDR P < 0.05", "n.s."), values=c("black", "grey")) +
#   ggtitle("RFM-Cortical Thickness Correlations")

### SAVE CORRELATION DATA FRAMES
save(BMIcorr, file="rsaves/BMI-Correlations_2022-03-29.Rda")
write.csv(BMIcorr, file="rsaves/BMI-Correlations_2022-03-29.csv", row.names = FALSE)
# save(RFMcorr, file="rsaves/RFM-Correlations_2021-09-21.Rda")
# write.csv(RFMcorr, file="rsaves/RFM-Correlations_2021-09-21.csv", row.names = FALSE)
rm(list = ls())

############################################################################

#########################
#
# Part III: PPS IN SINGLETONS
#
######################### ----

rm(list = ls())

## "SWITCH BOARD"
#-

## PREPARATION

# set wd
setwd("~/Job/Tartu Traineeship/R Analysis/abcd_bmi")

# packages
library(psych)
library(ggplot2)
library(glmnet)
library(cytominer)

# read files
data <- read.csv("rsaves/data-BL-clean_2022-02-18.csv", sep=",", dec=".", header=TRUE) # read cleaned Baseline
data_raw <- data

## CALCULATE all types PPS (correlation, lasso a=.01, lasso a=.1, lasso a=.5, lasso a=1)

set.seed(777)

data <- data[data$twinstatus == "singleton",] 
data <- data[,which(colnames(data)=="BMI95"):ncol(data)]
data <- data[,-c(which(colnames(data)=="Sep_PersScales"),
                 which(colnames(data)=="Sep_Cognition"),
                 which(colnames(data)=="Sep_CT"),
                 which(colnames(data)=="Sep_CA"),
                 which(colnames(data)=="Sep_CV"),
                 which(colnames(data)=="Sep_PersItems"),
                 which(colnames(data)=="Sep_Eating"),
                 which(colnames(data)=="Sep_Calculations"))]
data <- na.omit(data)
nrow(data) # 6.347 singeltons w/o missing values

#build structures
y_bmi <- data[, which(colnames(data)=="BMI95")]
#y_rfmp <- data[, which(colnames(data)=="RFMp")]
x_pers <- as.matrix(data[, (which(colnames(data)=="upps_negurgency")) : (which(colnames(data)=="prosociality"))])
x_cog <- as.matrix(data[, (which(colnames(data)=="nih_picvoc")) : (which(colnames(data)=="lmt_RT"))])
x_ct <- as.matrix(data[, (which(colnames(data)=="smri_thick_cdk_banksstslh")) : (which(colnames(data)=="smri_thick_cdk_insularh"))])
x_ca <- as.matrix(data[, (which(colnames(data)=="smri_area_cdk_banksstslh")) : (which(colnames(data)=="smri_area_cdk_insularh"))])
x_cv <- as.matrix(data[, (which(colnames(data)=="smri_vol_cdk_banksstslh")) : (which(colnames(data)=="smri_vol_cdk_insularh"))])
x_persitems <- as.matrix(data[, (which(colnames(data)=="upps6_y")) : (which(colnames(data)=="prosocial_q3_y"))])
x_eatitems <- as.matrix(data[, (which(colnames(data)=="cbcl_q24_p")) : (which(colnames(data)=="cbcl_q53_p"))])

# indentify highly intercorrelating items
#x_persitems2 <- data[, (which(colnames(data)=="upps6_y")) : (which(colnames(data)=="prosocial_q3_y"))]
#correlation_threshold(variables = names(x_persitems2), sample = x_persitems2, cutoff = 0.7)
#hist(cor(x_persitems2)[cor(x_persitems2)<1])
#hist(cor(x_ct)[cor(x_ct)<1])

x_list <- list()
x_list[[1]] <- x_pers
x_list[[2]] <- x_cog
x_list[[3]] <- x_ct
x_list[[4]] <- x_ca
x_list[[5]] <- x_cv
x_list[[6]] <- x_persitems
x_list[[7]] <- x_eatitems

data$PPS_Pers_Cor <- NA # new cols
data$PPS_Pers_L001 <- NA
data$PPS_Pers_L010 <- NA
data$PPS_Pers_L050 <- NA
data$PPS_Pers_L100 <- NA

data$PPS_Cog_Cor <- NA
data$PPS_Cog_L001 <- NA
data$PPS_Cog_L010 <- NA
data$PPS_Cog_L050 <- NA
data$PPS_Cog_L100 <- NA

data$PPS_CT_Cor <- NA
data$PPS_CT_L001 <- NA
data$PPS_CT_L010 <- NA
data$PPS_CT_L050 <- NA
data$PPS_CT_L100 <- NA

data$PPS_CA_Cor <- NA
data$PPS_CA_L001 <- NA
data$PPS_CA_L010 <- NA
data$PPS_CA_L050 <- NA
data$PPS_CA_L100 <- NA

data$PPS_CV_Cor <- NA
data$PPS_CV_L001 <- NA
data$PPS_CV_L010 <- NA
data$PPS_CV_L050 <- NA
data$PPS_CV_L100 <- NA

data$PPS_PersItems_Cor <- NA
data$PPS_PersItems_L001 <- NA
data$PPS_PersItems_L010 <- NA
data$PPS_PersItems_L050 <- NA
data$PPS_PersItems_L100 <- NA

data$PPS_EatItems_Cor <- NA
data$PPS_EatItems_L001 <- NA
data$PPS_EatItems_L010 <- NA
data$PPS_EatItems_L050 <- NA
data$PPS_EatItems_L100 <- NA

nfolds <- 20

folds <- sample(1:nfolds, nrow(data),replace=T)
table(folds)

catname <- c('personality_scales', 'cognition', 'corticalthickness', 'corticalarea', 'corticalvolume', 'personality_items', 'eating_items')
method <- c(999, 0.01, 0.1, 0.5, 1)
fits <- list()

for (i3 in 1:length(catname)) {
  print(catname[i3])
  for (i2 in 1:length(method)) {
    print(method[i2])
    if (method[i2] != 999) {
      print("lasso PPS")
      for (i1 in 1:nfolds) {
        print(i1)
        rowstoinclude <- folds != i1
        subx <- x_list[[i3]]
        subx <- subx[rowstoinclude,]
        suby <- y_bmi[rowstoinclude]
        fits[[i1]] <- cv.glmnet(subx, suby, alpha = method[i2], nfolds=10)
      }
      PPS <- rep(NA, nrow(data))
      for ( i1 in 1:nfolds) { 
        weights <- coef(fits[[i1]])
        weights <- weights[-1,]
        rowstoinclude <- folds == i1
        predx <- x_list[[i3]]
        predx <- predx[rowstoinclude,]
        score <- t(weights%*%(t(predx)))[,1]
        PPS[rowstoinclude] <- score
      }
    }
    if (method[i2] == 999) {
      print("correlation PPS")
      for (i1 in 1:nfolds) {
        print(i1)
        rowstoinclude <- folds != i1
        subx <- x_list[[i3]]
        subx <- subx[rowstoinclude,]
        suby <- y_bmi[rowstoinclude]
        fits[[i1]] <- corr.test(subx, suby)$ci[,2]
      }
      PPS <- rep(NA, nrow(data))
      for ( i1 in 1:nfolds) { 
        weights <- fits[[i1]]
        rowstoinclude <- folds == i1
        predx <- x_list[[i3]]
        predx <- predx[rowstoinclude,]
        score <- t(weights%*%(t(predx)))[,1]
        PPS[rowstoinclude] <- score
      }
    }
    PPS_column <- which(colnames(data)=="PPS_Pers_Cor") + (i3 - 1) * length(method) + (i2 - 1)
    data[, PPS_column] <- PPS
  }
}

## GET PREDICTION POWER OF PPSs & PLOT RESULTS

# remove outlier
# data$PPS_outlier <- "ok"
# for (i2 in 1:nrow(data)) {
#   for (i1 in which(colnames(data)=="PPS_Pers_Cor") : which(colnames(data)=="PPS_EatItems_Cor")) {
#     if (data[i2,i1] < -5 * sd(data[,i1]) | data[i2,i1] > 5 * sd(data[,i1])) {
#       data[i2,]$PPS_outlier <- "outlier"
#     }
#   }
# }
# table(data$PPS_outlier)
# # data <- subset(data, PPS_outlier == "ok") # uncomment to remove outliers

# calculate correlations and write into data frame
PPS_BMIcorr <- data.frame()

nPPs <- length(catname) * length(method) # how many PPS were calculated before?
for (i in 1:nPPs) {
  PPS_BMIcorr <- rbind(PPS_BMIcorr, corr.test(data$BMI95, data[, (which(colnames(data)=="PPS_Pers_Cor")+i-1)])$ci)
}
PPS_BMIcorr$PPS <- c(rep("Personality Scales", length(method)), rep("Cognition", length(method)), 
                     rep("Cortical Thickness", length(method)), rep("Cortical Area", length(method)),
                     rep("Cortical Volume", length(method)), rep("Personality Items", length(method)), 
                     rep("Eating Items", length(method)))
PPS_BMIcorr$Method <- c(rep(c("Correlation", "Lasso a=.01", "Lasso a=.1", "Lasso a=.5", "Lasso a=1"), length(catname)))

# plot

theme_set(theme_bw())

PPS_BMIcorr$Method = factor(PPS_BMIcorr$Method, levels = c("Correlation", "Lasso a=.01", "Lasso a=.1", "Lasso a=.5", "Lasso a=1"))

ggplot(PPS_BMIcorr, aes(x=reorder(PPS, r), y = r, ymin = lower, ymax = upper,  group = Method, color = Method))  +
  geom_point(position = position_dodge(width = .5), size = 4) +
  ylim(-.1, .5) +
  scale_x_discrete(name="", limits=rev) +
  geom_linerange(position = position_dodge(width = .5), size = 1.2) +
  geom_hline(aes(yintercept = 0))  + 
  labs(colour="Method") +
  ggtitle("Obesity-PPS Correlations (Singletons)")

# CONCLUSION: SIMPLEST METHOD - CORRELATIONS - YIELDS BEST RESULTS.

## SAVE DATA INCL PPS
save(data, file="rsaves/data-BL-singletons_2022-03-29.Rda")
write.csv(data, file="rsaves/data-BL-singletons_2022-03-29.csv", row.names = FALSE)
rm(list = ls())

############################################################################

#########################
#
# Part IV: PPS IN TWINS
#
######################### ----

rm(list = ls())

## "SWITCH BOARD"
#-

## PREPARATION

# set wd
setwd("~/Job/Tartu Traineeship/R Analysis/abcd_bmi")

# packages
library(psych)
library(ggplot2)
library(lme4)
library(lmerTest)
library(dplyr)

# read files
data <- read.csv("rsaves/data-BL-clean_2022-02-18.csv", sep=",", dec=".", header=TRUE) # read cleaned Baseline
data_raw <- data

## TWIN CORRELATIONS PPS-BMI

data <- subset(data_raw, select = c(ID, sex, age, twinstatus, family, BMI95:cbcl_q53_p))
data$family <- as.factor(data$family)

nrow(subset(data, twinstatus == 'singleton')) # N train sample
nrow(subset(data, twinstatus != 'singleton')) # N test sample

#build structures
y_bmi <- data[, which(colnames(data)=="BMI95")]
#y_rfmp <- data[, which(colnames(data)=="RFMp")]
x_pers <- as.matrix(data[, (which(colnames(data)=="upps_negurgency")) : (which(colnames(data)=="prosociality"))])
x_cog <- as.matrix(data[, (which(colnames(data)=="nih_picvoc")) : (which(colnames(data)=="lmt_RT"))])
x_ct <- as.matrix(data[, (which(colnames(data)=="smri_thick_cdk_banksstslh")) : (which(colnames(data)=="smri_thick_cdk_insularh"))])
x_ca <- as.matrix(data[, (which(colnames(data)=="smri_area_cdk_banksstslh")) : (which(colnames(data)=="smri_area_cdk_insularh"))])
x_cv <- as.matrix(data[, (which(colnames(data)=="smri_vol_cdk_banksstslh")) : (which(colnames(data)=="smri_vol_cdk_insularh"))])
x_persitems <- as.matrix(data[, (which(colnames(data)=="upps6_y")) : (which(colnames(data)=="prosocial_q3_y"))])
x_eatitems <- as.matrix(data[, (which(colnames(data)=="cbcl_q24_p")) : (which(colnames(data)=="cbcl_q53_p"))])

x_list <- list()
x_list[[1]] <- x_pers
x_list[[2]] <- x_cog
x_list[[3]] <- x_ct
x_list[[4]] <- x_ca
x_list[[5]] <- x_cv
x_list[[6]] <- x_persitems
x_list[[7]] <- x_eatitems

data$PPS_PersScales <- NA # new cols
data$PPS_Cog <- NA
data$PPS_CT <- NA
data$PPS_CA <- NA
data$PPS_CV <- NA
data$PPS_PersItems <- NA
data$PPS_EatItems <- NA
data$doesnteatwell <- data$cbcl_q42_p # name Eating items for later; not in PPS calc needed
data$overeating <- data$cbcl_q53_p 

catname <- c('personality_scales', 'cognition', 'corticalthickness', 'corticalarea', 'corticalvolume', 'personality_items', 'eating_items')
fits <- list()

for (i in 1:length(catname)) {
  print(catname[i])
  containsNA <- rowSums(is.na(x_list[[i]])) > 0
  rowstoinclude <- data$twinstatus == 'singleton' & containsNA == F
  subx <- x_list[[i]]
  subx <- subx[rowstoinclude,]
  suby <- y_bmi[rowstoinclude]
  fits[[i]] <- corr.test(subx, suby)$ci[,2]
  PPS <- rep(NA, nrow(data))
  weights <- fits[[i]]
  rowstoinclude <- data$twinstatus != 'singleton' & containsNA == F
  predx <- x_list[[i]]
  predx <- predx[rowstoinclude,]
  score <- t(weights%*%(t(predx)))[,1]
  PPS[rowstoinclude] <- score
  PPS_column <- which(colnames(data)=="PPS_PersScales") + (i - 1)
  data[, PPS_column] <- PPS
}


data <- subset(data, twinstatus != 'singleton', select=c(ID:BMI95, PPS_PersScales:overeating))
data$family <- as.factor(data$family)

PPS <- c("Personality Scales", "Cognition", "Cortical Thickness", "Cortical Surface Area", "Cortical Volume", "Personality Items", "Eating Items")

PPS_BMIcorr <- data.frame(cbind(PPS, 
  rbind(table(is.na(x_pers)[,1])[1], table(is.na(x_cog)[,1])[1], table(is.na(x_ct)[,1])[1], table(is.na(x_ca)[,1])[1], table(is.na(x_cv)[,1])[1], table(is.na(x_persitems)[,1])[1], table(is.na(x_eatitems)[,1])[1]),
  rbind(
    c(fixef(lmer(scale(BMI95) ~ scale(PPS_PersScales) + (1 | family), data = data))[2],
      confint(lmer(scale(BMI95) ~ scale(PPS_PersScales) + (1 | family), data = data))[4,]),
    c(fixef(lmer(scale(BMI95) ~ scale(PPS_Cog) + (1 | family), data = data))[2],
      confint(lmer(scale(BMI95) ~ scale(PPS_Cog) + (1 | family), data = data))[4,]),
    c(fixef(lmer(scale(BMI95) ~ scale(PPS_CT) + (1 | family), data = data))[2],
      confint(lmer(scale(BMI95) ~ scale(PPS_CT) + (1 | family), data = data))[4,]),
    c(fixef(lmer(scale(BMI95) ~ scale(PPS_CA) + (1 | family), data = data))[2],
      confint(lmer(scale(BMI95) ~ scale(PPS_CA) + (1 | family), data = data))[4,]),
    c(fixef(lmer(scale(BMI95) ~ scale(PPS_CV) + (1 | family), data = data))[2],
      confint(lmer(scale(BMI95) ~ scale(PPS_CV) + (1 | family), data = data))[4,]),
    c(fixef(lmer(scale(BMI95) ~ scale(PPS_PersItems) + (1 | family), data = data))[2],
      confint(lmer(scale(BMI95) ~ scale(PPS_PersItems) + (1 | family), data = data))[4,]),
    c(fixef(lmer(scale(BMI95) ~ scale(PPS_EatItems) + (1 | family), data = data))[2],
      confint(lmer(scale(BMI95) ~ scale(PPS_EatItems) + (1 | family), data = data))[4,]))))
names(PPS_BMIcorr) <- c("PPS", "N_train+test", "B", "lower", "upper")
PPS_BMIcorr[,2:5] <- lapply(PPS_BMIcorr[,2:5], as.character)
PPS_BMIcorr[,2:5] <- lapply(PPS_BMIcorr[,2:5], as.numeric)
#PPS_BMIcorr$Significance <- ifelse(PPS_BMIcorr$lower > 0, "P < 0.05", "n.s.")
print(PPS_BMIcorr)

# get Ns and Ps
summary(lmer(scale(BMI95) ~ scale(PPS_Cog) + (1 | family), data = data))
summary(lmer(scale(BMI95) ~ scale(PPS_CT) + (1 | family), data = data))
summary(lmer(scale(BMI95) ~ scale(PPS_CA) + (1 | family), data = data))
summary(lmer(scale(BMI95) ~ scale(PPS_CV) + (1 | family), data = data))
summary(lmer(scale(BMI95) ~ scale(PPS_PersItems) + (1 | family), data = data))
summary(lmer(scale(BMI95) ~ scale(PPS_EatItems) + (1 | family), data = data))

PPS_BMIcorr <- subset(PPS_BMIcorr, PPS != "Personality Scales")

# Plot
theme_set(theme_bw())
ggplot(PPS_BMIcorr, aes(x = reorder(PPS, B), y = B, ymin = lower, ymax = upper))  +
  geom_point(size = 4) +
  ylim(0.0, 0.45) +
  scale_x_discrete(name="", limits=rev) +
  geom_linerange(size = 1.4) +
  geom_hline(aes(yintercept = 0))  +
  #ggtitle("Obesity-PPS Correlations (Twins & Sibs)") +
  theme(text = element_text(size=14), legend.position="bottom", legend.title = element_blank())

# Correlation Matrix: doesn't control for family!
nrow(subset(na.omit(data), select=c(BMI95, PPS_Cog:PPS_EatItems)))
cor(subset(data, select=c(BMI95, PPS_Cog:PPS_EatItems)), use="complete.obs")

# Correlations with lmer() and p-Values
summary(lmer(scale(BMI95) ~ scale(PPS_EatItems) + (1 | family), data = data))
summary(lmer(scale(BMI95) ~ scale(PPS_PersItems) + (1 | family), data = data))
summary(lmer(scale(BMI95) ~ scale(PPS_Cog) + (1 | family), data = data))
summary(lmer(scale(BMI95) ~ scale(PPS_CT) + (1 | family), data = data))
summary(lmer(scale(BMI95) ~ scale(PPS_CA) + (1 | family), data = data))
summary(lmer(scale(BMI95) ~ scale(PPS_CV) + (1 | family), data = data))

summary(lmer(scale(PPS_EatItems) ~ scale(PPS_PersItems) + (1 | family), data = data))
summary(lmer(scale(PPS_EatItems) ~ scale(PPS_Cog) + (1 | family), data = data))
summary(lmer(scale(PPS_EatItems) ~ scale(PPS_CT) + (1 | family), data = data))
summary(lmer(scale(PPS_EatItems) ~ scale(PPS_CA) + (1 | family), data = data))
summary(lmer(scale(PPS_EatItems) ~ scale(PPS_CV) + (1 | family), data = data))

summary(lmer(scale(PPS_PersItems) ~ scale(PPS_Cog) + (1 | family), data = data))
summary(lmer(scale(PPS_PersItems) ~ scale(PPS_CT) + (1 | family), data = data))
summary(lmer(scale(PPS_PersItems) ~ scale(PPS_CA) + (1 | family), data = data))
summary(lmer(scale(PPS_PersItems) ~ scale(PPS_CV) + (1 | family), data = data))

summary(lmer(scale(PPS_Cog) ~ scale(PPS_CT) + (1 | family), data = data))
summary(lmer(scale(PPS_Cog) ~ scale(PPS_CA) + (1 | family), data = data))
summary(lmer(scale(PPS_Cog) ~ scale(PPS_CV) + (1 | family), data = data))

summary(lmer(scale(PPS_CT) ~ scale(PPS_CA) + (1 | family), data = data))
summary(lmer(scale(PPS_CT) ~ scale(PPS_CV) + (1 | family), data = data))

summary(lmer(scale(PPS_CA) ~ scale(PPS_CV) + (1 | family), data = data))

## SAVE DATA
save(data, file="rsaves/data-BL-twinPPS_2022-03-29.Rda")
write.csv(data, file="rsaves/data-BL-twinPPS_2022-03-29.csv", row.names = FALSE)
save(PPS_BMIcorr, file="rsaves/data-BL-twinPPS-correlations_2022-03-29.Rda")
write.csv(PPS_BMIcorr, file="rsaves/data-BL-twinPPS-correlations_2022-03-29.csv", row.names = FALSE)
#rm(list = ls())
#load(file="rsaves/data-BL-twinPPS_2022-03-29.Rda")

### WRITE WIDE DATA FILES

data$family <- as.factor(data$family)
data$family <- droplevels(data$family)

pairID <- rep(NA, nrow(data))
familyvector <- (levels(data$family))

for (i in 1:length(familyvector)) {
  famnr <- familyvector[i]
  pairtoinclude <- data$family == famnr
  if (table(pairtoinclude)[2] == 2){
    pairID[pairtoinclude] <- c(1,2)
  }
  else {
    print(paste('Error within family', i))
  }
}

data$pairID <- pairID
table(data$pairID)

data <- data[is.na(data$pairID)==F,]

wide <- reshape(data, idvar = c("family", "twinstatus"), timevar = "pairID", direction = "wide", sep = "_T")

# sanity check
nrow(subset(wide, ID_T1 != ID_T2)) # all pair should have 2 different IDs
cor(wide$BMI95_T1, wide$BMI95_T2) # should correlate, but be different from another
cor(wide$BMI95_T1, wide$PPS_EatItems_T2, use="complete.obs") # should not correlate
cor(wide$BMI95_T1, wide$PPS_EatItems_T1, use="complete.obs") # should correlate around r=.40

## DoC FULL Data Set
wideAll <- subset(wide, select = c(twinstatus, BMI95_T1, PPS_Cog_T1, PPS_CT_T1, PPS_CA_T1, PPS_CV_T1, PPS_PersItems_T1, PPS_EatItems_T1, doesnteatwell_T1, overeating_T1, BMI95_T2, PPS_Cog_T2, PPS_CT_T2, PPS_CA_T2, PPS_CV_T2, PPS_PersItems_T2, PPS_EatItems_T2, doesnteatwell_T2, overeating_T2))
wideAll$zyg <- wideAll$twinstatus
wideAll <- wideAll[,-1]
save(wideAll, file="rsaves/wideAll_2022-03-29.Rda")
write.csv(wideAll, file="rsaves/wideAll_2022-03-29.csv", row.names = FALSE)

# additional sanity check: is BMI heritable?
cor(wideAll %>% filter(zyg=="MZ") %>% select(BMI95_T1,BMI95_T2) )
cor(wideAll %>% filter(zyg=="DZ") %>% select(BMI95_T1,BMI95_T2) )

############################################################################

#########################
#
# Part V: DoC
#
######################### ----

rm(list = ls())

## "SWITCH BOARD"
#-

## PREPARATION

# set wd
setwd("~/Job/Tartu Traineeship/R Analysis/abcd_bmi")

# packages
library(ggplot2)
library(dplyr)
library(umx)

# read files
data <- read.csv("rsaves/wideAll_2022-03-29.csv", sep=",", dec=".", header=TRUE) # read Wide PPS data
data_raw <- data #for reload: data <- data_raw


## UMX PREP

table(data$zyg)
levels(data$zyg) <- c("DZ", "MZ", "DZ") # overrides Siblings as DZ
table(data$zyg)

data <- umx_scale(data)

mzData = data[data$zyg %in% "MZ", ]
dzData = data[data$zyg %in% "DZ", ]

# Calculate Ns
print(nrow(na.omit(subset(mzData, select=c(BMI95_T1, BMI95_T2, PPS_Cog_T1, PPS_Cog_T2)))))
print(nrow(na.omit(subset(dzData, select=c(BMI95_T1, BMI95_T2, PPS_Cog_T1, PPS_Cog_T2)))))
print(nrow(na.omit(subset(mzData, select=c(BMI95_T1, BMI95_T2, PPS_CT_T1, PPS_CT_T2)))))
print(nrow(na.omit(subset(dzData, select=c(BMI95_T1, BMI95_T2, PPS_CT_T1, PPS_CT_T2)))))
print(nrow(na.omit(subset(mzData, select=c(BMI95_T1, BMI95_T2, PPS_CA_T1, PPS_CA_T2)))))
print(nrow(na.omit(subset(dzData, select=c(BMI95_T1, BMI95_T2, PPS_CA_T1, PPS_CA_T2)))))
print(nrow(na.omit(subset(mzData, select=c(BMI95_T1, BMI95_T2, PPS_CV_T1, PPS_CV_T2)))))
print(nrow(na.omit(subset(dzData, select=c(BMI95_T1, BMI95_T2, PPS_CV_T1, PPS_CV_T2)))))
print(nrow(na.omit(subset(mzData, select=c(BMI95_T1, BMI95_T2, PPS_PersItems_T1, PPS_PersItems_T2)))))
print(nrow(na.omit(subset(dzData, select=c(BMI95_T1, BMI95_T2, PPS_PersItems_T1, PPS_PersItems_T2)))))
print(nrow(na.omit(subset(mzData, select=c(BMI95_T1, BMI95_T2, PPS_EatItems_T1, PPS_EatItems_T2)))))
print(nrow(na.omit(subset(dzData, select=c(BMI95_T1, BMI95_T2, PPS_EatItems_T1, PPS_EatItems_T2)))))

## ACE ANALYSIS

ace_BMI = umxACE(selDVs = "BMI95", sep = "_T", dzData = dzData, mzData = mzData)
ace_Cog = umxACE(selDVs = "PPS_Cog", sep = "_T", dzData = dzData, mzData = mzData)
ace_CT = umxACE(selDVs = "PPS_CT", sep = "_T", dzData = dzData, mzData = mzData)
ace_CA = umxACE(selDVs = "PPS_CA", sep = "_T", dzData = dzData, mzData = mzData)
ace_CV = umxACE(selDVs = "PPS_CV", sep = "_T", dzData = dzData, mzData = mzData)
ace_PersItems = umxACE(selDVs = "PPS_PersItems", sep = "_T", dzData = dzData, mzData = mzData)
ace_EatItems = umxACE(selDVs = "PPS_EatItems", sep = "_T", dzData = dzData, mzData = mzData)

# Genetic and environmental correlations
biACE_Cog = umxACE(selDVs = c("BMI95", "PPS_Cog"), sep = "_T", dzData = dzData, mzData = mzData)
umxSummaryACE(biACE_Cog, showRg = TRUE, std = T)
biACE_CT = umxACE(selDVs = c("BMI95", "PPS_CT"), sep = "_T", dzData = dzData, mzData = mzData)
umxSummaryACE(biACE_CT, showRg = TRUE, std = T)
biACE_CA = umxACE(selDVs = c("BMI95", "PPS_CA"), sep = "_T", dzData = dzData, mzData = mzData)
umxSummaryACE(biACE_CA, showRg = TRUE, std = T)
biACE_CV = umxACE(selDVs = c("BMI95", "PPS_CV"), sep = "_T", dzData = dzData, mzData = mzData)
umxSummaryACE(biACE_CV, showRg = TRUE, std = T)
biACE_PPS_PersItems = umxACE(selDVs = c("BMI95", "PPS_PersItems"), sep = "_T", dzData = dzData, mzData = mzData)
umxSummaryACE(biACE_PPS_PersItems, showRg = TRUE, std = T)
biACE_PPS_EatItems = umxACE(selDVs = c("BMI95", "PPS_EatItems"), sep = "_T", dzData = dzData, mzData = mzData)
umxSummaryACE(biACE_PPS_EatItems, showRg = TRUE, std = T)

## DOC ANALYSIS

# prep results data frame
docres <- data.frame(matrix(ncol = 10, nrow = 24))
colnames(docres) <- c("PPS", "Model", "ChiSquared", "AIC", "a2bBeta", "a2b_lowerCI", "a2b_upperCI", "b2aBeta", "b2a_lowerCI", "b2a_upperCI")
docres$PPS <- c(rep("Cogntion", 4), rep("CorticalThickness", 4), rep("CorticalArea", 4), rep("CorticalVolume", 4), rep("GeneralBehavior", 4), rep("EatingBehavior", 4))
docres$Model <- rep(c("Correlated", "BMI-to-PPS", "PPS-to-BMI", "Reciprocal"), 6)
doccompare <- list()

# set variable names
var1 = "BMI95"
var2vec <- c("PPS_Cog", "PPS_CT", "PPS_CA", "PPS_CV", "PPS_PersItems", "PPS_EatItems")

# do DoC

for (i in 1:length(var2vec)) {
  
  print(var2vec[i])
  var2 = var2vec[i]
  
  # Baseline DoC
  DoC  = umxDoC(var1= var1, var2= var2, mzData= mzData, dzData= dzData, sep = "_T", causal= TRUE, autoRun=F)
  parnames <- names(coef(DoC))
  fixlist <- parnames[7:12]
  
  # Correlated
  Chol = umxDoC(var1= var1, var2= var2, mzData= mzData, dzData= dzData, sep = "_T", causal= FALSE, autoRun=F)
  # fix factor loadings to 1
  Chol = umxModify(Chol, c("FacLoad_r1c1","FacLoad_r2c2"), free = FALSE, value=1.0, name = "Chol", autoRun=F)
  # fix residuals as cs es to zero
  Chol <- umxModify(Chol, fixlist, free = FALSE, value=0, name = "Chol", autoRun=F)
  # free up the latent factor e's
  Chol = umxModify(Chol, c("e_r1c1","e_r2c2"), free = TRUE, value=1.0, name = "Chol", autoRun=T)
  # make reference model
  CholRef <- mxRefModels(Chol, run=TRUE)
  # get fit
  CholSum <- summary(Chol, refModels=CholRef)
  
  # A2B = BMI to PPS
  a2b   = umxModify(DoC, "a2b", free = TRUE, name = "a2b", autoRun=F)
  # fix factor loadings to 1
  a2b = umxModify(a2b, c("FacLoad_r1c1","FacLoad_r2c2"), free = FALSE, value=1.0, name = "a2b", autoRun=F)
  # fix residuals as cs es to zero
  a2b <- umxModify(a2b, fixlist, free = FALSE, value=0, name = "a2b", autoRun=F)
  # free up the latent factor e's
  a2b = umxModify(a2b, c("e_r1c1","e_r2c2"), free = TRUE, value=1.0, name = "a2b", autoRun=T)
  # make reference model
  a2bRef <- mxRefModels(a2b, run=TRUE)
  # get fit
  a2bSum <- summary(a2b, refModels=a2bRef)
  a2bB <- umxConfint(a2b, parm='all', run=T)
  
  # B2A = PPS to BMI
  b2a   = umxModify(DoC, "b2a", free = TRUE, name = "b2a", autoRun=F)
  # fix factor loadings to 1
  b2a = umxModify(b2a, c("FacLoad_r1c1","FacLoad_r2c2"), free = FALSE, value=1.0, name = "b2a", autoRun=F)
  # fix residuals as cs es to zero
  b2a <- umxModify(b2a, fixlist, free = FALSE, value=0, name = "b2a", autoRun=F)
  # free up the latent factor e's
  b2a = umxModify(b2a, c("e_r1c1","e_r2c2"), free = TRUE, value=1.0, name = "b2a", autoRun=T)
  # make reference model
  b2aRef <- mxRefModels(b2a, run=TRUE)
  # get fit
  b2aSum <- summary(b2a, refModels=b2aRef)
  b2aB <- umxConfint(b2a, parm='all', run=T)
  
  # Reciprocal
  Recip   = umxModify(DoC, c("a2b", "b2a"), free = TRUE, name = "Recip", autoRun=F)
  # fix factor loadings to 1
  Recip = umxModify(Recip, c("FacLoad_r1c1","FacLoad_r2c2"), free = FALSE, value=1.0, name = "Recip", autoRun=F)
  # fix residuals as cs es to zero
  Recip <- umxModify(Recip, fixlist, free = FALSE, value=0, name = "Recip", autoRun=F)
  # free up the latent factor e's
  Recip = umxModify(Recip, c("e_r1c1","e_r2c2"), free = TRUE, value=1.0, name = "Recip", autoRun=T)
  # make reference model
  RecipRef <- mxRefModels(Recip, run=TRUE)
  # get fit
  RecipSum <- summary(Recip, refModels=RecipRef)
  RecipB <- umxConfint(Recip, parm='all', run=T)
  
  # write into data frame
  
  docres[1+(i-1)*4, 3] <- CholSum$Chi
  docres[1+(i-1)*4, 4] <- extractAIC(Chol)
  docres[1+(i-1)*4, 5] <- NA
  
  docres[2+(i-1)*4, 3] <- a2bSum$Chi
  docres[2+(i-1)*4, 4] <- extractAIC(a2b)
  docres[2+(i-1)*4, 5] <- a2bB$output$confidenceIntervals[7,2]
  docres[2+(i-1)*4, 6] <- a2bB$output$confidenceIntervals[7,1]
  docres[2+(i-1)*4, 7] <- a2bB$output$confidenceIntervals[7,3]
  
  docres[3+(i-1)*4, 3] <- b2aSum$Chi
  docres[3+(i-1)*4, 4] <- extractAIC(b2a)
  docres[3+(i-1)*4, 8] <- b2aB$output$confidenceIntervals[7,2]
  docres[3+(i-1)*4, 9] <- b2aB$output$confidenceIntervals[7,1]
  docres[3+(i-1)*4, 10] <- b2aB$output$confidenceIntervals[7,3]
  
  docres[4+(i-1)*4, 3] <- RecipSum$Chi
  docres[4+(i-1)*4, 4] <- extractAIC(Recip)
  docres[4+(i-1)*4, 5] <- RecipB$output$confidenceIntervals[7,2]
  docres[4+(i-1)*4, 6] <- RecipB$output$confidenceIntervals[7,1]
  docres[4+(i-1)*4, 7] <- RecipB$output$confidenceIntervals[7,3]
  docres[4+(i-1)*4, 8] <- RecipB$output$confidenceIntervals[8,2]
  docres[4+(i-1)*4, 9] <- RecipB$output$confidenceIntervals[8,1]
  docres[4+(i-1)*4, 10] <- RecipB$output$confidenceIntervals[8,3]
  
  # Compare
  print(var2vec[i])
  doccompare[[i*2-1]] <- umxCompare(Chol, c(a2b, b2a, Recip))
  doccompare[[i*2]] <- umxCompare(Recip, c(a2b, b2a))
}

### SHOW RESULTS

print(docres)

# Cognition
print(doccompare[[1]])
print(doccompare[[2]])
# Cortical Thickness
print(doccompare[[3]])
print(doccompare[[4]])
# Cortical Surface Area
print(doccompare[[5]])
print(doccompare[[6]])
# Cortical Volume
print(doccompare[[7]])
print(doccompare[[8]])
# Personality
print(doccompare[[9]])
print(doccompare[[10]])
# Eating
print(doccompare[[11]])
print(doccompare[[12]])

### MAKE GRAPH

docres$Model = factor(docres$Model, levels = c("Correlated", "BMI-to-PPS", "PPS-to-BMI", "Reciprocal"))
docres$PPS = factor(docres$PPS, levels = c("Cogntion", "CorticalThickness", "CorticalArea", "CorticalVolume", "GeneralBehavior", "EatingBehavior"))

theme_set(theme_bw())

ggplot(data=docres, aes(x=Model, y=ChiSquared, fill=Model)) +
  geom_bar(stat="identity", position=position_dodge()) +
  facet_wrap(~ PPS, scales="free_y")

ggplot(data=docres, aes(x=Model, y=AIC, fill=Model)) +
  geom_bar(stat="identity", position=position_dodge()) +
  facet_wrap(~ PPS, scales="free_y")

docbeta <- subset(docres, Model == "BMI-to-PPS" | Model == "PPS-to-BMI")

ggplot(data=docbeta, aes(x=Model, y=Beta)) +
  geom_bar(stat="identity", position=position_dodge()) +
  facet_wrap(~ PPS, scales="free_y")


############################################################################

#########################
#
# Part VI: CROSS LAGGED PANEL MODELS
#
######################### ----

rm(list = ls())

## "SWITCH BOARD"
#-

## PREPARATION

# set wd
setwd("~/Job/Tartu Traineeship/R Analysis/abcd_bmi")

# packages
library(psych)
library(ggplot2)
library(lavaan)
library(lavaan.survey)
library(lme4)
library(lmerTest)
library(lessR)

# set seed
set.seed(777)

# read files

data1 <- read.csv("rsaves/data-BL-orig_2022-02-18.csv", sep=",", dec=".", header=TRUE) # read Baseline
data2 <- read.csv("rsaves/data-1yFU-orig_2022-02-18.csv", sep=",", dec=".", header=TRUE) # read 1y FU
data3 <- read.csv("rsaves/data-2yFU-orig_2022-02-18.csv", sep=",", dec=".", header=TRUE) # read 2y FU

## DROP VARIABLES NOT IN THE DATA SET

# for Baseline:
data1 <- data1[,-c(which(colnames(data1)=="socialresponsiveness"):which(colnames(data1)=="eat_aggression"))] # delete instruments not in the set
data1 <- data1[,-which(colnames(data1)=="bisbas5_y")] # delete BIS/BAS item that was never answered

# for 1-year-FU:
data2 <- data2[,-c(which(colnames(data2)=="eat_attention"):which(colnames(data2)=="bis_drive"))] 
data2 <- data2[,-c((which(colnames(data2)=="Sep_CT")+1):(which(colnames(data2)=="Sep_CA")-1))]
data2 <- data2[,-c((which(colnames(data2)=="Sep_CA")+1):(which(colnames(data2)=="Sep_CV")-1))]
data2 <- data2[,-c((which(colnames(data2)=="Sep_CV")+1):(which(colnames(data2)=="Sep_PersItems")-1))]

# for 2-year-FU:
data3 <- data3[,-which(colnames(data3)=="socialresponsiveness")]
data3 <- data3[,-c(which(colnames(data3)=="nih_list"):which(colnames(data3)=="nih_card"))] 
data3 <- data3[,-which(colnames(data3)=="matrixreasoning")]
data3 <- data3[,-which(colnames(data3)=="posaffect")]
data3 <- data3[,-which(colnames(data3)=="bisbas5_y")]

datalist <- list()
datalist[[1]] <- data1
datalist[[2]] <- data2
datalist[[3]] <- data3

# model 1 for personality and cognition
mod1 <- data[,i] ~ data$sex*data$puberty + data$age + data$race + data$hispanic + data$handedness + data$site
# model 2 for cortical thickness
mod2 <- data[,i] ~ data$sex*data$puberty + data$age + data$race + data$hispanic + data$handedness + data$site + data$meanthickness
# model 3 for cortical area
mod3 <- data[,i] ~ data$sex*data$puberty + data$age + data$race + data$hispanic + data$handedness + data$site + data$totalarea
# model 4 for cortical volume
mod4 <- data[,i] ~ data$sex*data$puberty + data$age + data$race + data$hispanic + data$handedness + data$site + data$totalvolume

# other potential controls: data$married, data$singleParent, data$stimulants, data$income, data$collgemat, data$collgepat, data$sex_menstruation

set.seed(777)

for (j in 1:3) {
  print(j)
  data <- datalist[[j]]
  print(dim(data))
  data_raw <- data
  
  ## EXCLUDE CASES
  
  data <- data[is.na(data$sex)==F & +
                 is.na(data$puberty)==F & +
                 is.na(data$race)==F & +
                 is.na(data$hispanic)==F & +
                 is.na(data$age)==F & +
                 #is.na(data$RFMp)==F & +
                 is.na(data$BMI95)==F,]
  NrExcluded <- as.numeric(nrow(data_raw) - nrow(data))
  SumExcluded <- NrExcluded
  print(c("Excluded for Missing Important Values: ", NrExcluded))
  
  data <- data[data$diabetes == 0 & data$braininjury == 0 & +
                 data$cancer == 0 & data$cerebralpalsy == 0 & +
                 data$epilepsy == 0,]
  NrExcluded <- as.numeric(nrow(data_raw) - nrow(data)) - SumExcluded
  SumExcluded <- SumExcluded + NrExcluded
  print(c("Excluded for Medical Conditions: ", NrExcluded))
  
  data <- data[data$BMIz <= 5,] # & data$RFMp > 0,]
  NrExcluded <- as.numeric(nrow(data_raw) - nrow(data)) - SumExcluded
  SumExcluded <- SumExcluded + NrExcluded
  print(c("Excluded for low/irrational high BMI (and RFMp): ", NrExcluded))
  
  data <- data[data$BMIz > -2,]
  NrExcluded <- as.numeric(nrow(data_raw) - nrow(data)) - SumExcluded
  SumExcluded <- SumExcluded + NrExcluded
  print(c("Excluded for Underweight (BMI 'wasted'): ", NrExcluded))
  
  data$birthcontrol[is.na(data$birthcontrol) == T] <- 0
  data <- data[data$birthcontrol == 0,]
  NrExcluded <- as.numeric(nrow(data_raw) - nrow(data)) - SumExcluded
  SumExcluded <- SumExcluded + NrExcluded
  print(c("Excluded for taking birth control: ", NrExcluded))
  
  # work on Eating items
  data <- data[,-which(colnames(data)=="cbcl_q55_p")] #delete item "Overweight"
  data$cbcl_q24_p <- as.factor(data$cbcl_q24_p) # make Eating items 0/1 bc of low freq of "2"
  levels(data$cbcl_q24_p) <- c(0, 1, 1)
  data$cbcl_q24_p <- as.numeric(as.character(data$cbcl_q24_p))
  data$cbcl_q53_p <- as.factor(data$cbcl_q53_p)
  levels(data$cbcl_q53_p) <- c(0, 1, 1)
  data$cbcl_q53_p <- as.numeric(as.character(data$cbcl_q53_p))
  
  # Delete MRI data, that doesn't pass quality control
  if (j != 2) {
    data$mriqc <- ifelse(data$mriqc==1, 1, 0)
    table(data$mriqc) # excluded by MRI QC 445, 203, 515
    tempdat1 <- subset(data, mriqc==1)
    tempdat2 <- subset(data, mriqc!=1)
    tempdat2[, which(colnames(tempdat2)=="Sep_CT") : which(colnames(tempdat2)=="Sep_PersItems")] <- NA
    data <- rbind(tempdat1, tempdat2)
  }
  
  ## RESIDUALISE
  
  data$sex <- as.factor(data$sex)
  data$sex <- relevel(data$sex, ref = "0")
  data$race <- as.factor(data$race)
  data$race <- relevel(data$race, ref = "0")
  data$hispanic <- as.factor(data$hispanic)
  data$hispanic <- relevel(data$hispanic, ref = "0")
  data$handedness <- as.factor(data$handedness)
  data$handedness <- relevel(data$handedness, ref = "1")
  data$site <- as.factor(data$site)
  data$site <- relevel(data$site, ref = "site16")
  
  i <- which(colnames(data)=="BMI95") 
    data[,i] <- residuals(lm(mod1, na.action = na.exclude)) 

  for (i in (which(colnames(data)=="Sep_PersScales")+1) : (which(colnames(data)=="Sep_Cognition")-1)) {
    data[,i] <- residuals(lm(mod1, na.action = na.exclude)) }

  for (i in (which(colnames(data)=="Sep_Cognition")+1) : (which(colnames(data)=="Sep_CT")-1)) {
    data[,i] <- residuals(lm(mod1, na.action = na.exclude)) }
  
  if (j != 2) {
  
    for (i in (which(colnames(data)=="Sep_CT")+1) : (which(colnames(data)=="Sep_CA")-1)) {
      data[,i] <- residuals(lm(mod2, na.action = na.exclude)) }
  
    for (i in (which(colnames(data)=="Sep_CA")+1) : (which(colnames(data)=="Sep_CV")-1)) {
      data[,i] <- residuals(lm(mod3, na.action = na.exclude)) }
  
    for (i in (which(colnames(data)=="Sep_CV")+1) : (which(colnames(data)=="Sep_PersItems")-1)) {
      data[,i] <- residuals(lm(mod4, na.action = na.exclude)) }
    
  }
  
  for (i in (which(colnames(data)=="Sep_PersItems")+1) : (which(colnames(data)=="Sep_Eating")-1)) {
    data[,i] <- residuals(lm(mod1, na.action = na.exclude)) }

  for (i in (which(colnames(data)=="Sep_Eating")+1) : (which(colnames(data)=="Sep_Calculations")-1)) {
    data[,i] <- residuals(lm(mod1, na.action = na.exclude)) }
  
  if (j != 2) {
    data <- subset(data, select = c(ID, Sep_IdControls, family, zygosity1, Sep_Obesity, BMI95, Sep_Cognition:nih_flanker,  nih_pattern:ravlt_delayedrecall, lmt_correct:Sep_PersItems, upps6_y:Sep_Calculations))
  }
  if (j == 2) {
    data <- subset(data, select = c(ID, Sep_IdControls, family, zygosity1, Sep_Obesity, BMI95, Sep_PersItems, cbcl_q01_p:Sep_Calculations))
  }
  
  # names and write in list
  namesvec <- names(data)
  namesvec <- paste0(namesvec, j)
  names(data) <- c("ID", namesvec[2:length(namesvec)])
  
  datalist[[j]] <- data
}

data1 <- datalist[[1]]
data2 <- datalist[[2]]
data3 <- datalist[[3]]

data <- merge(data1, data3, by="ID", all = F)
data <- data[,-c(which(colnames(data)=="Sep_IdControls3"), which(colnames(data)=="family3"), which(colnames(data)=="zygosity13"))]

datatri <- subset(data, select = c(ID, Sep_IdControls1, family1, zygosity11, Sep_Obesity1, BMI951, Sep_PersItems1, cbcl_q01_p1:Sep_Calculations1, Sep_Obesity3, BMI953, Sep_PersItems3, cbcl_q01_p3:Sep_Calculations3))
datatri <- merge(datatri, data2, by="ID", all = F)
datatri <- subset(datatri, select = c(ID:Sep_Calculations1, Sep_Obesity2:Sep_Calculations2, Sep_Obesity3:Sep_Calculations3))

## only singletons?

data$famcount <- NA
data$family1 <- as.factor(data$family1)
data <- subset(data, is.na(family1)==F)
for (i in 1:nrow(data)) {
  famnr <- as.numeric(as.character(data[i,]$family1))
  famcnt <- nrow(subset(data, family1 == famnr))
  data[i,]$famcount <- famcnt }
datatri$famcount <- NA
datatri$family1 <- as.factor(datatri$family1)
datatri <- subset(datatri, is.na(family1)==F)
for (i in 1:nrow(datatri)) {
  famnr <- as.numeric(as.character(datatri[i,]$family1))
  famcnt <- nrow(subset(datatri, family1 == famnr))
  datatri[i,]$famcount <- famcnt }

#save(data, file="rsaves/CLPM-2times-raw_2022-03-31.Rda")
#write.csv(data, file="rsaves/CLPM-2times-raw_2022-03-31.csv", row.names = FALSE)
#save(datatri, file="rsaves/CLPM-3times-raw_2022-03-31.Rda")
#write.csv(datatri, file="rsaves/CLPM-3times-raw_2022-03-31.csv", row.names = FALSE)

# Shortcut to skip previous
rm(list = ls())
data <- read.csv("rsaves/CLPM-2times-raw_2022-03-31.csv", sep=",", dec=".", header=TRUE)
data <- subset(data, famcount==1)
set.seed(777)

## USING ONLY 2 TIMES

#build structures

nfolds <- 10
folds <- sample(1:nfolds, nrow(data), replace=T)

y_bmi <- data[, which(colnames(data)=="BMI951")]
x_cog1 <- as.matrix(data[, (which(colnames(data)=="nih_picvoc1")) : (which(colnames(data)=="lmt_RT1"))])
x_cog3 <- as.matrix(data[, (which(colnames(data)=="nih_picvoc3")) : (which(colnames(data)=="lmt_RT3"))])
x_ct1 <- as.matrix(data[, (which(colnames(data)=="smri_thick_cdk_banksstslh1")) : (which(colnames(data)=="smri_thick_cdk_insularh1"))])
x_ct3 <- as.matrix(data[, (which(colnames(data)=="smri_thick_cdk_banksstslh3")) : (which(colnames(data)=="smri_thick_cdk_insularh3"))])
x_ca1 <- as.matrix(data[, (which(colnames(data)=="smri_area_cdk_banksstslh1")) : (which(colnames(data)=="smri_area_cdk_insularh1"))])
x_ca3 <- as.matrix(data[, (which(colnames(data)=="smri_area_cdk_banksstslh3")) : (which(colnames(data)=="smri_area_cdk_insularh3"))])
x_cv1 <- as.matrix(data[, (which(colnames(data)=="smri_vol_cdk_banksstslh1")) : (which(colnames(data)=="smri_vol_cdk_insularh1"))])
x_cv3 <- as.matrix(data[, (which(colnames(data)=="smri_vol_cdk_banksstslh3")) : (which(colnames(data)=="smri_vol_cdk_insularh3"))])
x_persitems1 <- as.matrix(data[, (which(colnames(data)=="upps6_y1")) : (which(colnames(data)=="prosocial_q3_y1"))])
x_persitems3 <- as.matrix(data[, (which(colnames(data)=="upps6_y3")) : (which(colnames(data)=="prosocial_q3_y3"))])
x_eatitems1 <- as.matrix(data[, (which(colnames(data)=="cbcl_q24_p1")) : (which(colnames(data)=="cbcl_q53_p1"))])
x_eatitems3 <- as.matrix(data[, (which(colnames(data)=="cbcl_q24_p3")) : (which(colnames(data)=="cbcl_q53_p3"))])

x1_list <- list()
x1_list[[1]] <- x_cog1
x1_list[[2]] <- x_ct1
x1_list[[3]] <- x_ca1
x1_list[[4]] <- x_cv1
x1_list[[5]] <- x_persitems1
x1_list[[6]] <- x_eatitems1
x3_list <- list()
x3_list[[1]] <- x_cog3
x3_list[[2]] <- x_ct3
x3_list[[3]] <- x_ca3
x3_list[[4]] <- x_cv3
x3_list[[5]] <- x_persitems3
x3_list[[6]] <- x_eatitems3

data$PPS_Cog1 <- NA # new cols
data$PPS_CT1 <- NA
data$PPS_CA1 <- NA
data$PPS_CV1 <- NA
data$PPS_PersItems1 <- NA
data$PPS_EatItems1 <- NA
data$PPS_Cog3 <- NA 
data$PPS_CT3 <- NA
data$PPS_CA3 <- NA
data$PPS_CV3 <- NA
data$PPS_PersItems3 <- NA
data$PPS_EatItems3 <- NA

catname <- c('cognition', 'corticalthickness', 'corticalarea', 'corticalvolume', 'personality_items', 'eating_items')
fits <- list()

data$containsNA <- NA

for (i in 1:length(catname)) {
  print(catname[i])
  containsNA <- rowSums(is.na(x1_list[[i]])) > 0 | rowSums(is.na(x3_list[[i]])) > 0
  
  for (j in 1:nfolds) {
    rowstoinclude <- folds != j & containsNA == F
    subx <- x1_list[[i]]
    subx <- subx[rowstoinclude,]
    suby <- y_bmi[rowstoinclude]
    fits[[j]] <- corr.test(subx, suby)$ci[,2]
  }

  PPS1 <- rep(NA, nrow(data))
  PPS3 <- rep(NA, nrow(data))
  
  for (j in 1:nfolds) {
    weights <- fits[[j]]
    rowstoinclude <- folds == j & containsNA == F
    predx1 <- x1_list[[i]]
    predx1 <- predx1[rowstoinclude,]
    predx3 <- x3_list[[i]]
    predx3 <- predx3[rowstoinclude,]
    score1 <- t(weights%*%(t(predx1)))[,1]
    score3 <- t(weights%*%(t(predx3)))[,1]
    PPS1[rowstoinclude] <- score1 
    PPS3[rowstoinclude] <- score3 
  }
  
  PPS1_column <- which(colnames(data)=="PPS_Cog1") + (i - 1)
  data[, PPS1_column] <- PPS1
  PPS3_column <- which(colnames(data)=="PPS_Cog3") + (i - 1)
  data[, PPS3_column] <- PPS3
}

cor(data$BMI951, data$BMI953, use="complete.obs")
cor(data[,c(which(colnames(data)=="BMI951"), which(colnames(data)=="BMI953"), 774:785)], use="complete.obs")

PPSvars <- c("PPS_Cog", "PPS_CT", "PPS_CA", "PPS_CV", "PPS_PersItems", "PPS_EatItems")
sumlist <- list()
data_backup <- data

## Define model for lavaan
clpm <- '
# Easiest model
x2 ~ x1 + y1
y2 ~ x1 + y1
'

for (j in 1:length(PPSvars)) {
  
  selvars <- c("BMI951", "BMI953", paste0(PPSvars[j],1),  paste0(PPSvars[j],3))
  data_clpm <- data[selvars]
  names(data_clpm) <- c("x1", "x2", "y1", "y2")
  data_clpm <- na.omit(data_clpm)
  for (i in 2:ncol(data_clpm)) {
    data_clpm[,i] <- scale(data_clpm[,i])
  }
  print(PPSvars[j])
  print(nrow(data_clpm))
  fit <- sem(clpm, data = data_clpm, missing = 'listwise', estimator="MLM")
  summary(fit, fit.measures = T, rsquare=T, standardized=T)
  print(standardizedSolution(fit, type="std.all", ci=T))
}


# Shortcut to skip previous
rm(list = ls())
datatri <- read.csv("rsaves/CLPM-3times-raw_2022-03-29.csv", sep=",", dec=".", header=TRUE)
datatri <- subset(datatri, famcount==1)
set.seed(777)

## USING ALL 3 TIMES

data <- datatri

#build structures

nfolds <- 10
folds <- sample(1:nfolds, nrow(data), replace=T)

y_bmi <- data[, which(colnames(data)=="BMI951")]
x_persitems1 <- as.matrix(data[, (which(colnames(data)=="Sep_PersItems1")+1) : (which(colnames(data)=="Sep_Eating1")-1)])
x_persitems2 <- as.matrix(data[, (which(colnames(data)=="Sep_PersItems2")+1) : (which(colnames(data)=="Sep_Eating2")-1)])
x_persitems3 <- as.matrix(data[, (which(colnames(data)=="Sep_PersItems3")+1) : (which(colnames(data)=="Sep_Eating3")-1)])
x_eatitems1 <- as.matrix(data[, (which(colnames(data)=="Sep_Eating1")+1) : (which(colnames(data)=="Sep_Calculations1")-1)])
x_eatitems2 <- as.matrix(data[, (which(colnames(data)=="Sep_Eating2")+1) : (which(colnames(data)=="Sep_Calculations2")-1)])
x_eatitems3 <- as.matrix(data[, (which(colnames(data)=="Sep_Eating3")+1) : (which(colnames(data)=="Sep_Calculations3")-1)])

x1_list <- list()
x1_list[[1]] <- x_persitems1
x1_list[[2]] <- x_eatitems1
x2_list <- list()
x2_list[[1]] <- x_persitems2
x2_list[[2]] <- x_eatitems2
x3_list <- list()
x3_list[[1]] <- x_persitems3
x3_list[[2]] <- x_eatitems3

data$PPS_PersItems1 <- NA # new cols
data$PPS_EatItems1 <- NA
data$PPS_PersItems2 <- NA
data$PPS_EatItems2 <- NA
data$PPS_PersItems3 <- NA
data$PPS_EatItems3 <- NA

catname <- c('personality_items', 'eating_items')
fits <- list()

for (i in 1:length(catname)) {
  print(catname[i])
  containsNA <- rowSums(is.na(x1_list[[i]])) > 0 | rowSums(is.na(x3_list[[i]]))
  
  for (j in 1:nfolds) {
    rowstoinclude <- folds != j & containsNA == F
    subx <- x1_list[[i]]
    subx <- subx[rowstoinclude,]
    suby <- y_bmi[rowstoinclude]
    fits[[j]] <- corr.test(subx, suby)$ci[,2]
  }
  
  PPS1 <- rep(NA, nrow(data))
  PPS2 <- rep(NA, nrow(data))
  PPS3 <- rep(NA, nrow(data))
  
  for (j in 1:nfolds) {
    weights <- fits[[j]]
    rowstoinclude <- folds == j & containsNA == F
    predx1 <- x1_list[[i]]
    predx1 <- predx1[rowstoinclude,]
    predx2 <- x2_list[[i]]
    predx2 <- predx2[rowstoinclude,]
    predx3 <- x3_list[[i]]
    predx3 <- predx3[rowstoinclude,]
    score1 <- t(weights%*%(t(predx1)))[,1]
    score2 <- t(weights%*%(t(predx2)))[,1]
    score3 <- t(weights%*%(t(predx3)))[,1]
    PPS1[rowstoinclude] <- score1 
    PPS2[rowstoinclude] <- score2
    PPS3[rowstoinclude] <- score3 
  }
  
  PPS1_column <- which(colnames(data)=="PPS_PersItems1") + (i - 1)
  data[, PPS1_column] <- PPS1
  PPS2_column <- which(colnames(data)=="PPS_PersItems2") + (i - 1)
  data[, PPS2_column] <- PPS2
  PPS3_column <- which(colnames(data)=="PPS_PersItems3") + (i - 1)
  data[, PPS3_column] <- PPS3
}

cor(data[,384:389], use="complete.obs")

PPSvars <- c("PPS_PersItems", "PPS_EatItems")
sumlist <- list()
data_backup <- data

# RI-CLPM Code from:
# Mund, M., & Nestler, S. (2019). Beyond the Cross-Lagged Panel Model: Next-generation statistical 
# tools for analyzing interdependencies across the life course. Advances in Life Course Research, 
# 41, 100249. https://doi.org/10.1016/j.alcr.2018.10.002

## Define model for lavaan
### note: in this implementation, we assume that the intercepts are 
### equal over time. in the model below, we will allow the intercepts
### to change over time---this second model is equivalent to the model 
### published by Hamaker et al.
riclpm <- '
# Define intercept factors
ix =~ 1*x1+1*x2+1*x3
iy =~ 1*y1+1*y2+1*y3
# Define phantom latent variables
etax1 =~ 1*x1
etax2 =~ 1*x2
etax3 =~ 1*x3
etay1 =~ 1*y1
etay2 =~ 1*y2
etay3 =~ 1*y3
# Autoregressive effects
etax2 ~ a1*etax1
etax3 ~ a1*etax2
etay2 ~ a2*etay1
etay3 ~ a2*etay2
# Crosslagged effects
etay2 ~ c1*etax1
etay3 ~ c1*etax2
etax2 ~ c2*etay1
etax3 ~ c2*etay2
# Some further constraints on the variance structure
# 1. Set error variances of the observed variables to zero
x1 ~~ 0*x1
x2 ~~ 0*x2
x3 ~~ 0*x3
y1 ~~ 0*y1
y2 ~~ 0*y2
y3 ~~ 0*y3
# 2. Let lavaan estimate the variance of the latent variables
etax1 ~~ varx1*etax1
etax2 ~~ varx2*etax2
etax3 ~~ varx3*etax3
etay1 ~~ vary1*etay1
etay2 ~~ vary2*etay2
etay3 ~~ vary3*etay3
# 3. We also want estimates of the intercept factor variances and an
#    estimate of their covariance
ix ~~ varix*ix
iy ~~ variy*iy
ix ~~ covi*iy
# 4. We have to define that the covariance between the intercepts and
#    the latents of the first time point are zero
etax1 ~~ 0*ix
etay1 ~~ 0*ix
etax1 ~~ 0*iy
etay1 ~~ 0*iy
# 5. Finally, we estimate the covariance between the latents of x and y
#    of the first time point, the second time-point and so on. note that
#    for the second to fourth time point the correlation is constrained to
#    the same value
etax1 ~~ cov1*etay1
etax2 ~~ e1*etay2
etax3 ~~ e1*etay3
# The model also contains a mean structure and we have to define some
# constraints for this part of the model. the assumption is that 
# the observed intercepts vary freely over time and we do not want to 
# estimate the means of the latent intercept factors
x1 ~ 1
x2 ~ 1
x3 ~ 1
y1 ~ 1
y2 ~ 1
y3 ~ 1
etax1 ~ 0*1
etax2 ~ 0*1
etax3 ~ 0*1
etay1 ~ 0*1
etay2 ~ 0*1
etay3 ~ 0*1
ix ~ 0*1
iy ~ 0*1
## define correlations
cori := covi / (sqrt(varix) * sqrt(variy))
cor1 := cov1 / (sqrt(varx1) * sqrt(vary1))
cort2 := e1 / (sqrt(varx2) * sqrt(vary2))
cort3 := e1 / (sqrt(varx3) * sqrt(vary3))
'

for (j in 1:length(PPSvars)) {
  
  selvars <- c("BMI951", "BMI952", "BMI953", paste0(PPSvars[j],1), paste0(PPSvars[j],2), paste0(PPSvars[j],3))
  data_clpm <- data[selvars]
  names(data_clpm) <- c("x1", "x2", "x3", "y1", "y2", "y3")
  data_clpm <- na.omit(data_clpm)
  for (i in 1:ncol(data_clpm)) {
    data_clpm[,i] <- scale(data_clpm[,i])
  }
  print(PPSvars[j])
  print(nrow(data_clpm))
  fit <- sem(riclpm, data = data_clpm)
  summary(fit, fit.measures = T, rsquare=TRUE)
  print(parameterEstimates(fit, ci=T))
}

##################################################################

#########################
#
# Part VII: SUMMARY CHART
#
#########################

rm(list = ls())

## "SWITCH BOARD"
#-

## PREPARATION

# set wd
setwd("~/Job/Tartu Traineeship/R Analysis/abcd_bmi")

# packages
library(ggplot2)
library(dplyr)
library(tidyverse)
library(forcats)

df <- data.frame(var1 = c(rep("Personality/Psychopathology",6), rep("Eating Behavior",6), rep("Cognition",4),
                          rep("Cortical Thickness",4), rep("Cortical Surface Area",4), rep("Cortical Volume",4)),
                 var2 = c(rep(c(rep("DoC",2), rep("Traditional CLPM",2), rep("RI-CLPM",2)),2), 
                          rep(c(rep("DoC",2), rep("Traditional CLPM",2)),4)),
                 var3 = rep(c("Obesity to PPS", "PPS to Obesity"),14),
                 Beta = c(NA,.09,.07,.03,.03,.04,.26,.16,.22,.05,.11,.02,.05,NA,-.04,-.01,.15,NA,.08,.00,.07,NA,.02,.00,NA,.07,.02,.00),
                 lower = c(NA,.05,.04,.01,-.05,.02,.20,.11,.18,.02,.03,.00,.00,NA,-.08,-.03,.11,NA,.05,-.02,.03,NA,.01,-.02,NA,.04,.00,-.02),
                 upper = c(NA,.12,.10,.05,.11,.07,.32,.20,.26,.08,.19,.05,.09,NA,-.01,.02,.20,NA,.11,.02,0.11,NA,.03,.02,NA,.11,.03,.02))

df$var1 <- factor(df$var1, levels=unique(df$var1))
df$var2 <- factor(df$var2, levels=unique(df$var2))

theme_set(theme_bw())

ggplot(data=df, aes(x=var2, y=Beta, fill=var3)) +
  geom_bar(stat="identity", position=position_dodge(width = .75))+
  geom_errorbar(aes(ymin = lower, ymax = upper), width = 0.2, size = 1, 
                position=position_dodge(width = .75)) + 
  labs(x = NULL) +
  theme(legend.position="bottom", legend.title = element_blank(),
        text = element_text(size=14)) +
  scale_fill_manual(values=c("grey47", "grey75"))+
  geom_hline(yintercept=0, linetype="dashed")+
  facet_wrap(~ var1, ncol=2)
```
